## Supplementary Table S1 for "Comparison of machine-learning and logistic regression models to predict 30-day unplanned readmission: a development and validation study"

| **Supplementary Table S1. Details and frequency of diagnosis codes (International Classification of Diseases 10th Revision codes)** | | | | | | | | | | | | | | | |
| --- | --- | --- | --- | --- | --- | --- | --- | --- | --- | --- | --- | --- | --- | --- | --- |
| Variables | Derivation dataset (N=339,513) | | | | | | Validation dataset (N=118,074) | | | | | | Use of the variable | | |
|  | Total  N=339,513 | | Outcome (-)  N=316,405 | | Outcome (+)  N=23,108 | | Total  N=118,074 | | Outcome (-)  N=110,567 | | Outcome (+)  N=7,507 | | Data pattern  1&2 | Data pattern  3&4 | Data pattern  5&6 |
|  | n | % | n | % | n | % | n | % | n | % | n | % |  |  |  |
| A02 Other salmonella infections | 76 | 0.02 | masked | masked | masked | masked | 26 | 0.02 | masked | masked | masked | masked |  |  | ✓ |
| A04 Other bacterial intestinal infections | 2143 | 0.63 | 1978 | 0.63 | 165 | 0.71 | 804 | 0.68 | 716 | 0.65 | 88 | 1.17 |  |  | ✓ |
| A08 Viral and other specified intestinal infections | 2499 | 0.74 | 2394 | 0.76 | 105 | 0.45 | 587 | 0.5 | 568 | 0.51 | 19 | 0.25 |  |  | ✓ |
| A09 Other gastroenteritis and colitis of infectious and unspecified origin | 14387 | 4.24 | 13347 | 4.22 | 1040 | 4.5 | 5042 | 4.27 | 4694 | 4.25 | 348 | 4.64 |  | ✓ | ✓ |
| A16 Respiratory tuberculosis, not confirmed bacteriologically or histologically | 174 | 0.05 | 152 | 0.05 | 22 | 0.1 | 79 | 0.07 | 66 | 0.06 | 13 | 0.17 |  |  | ✓ |
| A18 Tuberculosis of other organs | 53 | 0.02 | 47 | 0.01 | 6 | 0.03 | 14 | 0.01 | 12 | 0.01 | 2 | 0.03 |  |  | ✓ |
| A31 Infection due to other mycobacteria | 653 | 0.19 | 588 | 0.19 | 65 | 0.28 | 237 | 0.2 | 215 | 0.19 | 22 | 0.29 |  |  | ✓ |
| A37 Whooping cough | 53 | 0.02 | 48 | 0.02 | 5 | 0.02 | 15 | 0.01 | 14 | 0.01 | 1 | 0.01 |  |  | ✓ |
| A40 Streptococcal sepsis | 145 | 0.04 | 134 | 0.04 | 11 | 0.05 | 48 | 0.04 | 44 | 0.04 | 4 | 0.05 |  |  | ✓ |
| A41 Other sepsis | 5614 | 1.65 | 4805 | 1.52 | 809 | 3.5 | 1611 | 1.36 | 1416 | 1.28 | 195 | 2.6 |  | ✓ | ✓ |
| A46 Other sepsis | 108 | 0.03 | 106 | 0.03 | 2 | 0.01 | 62 | 0.05 | 62 | 0.06 | 0 | 0 |  |  | ✓ |
| A48 Other bacterial diseases, not elsewhere classified | 88 | 0.03 | 84 | 0.03 | 4 | 0.02 | 42 | 0.04 | 42 | 0.04 | 0 | 0 |  |  | ✓ |
| A49 Bacterial infection of unspecified site | 3060 | 0.9 | 2799 | 0.88 | 261 | 1.13 | 1149 | 0.97 | 1045 | 0.95 | 104 | 1.39 |  |  | ✓ |
| A56 Other sexually transmitted chlamydial diseases | 28 | 0.01 | 27 | 0.01 | 1 | 0 | 12 | 0.01 | 12 | 0.01 | 0 | 0 |  |  | ✓ |
| A60 Anogenital herpesviral [herpes simplex] infection | 103 | 0.03 | 100 | 0.03 | 3 | 0.01 | 42 | 0.04 | 39 | 0.04 | 3 | 0.04 |  |  | ✓ |
| A87 Viral meningitis | 104 | 0.03 | 100 | 0.03 | 4 | 0.02 | 25 | 0.02 | 25 | 0.02 | 0 | 0 |  |  | ✓ |
| B00 Eczema herpeticum | 728 | 0.21 | 684 | 0.22 | 44 | 0.19 | 224 | 0.19 | 214 | 0.19 | 10 | 0.13 |  |  | ✓ |
| B01 Varicella [chickenpox] | 59 | 0.02 | 57 | 0.02 | 2 | 0.01 | 18 | 0.02 | 16 | 0.01 | 2 | 0.03 |  |  | ✓ |
| B02 Zoster [herpes zoster] | 1814 | 0.53 | 1674 | 0.53 | 140 | 0.61 | 536 | 0.45 | 497 | 0.45 | 39 | 0.52 |  |  | ✓ |
| B07 Viral warts | 32 | 0.01 | 28 | 0.01 | 4 | 0.02 | 13 | 0.01 | 12 | 0.01 | 1 | 0.01 |  |  | ✓ |
| B08 Other viral infections characterized by skin and mucous membrane lesions, not elsewhere classified | 630 | 0.19 | 606 | 0.19 | 24 | 0.1 | 210 | 0.18 | 197 | 0.18 | 13 | 0.17 |  |  | ✓ |
| B09 Unspecified viral infection characterized by skin and mucous membrane lesions | 37 | 0.01 | 35 | 0.01 | 2 | 0.01 | 23 | 0.02 | 22 | 0.02 | 1 | 0.01 |  |  | ✓ |
| B15 Acute hepatitis A | 16 | 0 | 16 | 0.01 | 0 | 0 | 13 | 0.01 | 13 | 0.01 | 0 | 0 |  |  | ✓ |
| B16 Acute hepatitis B | 1092 | 0.32 | 1034 | 0.33 | 58 | 0.25 | 396 | 0.34 | 372 | 0.34 | 24 | 0.32 |  |  | ✓ |
| B17 Other acute viral hepatitis | 324 | 0.1 | 310 | 0.1 | 14 | 0.06 | 114 | 0.1 | 112 | 0.1 | 2 | 0.03 |  |  | ✓ |
| B18 Chronic viral hepatitis | 7307 | 2.15 | 6711 | 2.12 | 596 | 2.58 | 2263 | 1.92 | 2123 | 1.92 | 140 | 1.86 |  | ✓ | ✓ |
| B24 Unspecified human immunodeficiency virus [HIV] disease | 31 | 0.01 | 28 | 0.01 | 3 | 0.01 | 14 | 0.01 | 13 | 0.01 | 1 | 0.01 |  |  | ✓ |
| B25 Cytomegaloviral disease | 218 | 0.06 | 196 | 0.06 | 22 | 0.1 | 55 | 0.05 | 48 | 0.04 | 7 | 0.09 |  |  | ✓ |
| B27 Infectious mononucleosis | 302 | 0.09 | 300 | 0.09 | 2 | 0.01 | 124 | 0.11 | 120 | 0.11 | 4 | 0.05 |  |  | ✓ |
| B34 Viral infection of unspecified site | 431 | 0.13 | 415 | 0.13 | 16 | 0.07 | 159 | 0.13 | 151 | 0.14 | 8 | 0.11 |  |  | ✓ |
| B35 Dermatophytosis | 1259 | 0.37 | 1174 | 0.37 | 85 | 0.37 | 356 | 0.3 | 331 | 0.3 | 25 | 0.33 |  |  | ✓ |
| B36 Other superficial mycoses | 53 | 0.02 | 47 | 0.01 | 6 | 0.03 | 15 | 0.01 | 15 | 0.01 | 0 | 0 |  |  | ✓ |
| B37 Candidiasis | 1221 | 0.36 | 1073 | 0.34 | 148 | 0.64 | 465 | 0.39 | 430 | 0.39 | 35 | 0.47 |  |  | ✓ |
| B44 Aspergillosis | 475 | 0.14 | 387 | 0.12 | 88 | 0.38 | 179 | 0.15 | 151 | 0.14 | 28 | 0.37 |  |  | ✓ |
| B48 Other mycoses, not elsewhere classified | 816 | 0.24 | 743 | 0.23 | 73 | 0.32 | 329 | 0.28 | 308 | 0.28 | 21 | 0.28 |  |  | ✓ |
| B49 Unspecified mycosis | 713 | 0.21 | 619 | 0.2 | 94 | 0.41 | 142 | 0.12 | 119 | 0.11 | 23 | 0.31 |  |  | ✓ |
| B59 Pneumocystisiasis | 1685 | 0.5 | 1550 | 0.49 | 135 | 0.58 | 338 | 0.29 | 312 | 0.28 | 26 | 0.35 |  |  | ✓ |
| B86 Scabies | 19 | 0.01 | 17 | 0.01 | 2 | 0.01 | 17 | 0.01 | 13 | 0.01 | 4 | 0.05 |  |  | ✓ |
| B90 Sequelae of tuberculosis | 462 | 0.14 | 412 | 0.13 | 50 | 0.22 | 153 | 0.13 | 144 | 0.13 | 9 | 0.12 |  |  | ✓ |
| B99 Other and unspecified infectious diseases | 212 | 0.06 | 181 | 0.06 | 31 | 0.13 | 51 | 0.04 | 45 | 0.04 | 6 | 0.08 |  |  | ✓ |
| C01 Malignant neoplasm of base of tongue | 73 | 0.02 | 70 | 0.02 | 3 | 0.01 | 22 | 0.02 | 19 | 0.02 | 3 | 0.04 |  |  | ✓ |
| C02 Malignant neoplasm of other and unspecified parts of tongue | 266 | 0.08 | 252 | 0.08 | 14 | 0.06 | 102 | 0.09 | 97 | 0.09 | 5 | 0.07 |  |  | ✓ |
| C03 Malignant neoplasm of gum | 65 | 0.02 | 63 | 0.02 | 2 | 0.01 | 26 | 0.02 | 24 | 0.02 | 2 | 0.03 |  |  | ✓ |
| C04 Malignant neoplasm of floor of mouth | 68 | 0.02 | 65 | 0.02 | 3 | 0.01 | 15 | 0.01 | 15 | 0.01 | 0 | 0 |  |  | ✓ |
| C05 Malignant neoplasm of palate | 32 | 0.01 | 29 | 0.01 | 3 | 0.01 | 13 | 0.01 | 13 | 0.01 | 0 | 0 |  |  | ✓ |
| C06 Malignant neoplasm of other and unspecified parts of mouth | 52 | 0.02 | 48 | 0.02 | 4 | 0.02 | 18 | 0.02 | 16 | 0.01 | 2 | 0.03 |  |  | ✓ |
| C07 Malignant neoplasm of parotid gland | 136 | 0.04 | 128 | 0.04 | 8 | 0.03 | 42 | 0.04 | 40 | 0.04 | 2 | 0.03 |  |  | ✓ |
| C08 Malignant neoplasm of other and unspecified major salivary glands | 51 | 0.02 | 49 | 0.02 | 2 | 0.01 | 18 | 0.02 | 18 | 0.02 | 0 | 0 |  |  | ✓ |
| C10 Malignant neoplasm of oropharynx | 287 | 0.08 | 258 | 0.08 | 29 | 0.13 | 104 | 0.09 | 98 | 0.09 | 6 | 0.08 |  |  | ✓ |
| C11 Malignant neoplasm of nasopharynx | 144 | 0.04 | 134 | 0.04 | 10 | 0.04 | 42 | 0.04 | 40 | 0.04 | 2 | 0.03 |  |  | ✓ |
| C12 Malignant neoplasm of piriform sinus | 224 | 0.07 | 207 | 0.07 | 17 | 0.07 | 92 | 0.08 | 83 | 0.08 | 9 | 0.12 |  |  | ✓ |
| C13 Malignant neoplasm of hypopharynx | 347 | 0.1 | 322 | 0.1 | 25 | 0.11 | 171 | 0.14 | 148 | 0.13 | 23 | 0.31 |  |  | ✓ |
| C14 Malignant neoplasm of other and ill-defined sites in the lip, oral cavity and pharynx | 48 | 0.01 | 44 | 0.01 | 4 | 0.02 | 22 | 0.02 | 17 | 0.02 | 5 | 0.07 |  |  | ✓ |
| C15 Malignant neoplasm of oesophagus | 4374 | 1.29 | 4006 | 1.27 | 368 | 1.59 | 1636 | 1.39 | 1489 | 1.35 | 147 | 1.96 |  | ✓ | ✓ |
| C16 Malignant neoplasm of stomach | 13983 | 4.12 | 12552 | 3.97 | 1431 | 6.19 | 4361 | 3.69 | 3961 | 3.58 | 400 | 5.33 |  | ✓ | ✓ |
| C17 Malignant neoplasm of small intestine | 434 | 0.13 | 376 | 0.12 | 58 | 0.25 | 152 | 0.13 | 138 | 0.12 | 14 | 0.19 |  |  | ✓ |
| C18 Malignant neoplasm of colon | 12986 | 3.82 | 11994 | 3.79 | 992 | 4.29 | 4420 | 3.74 | 4087 | 3.7 | 333 | 4.44 |  | ✓ | ✓ |
| C19 Malignant neoplasm of rectosigmoid junction | 1220 | 0.36 | 1139 | 0.36 | 81 | 0.35 | 373 | 0.32 | 342 | 0.31 | 31 | 0.41 |  |  | ✓ |
| C20 Malignant neoplasm of rectum | 7101 | 2.09 | 6526 | 2.06 | 575 | 2.49 | 2102 | 1.78 | 1936 | 1.75 | 166 | 2.21 |  | ✓ | ✓ |
| C21 Malignant neoplasm of anus and anal canal | 97 | 0.03 | 87 | 0.03 | 10 | 0.04 | 60 | 0.05 | 57 | 0.05 | 3 | 0.04 |  |  | ✓ |
| C22 Malignant neoplasm of liver and intrahepatic bile ducts | 6303 | 1.86 | 5676 | 1.79 | 627 | 2.71 | 1992 | 1.69 | 1813 | 1.64 | 179 | 2.38 |  | ✓ | ✓ |
| C23 Malignant neoplasm of gallbladder | 883 | 0.26 | 722 | 0.23 | 161 | 0.7 | 302 | 0.26 | 245 | 0.22 | 57 | 0.76 |  |  | ✓ |
| C24 Malignant neoplasm of other and unspecified parts of biliary tract | 2704 | 0.8 | 2090 | 0.66 | 614 | 2.66 | 849 | 0.72 | 687 | 0.62 | 162 | 2.16 |  |  | ✓ |
| C25 Malignant neoplasm of pancreas | 5348 | 1.58 | 4317 | 1.36 | 1031 | 4.46 | 1987 | 1.68 | 1637 | 1.48 | 350 | 4.66 |  | ✓ | ✓ |
| C30 Malignant neoplasm of nasal cavity and middle ear | 53 | 0.02 | 50 | 0.02 | 3 | 0.01 | 12 | 0.01 | 12 | 0.01 | 0 | 0 |  |  | ✓ |
| C31 Malignant neoplasm of accessory sinuses | 88 | 0.03 | 77 | 0.02 | 11 | 0.05 | 26 | 0.02 | 26 | 0.02 | 0 | 0 |  |  | ✓ |
| C32 Malignant neoplasm of larynx | 579 | 0.17 | 540 | 0.17 | 39 | 0.17 | 213 | 0.18 | 198 | 0.18 | 15 | 0.2 |  |  | ✓ |
| C34 Malignant neoplasm of bronchus and lung | 14126 | 4.16 | 12872 | 4.07 | 1254 | 5.43 | 4737 | 4.01 | 4298 | 3.89 | 439 | 5.85 |  | ✓ | ✓ |
| C37 Malignant neoplasm of thymus | 215 | 0.06 | 200 | 0.06 | 15 | 0.06 | 83 | 0.07 | 79 | 0.07 | 4 | 0.05 |  |  | ✓ |
| C38 Malignant neoplasm of heart, mediastinum and pleura | 80 | 0.02 | 77 | 0.02 | 3 | 0.01 | 11 | 0.01 | 9 | 0.01 | 2 | 0.03 |  |  | ✓ |
| C40 Malignant neoplasm of bone and articular cartilage of limbs | 119 | 0.04 | 113 | 0.04 | 6 | 0.03 | 44 | 0.04 | 39 | 0.04 | 5 | 0.07 |  |  | ✓ |
| C41 Malignant neoplasm of bone and articular cartilage of other and unspecified sites | 151 | 0.04 | 132 | 0.04 | 19 | 0.08 | 18 | 0.02 | 18 | 0.02 | 0 | 0 |  |  | ✓ |
| C43 Malignant melanoma of skin | 175 | 0.05 | 166 | 0.05 | 9 | 0.04 | 44 | 0.04 | 39 | 0.04 | 5 | 0.07 |  |  | ✓ |
| C44 Other malignant neoplasms of skin | 351 | 0.1 | 339 | 0.11 | 12 | 0.05 | 127 | 0.11 | 124 | 0.11 | 3 | 0.04 |  |  | ✓ |
| C45 Mesothelioma | 184 | 0.05 | 167 | 0.05 | 17 | 0.07 | 31 | 0.03 | 26 | 0.02 | 5 | 0.07 |  |  | ✓ |
| C47 Malignant neoplasm of peripheral nerves and autonomic nervous system | 14 | 0 | 14 | 0 | 0 | 0 | 11 | 0.01 | 10 | 0.01 | 1 | 0.01 |  |  | ✓ |
| C48 Malignant neoplasm of retroperitoneum and peritoneum | 639 | 0.19 | 583 | 0.18 | 56 | 0.24 | 155 | 0.13 | 142 | 0.13 | 13 | 0.17 |  |  | ✓ |
| C49 Malignant neoplasm of other connective and soft tissue | 529 | 0.16 | 496 | 0.16 | 33 | 0.14 | 244 | 0.21 | 228 | 0.21 | 16 | 0.21 |  |  | ✓ |
| C50 Malignant neoplasm of breast | 7571 | 2.23 | 7175 | 2.27 | 396 | 1.71 | 2434 | 2.06 | 2306 | 2.09 | 128 | 1.71 |  | ✓ | ✓ |
| C51 Malignant neoplasm of vulva | 63 | 0.02 | 56 | 0.02 | 7 | 0.03 | 33 | 0.03 | 32 | 0.03 | 1 | 0.01 |  |  | ✓ |
| C52 Malignant neoplasm of vagina | 53 | 0.02 | 46 | 0.01 | 7 | 0.03 | 10 | 0.01 | 10 | 0.01 | 0 | 0 |  |  | ✓ |
| C53 Malignant neoplasm of cervix uteri | 1735 | 0.51 | 1608 | 0.51 | 127 | 0.55 | 722 | 0.61 | 658 | 0.6 | 64 | 0.85 |  |  | ✓ |
| C54 Malignant neoplasm of corpus uteri | 2662 | 0.78 | 2509 | 0.79 | 153 | 0.66 | 847 | 0.72 | 789 | 0.71 | 58 | 0.77 |  |  | ✓ |
| C55 Malignant neoplasm of uterus, part unspecified | 219 | 0.06 | 199 | 0.06 | 20 | 0.09 | 41 | 0.03 | 39 | 0.04 | 2 | 0.03 |  |  | ✓ |
| C56 Malignant neoplasm of ovary | 3186 | 0.94 | 2964 | 0.94 | 222 | 0.96 | 1130 | 0.96 | 1049 | 0.95 | 81 | 1.08 |  |  | ✓ |
| C57 Malignant neoplasm of other and unspecified female genital organs | 96 | 0.03 | 88 | 0.03 | 8 | 0.03 | 35 | 0.03 | 30 | 0.03 | 5 | 0.07 |  |  | ✓ |
| C60 Malignant neoplasm of penis | 30 | 0.01 | 27 | 0.01 | 3 | 0.01 | 22 | 0.02 | 21 | 0.02 | 1 | 0.01 |  |  | ✓ |
| C61 Malignant neoplasm of prostate | 5293 | 1.56 | 4816 | 1.52 | 477 | 2.06 | 2135 | 1.81 | 1962 | 1.77 | 173 | 2.3 |  | ✓ | ✓ |
| C62 Malignant neoplasm of testis | 243 | 0.07 | 238 | 0.08 | 5 | 0.02 | 81 | 0.07 | 79 | 0.07 | 2 | 0.03 |  |  | ✓ |
| C64 Malignant neoplasm of kidney, except renal pelvis | 1781 | 0.52 | 1660 | 0.52 | 121 | 0.52 | 721 | 0.61 | 666 | 0.6 | 55 | 0.73 |  |  | ✓ |
| C65 Malignant neoplasm of renal pelvis | 721 | 0.21 | 676 | 0.21 | 45 | 0.19 | 324 | 0.27 | 300 | 0.27 | 24 | 0.32 |  |  | ✓ |
| C66 Malignant neoplasm of ureter | 947 | 0.28 | 863 | 0.27 | 84 | 0.36 | 295 | 0.25 | 277 | 0.25 | 18 | 0.24 |  |  | ✓ |
| C67 Malignant neoplasm of bladder | 5626 | 1.66 | 5182 | 1.64 | 444 | 1.92 | 2098 | 1.78 | 1958 | 1.77 | 140 | 1.86 |  | ✓ | ✓ |
| C68 Malignant neoplasm of other and unspecified urinary organs | 51 | 0.02 | 50 | 0.02 | 1 | 0 | 22 | 0.02 | 22 | 0.02 | 0 | 0 |  |  | ✓ |
| C71 Malignant neoplasm of brain | 319 | 0.09 | 296 | 0.09 | 23 | 0.1 | 103 | 0.09 | 91 | 0.08 | 12 | 0.16 |  |  | ✓ |
| C73 Malignant neoplasm of thyroid gland | 1192 | 0.35 | 1152 | 0.36 | 40 | 0.17 | 391 | 0.33 | 381 | 0.34 | 10 | 0.13 |  |  | ✓ |
| C74 Malignant neoplasm of adrenal gland | 25 | 0.01 | 23 | 0.01 | 2 | 0.01 | 17 | 0.01 | 16 | 0.01 | 1 | 0.01 |  |  | ✓ |
| C77 Secondary and unspecified malignant neoplasm of lymph nodes | 6926 | 2.04 | 6117 | 1.93 | 809 | 3.5 | 2336 | 1.98 | 2089 | 1.89 | 247 | 3.29 |  | ✓ | ✓ |
| C78 Secondary malignant neoplasm of respiratory and digestive organs | 15182 | 4.47 | 12825 | 4.05 | 2357 | 10.2 | 4940 | 4.18 | 4181 | 3.78 | 759 | 10.11 |  | ✓ | ✓ |
| C79 Secondary malignant neoplasm of other and unspecified sites | 7329 | 2.16 | 6273 | 1.98 | 1056 | 4.57 | 2579 | 2.18 | 2214 | 2 | 365 | 4.86 |  | ✓ | ✓ |
| C80 Malignant neoplasm, without specification of site | 1523 | 0.45 | 1243 | 0.39 | 280 | 1.21 | 417 | 0.35 | 334 | 0.3 | 83 | 1.11 |  |  | ✓ |
| C81 Hodgkin lymphoma | 269 | 0.08 | 244 | 0.08 | 25 | 0.11 | 76 | 0.06 | 71 | 0.06 | 5 | 0.07 |  |  | ✓ |
| C82 Follicular lymphoma | 1113 | 0.33 | 1053 | 0.33 | 60 | 0.26 | 244 | 0.21 | 231 | 0.21 | 13 | 0.17 |  |  | ✓ |
| C83 Non-follicular lymphoma | 3295 | 0.97 | 3074 | 0.97 | 221 | 0.96 | 1226 | 1.04 | 1143 | 1.03 | 83 | 1.11 |  |  | ✓ |
| C84 Mature T/NK-cell lymphomas | 245 | 0.07 | 215 | 0.07 | 30 | 0.13 | 69 | 0.06 | 63 | 0.06 | 6 | 0.08 |  |  | ✓ |
| C85 Other and unspecified types of non-Hodgkin lymphoma | 1288 | 0.38 | 1194 | 0.38 | 94 | 0.41 | 400 | 0.34 | 376 | 0.34 | 24 | 0.32 |  |  | ✓ |
| C86 Other specified types of T/NK-cell lymphoma | 173 | 0.05 | 160 | 0.05 | 13 | 0.06 | 52 | 0.04 | 46 | 0.04 | 6 | 0.08 |  |  | ✓ |
| C88 Malignant immunoproliferative diseases | 288 | 0.08 | 270 | 0.09 | 18 | 0.08 | 63 | 0.05 | 59 | 0.05 | 4 | 0.05 |  |  | ✓ |
| C90 Multiple myeloma and malignant plasma cell neoplasms | 1226 | 0.36 | 1092 | 0.35 | 134 | 0.58 | 409 | 0.35 | 365 | 0.33 | 44 | 0.59 |  |  | ✓ |
| C91 Lymphoid leukaemia | 761 | 0.22 | 690 | 0.22 | 71 | 0.31 | 199 | 0.17 | 187 | 0.17 | 12 | 0.16 |  |  | ✓ |
| C92 Myeloid leukaemia | 1712 | 0.5 | 1517 | 0.48 | 195 | 0.84 | 498 | 0.42 | 449 | 0.41 | 49 | 0.65 |  |  | ✓ |
| C93 Monocytic leukaemia | 104 | 0.03 | 82 | 0.03 | 22 | 0.1 | 18 | 0.02 | 15 | 0.01 | 3 | 0.04 |  |  | ✓ |
| C95 Leukaemia of unspecified cell type | 40 | 0.01 | 35 | 0.01 | 5 | 0.02 | 17 | 0.01 | 14 | 0.01 | 3 | 0.04 |  |  | ✓ |
| D00 Carcinoma in situ of oral cavity, oesophagus and stomach | 65 | 0.02 | 61 | 0.02 | 4 | 0.02 | 34 | 0.03 | 34 | 0.03 | 0 | 0 |  |  | ✓ |
| D01 Carcinoma in situ of other and unspecified digestive organs | 48 | 0.01 | 47 | 0.01 | 1 | 0 | 24 | 0.02 | 22 | 0.02 | 2 | 0.03 |  |  | ✓ |
| D02 Carcinoma in situ of middle ear and respiratory system | 20 | 0.01 | 19 | 0.01 | 1 | 0 | 11 | 0.01 | 11 | 0.01 | 0 | 0 |  |  | ✓ |
| D04 Carcinoma in situ of skin | 37 | 0.01 | 35 | 0.01 | 2 | 0.01 | 15 | 0.01 | 15 | 0.01 | 0 | 0 |  |  | ✓ |
| D05 Carcinoma in situ of breast | 13 | 0 | 13 | 0 | 0 | 0 | 12 | 0.01 | 11 | 0.01 | 1 | 0.01 |  |  | ✓ |
| D06 Carcinoma in situ of cervix uteri | 154 | 0.05 | 152 | 0.05 | 2 | 0.01 | 48 | 0.04 | 48 | 0.04 | 0 | 0 |  |  | ✓ |
| D09 Carcinoma in situ of other and unspecified sites | 73 | 0.02 | 67 | 0.02 | 6 | 0.03 | 46 | 0.04 | 44 | 0.04 | 2 | 0.03 |  |  | ✓ |
| D10 Benign neoplasm of mouth and pharynx | 28 | 0.01 | 26 | 0.01 | 2 | 0.01 | 19 | 0.02 | 19 | 0.02 | 0 | 0 |  |  | ✓ |
| D11 Benign neoplasm of major salivary glands | 202 | 0.06 | 200 | 0.06 | 2 | 0.01 | 61 | 0.05 | 61 | 0.06 | 0 | 0 |  |  | ✓ |
| D12 Benign neoplasm of colon, rectum, anus and anal canal | 1522 | 0.45 | 1458 | 0.46 | 64 | 0.28 | 626 | 0.53 | 593 | 0.54 | 33 | 0.44 |  |  | ✓ |
| D13 Benign neoplasm of other and ill-defined parts of digestive system | 1063 | 0.31 | 1039 | 0.33 | 24 | 0.1 | 456 | 0.39 | 446 | 0.4 | 10 | 0.13 |  |  | ✓ |
| D14 Benign neoplasm of middle ear and respiratory system | 145 | 0.04 | 140 | 0.04 | 5 | 0.02 | 45 | 0.04 | 44 | 0.04 | 1 | 0.01 |  |  | ✓ |
| D15 Benign neoplasm of other and unspecified intrathoracic organs | 127 | 0.04 | 124 | 0.04 | 3 | 0.01 | 38 | 0.03 | 37 | 0.03 | 1 | 0.01 |  |  | ✓ |
| D16 Benign neoplasm of bone and articular cartilage | 83 | 0.02 | 83 | 0.03 | 0 | 0 | 29 | 0.02 | 29 | 0.03 | 0 | 0 |  |  | ✓ |
| D17 Benign lipomatous neoplasm | 222 | 0.07 | 218 | 0.07 | 4 | 0.02 | 74 | 0.06 | 74 | 0.07 | 0 | 0 |  |  | ✓ |
| D18 Haemangioma and lymphangioma, any site | 238 | 0.07 | 227 | 0.07 | 11 | 0.05 | 78 | 0.07 | 71 | 0.06 | 7 | 0.09 |  |  | ✓ |
| D21 Other benign neoplasms of connective and other soft tissue | 85 | 0.03 | 82 | 0.03 | 3 | 0.01 | 34 | 0.03 | 34 | 0.03 | 0 | 0 |  |  | ✓ |
| D23 Other benign neoplasms of skin | 75 | 0.02 | 71 | 0.02 | 4 | 0.02 | 27 | 0.02 | 25 | 0.02 | 2 | 0.03 |  |  | ✓ |
| D24 Benign neoplasm of breast | 68 | 0.02 | 68 | 0.02 | 0 | 0 | 19 | 0.02 | 18 | 0.02 | 1 | 0.01 |  |  | ✓ |
| D25 Leiomyoma of uterus | 3475 | 1.02 | 3399 | 1.07 | 76 | 0.33 | 1373 | 1.16 | 1351 | 1.22 | 22 | 0.29 |  | ✓ | ✓ |
| D27 Benign neoplasm of ovary | 1820 | 0.54 | 1786 | 0.56 | 34 | 0.15 | 671 | 0.57 | 661 | 0.6 | 10 | 0.13 |  |  | ✓ |
| D30 Benign neoplasm of urinary organs | 49 | 0.01 | 45 | 0.01 | 4 | 0.02 | 19 | 0.02 | 17 | 0.02 | 2 | 0.03 |  |  | ✓ |
| D32 Benign neoplasm of meninges | 260 | 0.08 | 247 | 0.08 | 13 | 0.06 | 95 | 0.08 | 94 | 0.09 | 1 | 0.01 |  |  | ✓ |
| D33 Benign neoplasm of brain and other parts of central nervous system | 34 | 0.01 | 34 | 0.01 | 0 | 0 | 14 | 0.01 | 13 | 0.01 | 1 | 0.01 |  |  | ✓ |
| D34 Benign neoplasm of thyroid gland | 186 | 0.05 | 182 | 0.06 | 4 | 0.02 | 52 | 0.04 | 51 | 0.05 | 1 | 0.01 |  |  | ✓ |
| D35 Benign neoplasm of other and unspecified endocrine glands | 243 | 0.07 | 236 | 0.07 | 7 | 0.03 | 84 | 0.07 | 82 | 0.07 | 2 | 0.03 |  |  | ✓ |
| D36 Benign neoplasm of other and unspecified sites | 81 | 0.02 | 80 | 0.03 | 1 | 0 | 31 | 0.03 | 31 | 0.03 | 0 | 0 |  |  | ✓ |
| D37 Neoplasm of uncertain or unknown behaviour of oral cavity and digestive organs | 2399 | 0.71 | 2254 | 0.71 | 145 | 0.63 | 883 | 0.75 | 819 | 0.74 | 64 | 0.85 |  |  | ✓ |
| D38 Neoplasm of uncertain or unknown behaviour of middle ear and respiratory and intrathoracic organs | 555 | 0.16 | 531 | 0.17 | 24 | 0.1 | 204 | 0.17 | 200 | 0.18 | 4 | 0.05 |  |  | ✓ |
| D39 Neoplasm of uncertain or unknown behaviour of female genital organs | 1559 | 0.46 | 1530 | 0.48 | 29 | 0.13 | 554 | 0.47 | 541 | 0.49 | 13 | 0.17 |  |  | ✓ |
| D40 Neoplasm of uncertain or unknown behaviour of male genital organs | 44 | 0.01 | 43 | 0.01 | 1 | 0 | 18 | 0.02 | 18 | 0.02 | 0 | 0 |  |  | ✓ |
| D41 Neoplasm of uncertain or unknown behaviour of urinary organs | 335 | 0.1 | 319 | 0.1 | 16 | 0.07 | 156 | 0.13 | 146 | 0.13 | 10 | 0.13 |  |  | ✓ |
| D43 Neoplasm of uncertain or unknown behaviour of brain and central nervous system | 280 | 0.08 | 267 | 0.08 | 13 | 0.06 | 108 | 0.09 | 103 | 0.09 | 5 | 0.07 |  |  | ✓ |
| D44 Neoplasm of uncertain or unknown behaviour of endocrine glands | 629 | 0.19 | 594 | 0.19 | 35 | 0.15 | 274 | 0.23 | 264 | 0.24 | 10 | 0.13 |  |  | ✓ |
| D46 Myelodysplastic syndromes | 2172 | 0.64 | 1790 | 0.57 | 382 | 1.65 | 644 | 0.55 | 525 | 0.47 | 119 | 1.59 |  |  | ✓ |
| D47 Other neoplasms of uncertain or unknown behaviour of lymphoid, haematopoietic and related tissue | 373 | 0.11 | 297 | 0.09 | 76 | 0.33 | 121 | 0.1 | 104 | 0.09 | 17 | 0.23 |  |  | ✓ |
| D48 Neoplasm of uncertain or unknown behaviour of other and unspecified sites | 1008 | 0.3 | 955 | 0.3 | 53 | 0.23 | 294 | 0.25 | 280 | 0.25 | 14 | 0.19 |  |  | ✓ |
| D50 Iron deficiency anaemia | 18420 | 5.43 | 16806 | 5.31 | 1614 | 6.98 | 6336 | 5.37 | 5824 | 5.27 | 512 | 6.82 | ✓ | ✓ | ✓ |
| D51 Vitamin B12 deficiency anaemia | 139 | 0.04 | 122 | 0.04 | 17 | 0.07 | 54 | 0.05 | 46 | 0.04 | 8 | 0.11 |  |  | ✓ |
| D52 Folate deficiency anaemia | 390 | 0.11 | 333 | 0.11 | 57 | 0.25 | 158 | 0.13 | 135 | 0.12 | 23 | 0.31 |  |  | ✓ |
| D53 Other nutritional anaemias | 163 | 0.05 | 140 | 0.04 | 23 | 0.1 | 64 | 0.05 | 58 | 0.05 | 6 | 0.08 |  |  | ✓ |
| D59 Acquired haemolytic anaemia | 292 | 0.09 | 262 | 0.08 | 30 | 0.13 | 114 | 0.1 | 98 | 0.09 | 16 | 0.21 |  |  | ✓ |
| D61 Other aplastic anaemias | 902 | 0.27 | 767 | 0.24 | 135 | 0.58 | 317 | 0.27 | 281 | 0.25 | 36 | 0.48 |  |  | ✓ |
| D62 Acute posthaemorrhagic anaemia | 2532 | 0.75 | 2361 | 0.75 | 171 | 0.74 | 709 | 0.6 | 667 | 0.6 | 42 | 0.56 |  |  | ✓ |
| D64 Other anaemias | 8311 | 2.45 | 7200 | 2.28 | 1111 | 4.81 | 2529 | 2.14 | 2207 | 2 | 322 | 4.29 |  | ✓ | ✓ |
| D65 Disseminated intravascular coagulation [defibrination syndrome] | 1612 | 0.47 | 1442 | 0.46 | 170 | 0.74 | 415 | 0.35 | 364 | 0.33 | 51 | 0.68 |  |  | ✓ |
| D66 Hereditary factor VIII deficiency | 65 | 0.02 | 48 | 0.02 | 17 | 0.07 | 13 | 0.01 | 10 | 0.01 | 3 | 0.04 |  |  | ✓ |
| D68 Other coagulation defects | 1136 | 0.33 | 1052 | 0.33 | 84 | 0.36 | 338 | 0.29 | 314 | 0.28 | 24 | 0.32 |  |  | ✓ |
| D69 Purpura and other haemorrhagic conditions | 3814 | 1.12 | 3368 | 1.06 | 446 | 1.93 | 1044 | 0.88 | 930 | 0.84 | 114 | 1.52 |  |  | ✓ |
| D70 Agranulocytosis | 7075 | 2.08 | 6374 | 2.01 | 701 | 3.03 | 2020 | 1.71 | 1804 | 1.63 | 216 | 2.88 |  | ✓ | ✓ |
| D72 Other disorders of white blood cells | 181 | 0.05 | 161 | 0.05 | 20 | 0.09 | 74 | 0.06 | 71 | 0.06 | 3 | 0.04 |  |  | ✓ |
| D73 Diseases of spleen | 93 | 0.03 | 81 | 0.03 | 12 | 0.05 | 47 | 0.04 | 41 | 0.04 | 6 | 0.08 |  |  | ✓ |
| D75 Other diseases of blood and blood-forming organs | 130 | 0.04 | 122 | 0.04 | 8 | 0.03 | 35 | 0.03 | 33 | 0.03 | 2 | 0.03 |  |  | ✓ |
| D76 Other specified diseases with participation of lymphoreticular and reticulohistiocytic tissue | 126 | 0.04 | 116 | 0.04 | 10 | 0.04 | 30 | 0.03 | 27 | 0.02 | 3 | 0.04 |  |  | ✓ |
| D80 Immunodeficiency with predominantly antibody defects | 106 | 0.03 | 98 | 0.03 | 8 | 0.03 | 42 | 0.04 | 35 | 0.03 | 7 | 0.09 |  |  | ✓ |
| D84 Other immunodeficiencies | 224 | 0.07 | 216 | 0.07 | 8 | 0.03 | 67 | 0.06 | 60 | 0.05 | 7 | 0.09 |  |  | ✓ |
| D86 Sarcoidosis | 260 | 0.08 | 237 | 0.07 | 23 | 0.1 | 83 | 0.07 | 79 | 0.07 | 4 | 0.05 |  |  | ✓ |
| D89 Other disorders involving the immune mechanism, not elsewhere classified | 142 | 0.04 | 133 | 0.04 | 9 | 0.04 | 67 | 0.06 | 60 | 0.05 | 7 | 0.09 |  |  | ✓ |
| E03 Other hypothyroidism | 4144 | 1.22 | 3743 | 1.18 | 401 | 1.74 | 1591 | 1.35 | 1448 | 1.31 | 143 | 1.9 |  | ✓ | ✓ |
| E04 Other nontoxic goitre | 481 | 0.14 | 468 | 0.15 | 13 | 0.06 | 163 | 0.14 | 156 | 0.14 | 7 | 0.09 |  |  | ✓ |
| E05 Thyrotoxicosis [hyperthyroidism] | 1323 | 0.39 | 1255 | 0.4 | 68 | 0.29 | 479 | 0.41 | 458 | 0.41 | 21 | 0.28 |  |  | ✓ |
| E06 Thyroiditis | 488 | 0.14 | 458 | 0.14 | 30 | 0.13 | 240 | 0.2 | 229 | 0.21 | 11 | 0.15 |  |  | ✓ |
| E10 Type 1 diabetes mellitus | 1167 | 0.34 | 1074 | 0.34 | 93 | 0.4 | 376 | 0.32 | 359 | 0.32 | 17 | 0.23 |  |  | ✓ |
| E11 Type 2 diabetes mellitus | 62028 | 18.27 | 57089 | 18.04 | 4939 | 21.37 | 22353 | 18.93 | 20670 | 18.69 | 1683 | 22.42 | ✓ | ✓ | ✓ |
| E13 Other specified diabetes mellitus | 723 | 0.21 | 635 | 0.2 | 88 | 0.38 | 281 | 0.24 | 236 | 0.21 | 45 | 0.6 |  |  | ✓ |
| E14 Unspecified diabetes mellitus | 3365 | 0.99 | 3144 | 0.99 | 221 | 0.96 | 1065 | 0.9 | 1005 | 0.91 | 60 | 0.8 |  |  | ✓ |
| E15 Nondiabetic hypoglycaemic coma | 48 | 0.01 | 45 | 0.01 | 3 | 0.01 | 10 | 0.01 | 10 | 0.01 | 0 | 0 |  |  | ✓ |
| E16 Other disorders of pancreatic internal secretion | 1079 | 0.32 | 996 | 0.31 | 83 | 0.36 | 374 | 0.32 | 355 | 0.32 | 19 | 0.25 |  |  | ✓ |
| E20 Hypoparathyroidism | 173 | 0.05 | 162 | 0.05 | 11 | 0.05 | 61 | 0.05 | 54 | 0.05 | 7 | 0.09 |  |  | ✓ |
| E21 Hyperparathyroidism and other disorders of parathyroid gland | 548 | 0.16 | 512 | 0.16 | 36 | 0.16 | 211 | 0.18 | 197 | 0.18 | 14 | 0.19 |  |  | ✓ |
| E22 Hyperfunction of pituitary gland | 158 | 0.05 | 144 | 0.05 | 14 | 0.06 | 80 | 0.07 | 73 | 0.07 | 7 | 0.09 |  |  | ✓ |
| E23 Hypofunction and other disorders of pituitary gland | 425 | 0.13 | 396 | 0.13 | 29 | 0.13 | 145 | 0.12 | 138 | 0.12 | 7 | 0.09 |  |  | ✓ |
| E26 Hyperaldosteronism | 117 | 0.03 | 114 | 0.04 | 3 | 0.01 | 48 | 0.04 | 47 | 0.04 | 1 | 0.01 |  |  | ✓ |
| E27 Other disorders of adrenal gland | 301 | 0.09 | 278 | 0.09 | 23 | 0.1 | 144 | 0.12 | 128 | 0.12 | 16 | 0.21 |  |  | ✓ |
| E28 Ovarian dysfunction | 222 | 0.07 | 206 | 0.07 | 16 | 0.07 | 88 | 0.07 | 86 | 0.08 | 2 | 0.03 |  |  | ✓ |
| E34 Other endocrine disorders | 106 | 0.03 | 105 | 0.03 | 1 | 0 | 45 | 0.04 | 45 | 0.04 | 0 | 0 |  |  | ✓ |
| E41 Nutritional marasmus | 99 | 0.03 | 85 | 0.03 | 14 | 0.06 | 42 | 0.04 | 37 | 0.03 | 5 | 0.07 |  |  | ✓ |
| E43 Unspecified severe protein-energy malnutrition | 12 | 0 | 12 | 0 | 0 | 0 | 11 | 0.01 | 10 | 0.01 | 1 | 0.01 |  |  | ✓ |
| E44 Protein-energy malnutrition of moderate and mild degree | 55 | 0.02 | 48 | 0.02 | 7 | 0.03 | 19 | 0.02 | 16 | 0.01 | 3 | 0.04 |  |  | ✓ |
| E46 Unspecified protein-energy malnutrition | 735 | 0.22 | 622 | 0.2 | 113 | 0.49 | 258 | 0.22 | 232 | 0.21 | 26 | 0.35 |  |  | ✓ |
| E51 Thiamine deficiency | 186 | 0.05 | 171 | 0.05 | 15 | 0.06 | 54 | 0.05 | 47 | 0.04 | 7 | 0.09 |  |  | ✓ |
| E53 Deficiency of other B group vitamins | 691 | 0.2 | 633 | 0.2 | 58 | 0.25 | 334 | 0.28 | 303 | 0.27 | 31 | 0.41 |  |  | ✓ |
| E54 Ascorbic acid deficiency | 282 | 0.08 | 253 | 0.08 | 29 | 0.13 | 116 | 0.1 | 110 | 0.1 | 6 | 0.08 |  |  | ✓ |
| E56 Other vitamin deficiencies | 951 | 0.28 | 876 | 0.28 | 75 | 0.32 | 151 | 0.13 | 142 | 0.13 | 9 | 0.12 |  |  | ✓ |
| E61 Deficiency of other nutrient elements | 84 | 0.02 | 75 | 0.02 | 9 | 0.04 | 106 | 0.09 | 99 | 0.09 | 7 | 0.09 |  |  | ✓ |
| E63 Other nutritional deficiencies | 728 | 0.21 | 618 | 0.2 | 110 | 0.48 | 290 | 0.25 | 251 | 0.23 | 39 | 0.52 |  |  | ✓ |
| E66 Obesity | 736 | 0.22 | 714 | 0.23 | 22 | 0.1 | 337 | 0.29 | 333 | 0.3 | 4 | 0.05 |  |  | ✓ |
| E71 Disorders of branched-chain amino-acid metabolism and fatty-acid metabolism | 194 | 0.06 | 150 | 0.05 | 44 | 0.19 | 40 | 0.03 | 30 | 0.03 | 10 | 0.13 |  |  | ✓ |
| E72 Other disorders of amino-acid metabolism | 922 | 0.27 | 746 | 0.24 | 176 | 0.76 | 290 | 0.25 | 225 | 0.2 | 65 | 0.87 |  |  | ✓ |
| E77 Disorders of glycoprotein metabolism | 162 | 0.05 | 144 | 0.05 | 18 | 0.08 | 47 | 0.04 | 40 | 0.04 | 7 | 0.09 |  |  | ✓ |
| E78 Disorders of lipoprotein metabolism and other lipidaemias | 47550 | 14.01 | 44997 | 14.22 | 2553 | 11.05 | 18836 | 15.95 | 17860 | 16.15 | 976 | 13 | ✓ | ✓ | ✓ |
| E79 Disorders of purine and pyrimidine metabolism | 13454 | 3.96 | 12459 | 3.94 | 995 | 4.31 | 5110 | 4.33 | 4719 | 4.27 | 391 | 5.21 |  | ✓ | ✓ |
| E83 Disorders of mineral metabolism | 2937 | 0.87 | 2651 | 0.84 | 286 | 1.24 | 1044 | 0.88 | 927 | 0.84 | 117 | 1.56 |  |  | ✓ |
| E85 Amyloidosis | 183 | 0.05 | 164 | 0.05 | 19 | 0.08 | 83 | 0.07 | 76 | 0.07 | 7 | 0.09 |  |  | ✓ |
| E86 Volume depletion | 16366 | 4.82 | 14909 | 4.71 | 1457 | 6.31 | 5482 | 4.64 | 5021 | 4.54 | 461 | 6.14 |  |  | ✓ |
| E87 Other disorders of fluid, electrolyte and acid-base balance | 10152 | 2.99 | 9110 | 2.88 | 1042 | 4.51 | 3653 | 3.09 | 3292 | 2.98 | 361 | 4.81 |  | ✓ | ✓ |
| E88 Other metabolic disorders | 6153 | 1.81 | 5326 | 1.68 | 827 | 3.58 | 1436 | 1.22 | 1263 | 1.14 | 173 | 2.3 |  | ✓ | ✓ |
| E89 Postprocedural endocrine and metabolic disorders, not elsewhere classified | 246 | 0.07 | 225 | 0.07 | 21 | 0.09 | 66 | 0.06 | 60 | 0.05 | 6 | 0.08 |  |  | ✓ |
| F01 Vascular dementia | 204 | 0.06 | 189 | 0.06 | 15 | 0.06 | 86 | 0.07 | 77 | 0.07 | 9 | 0.12 |  |  | ✓ |
| F03 Unspecified dementia | 5306 | 1.56 | 4757 | 1.5 | 549 | 2.38 | 3101 | 2.63 | 2802 | 2.53 | 299 | 3.98 |  | ✓ | ✓ |
| F05 Delirium, not induced by alcohol and other psychoactive substances | 1015 | 0.3 | 914 | 0.29 | 101 | 0.44 | 593 | 0.5 | 547 | 0.49 | 46 | 0.61 |  |  | ✓ |
| F06 Other mental disorders due to brain damage and dysfunction and to physical disease | 391 | 0.12 | 380 | 0.12 | 11 | 0.05 | 128 | 0.11 | 123 | 0.11 | 5 | 0.07 |  |  | ✓ |
| F10 Mental and behavioural disorders due to use of alcohol | 919 | 0.27 | 876 | 0.28 | 43 | 0.19 | 337 | 0.29 | 309 | 0.28 | 28 | 0.37 |  |  | ✓ |
| F19 Mental and behavioural disorders due to multiple drug use and use of other psychoactive substances | 41 | 0.01 | 39 | 0.01 | 2 | 0.01 | 20 | 0.02 | 19 | 0.02 | 1 | 0.01 |  |  | ✓ |
| F20 Schizophrenia | 4302 | 1.27 | 3818 | 1.21 | 484 | 2.09 | 1567 | 1.33 | 1411 | 1.28 | 156 | 2.08 |  | ✓ | ✓ |
| F22 Persistent delusional disorders | 51 | 0.02 | 48 | 0.02 | 3 | 0.01 | 15 | 0.01 | 15 | 0.01 | 0 | 0 |  |  | ✓ |
| F23 Acute and transient psychotic disorders | 35 | 0.01 | 31 | 0.01 | 4 | 0.02 | 19 | 0.02 | 16 | 0.01 | 3 | 0.04 |  |  | ✓ |
| F25 Schizoaffective disorders | 35 | 0.01 | 32 | 0.01 | 3 | 0.01 | 10 | 0.01 | 9 | 0.01 | 1 | 0.01 |  |  | ✓ |
| F31 Bipolar affective disorder | 450 | 0.13 | 413 | 0.13 | 37 | 0.16 | 165 | 0.14 | 156 | 0.14 | 9 | 0.12 |  |  | ✓ |
| F32 Depressive episode | 4992 | 1.47 | 4582 | 1.45 | 410 | 1.77 | 1949 | 1.65 | 1806 | 1.63 | 143 | 1.9 |  | ✓ | ✓ |
| F34 Persistent mood [affective] disorders | 168 | 0.05 | 155 | 0.05 | 13 | 0.06 | 58 | 0.05 | 54 | 0.05 | 4 | 0.05 |  |  | ✓ |
| F41 Other anxiety disorders | 4958 | 1.46 | 4548 | 1.44 | 410 | 1.77 | 1525 | 1.29 | 1406 | 1.27 | 119 | 1.59 |  | ✓ | ✓ |
| F43 Reaction to severe stress, and adjustment disorders | 217 | 0.06 | 201 | 0.06 | 16 | 0.07 | 55 | 0.05 | 52 | 0.05 | 3 | 0.04 |  |  | ✓ |
| F44 Dissociative [conversion] disorders | 155 | 0.05 | 142 | 0.04 | 13 | 0.06 | 63 | 0.05 | 61 | 0.06 | 2 | 0.03 |  |  | ✓ |
| F45 Somatoform disorders | 1196 | 0.35 | 1114 | 0.35 | 82 | 0.35 | 295 | 0.25 | 266 | 0.24 | 29 | 0.39 |  |  | ✓ |
| F48 Other neurotic disorders | 435 | 0.13 | 409 | 0.13 | 26 | 0.11 | 127 | 0.11 | 114 | 0.1 | 13 | 0.17 |  |  | ✓ |
| F50 Eating disorders | 258 | 0.08 | 237 | 0.07 | 21 | 0.09 | 72 | 0.06 | 68 | 0.06 | 4 | 0.05 |  |  | ✓ |
| F51 Nonorganic sleep disorders | 22 | 0.01 | 22 | 0.01 | 0 | 0 | 10 | 0.01 | 10 | 0.01 | 0 | 0 |  |  | ✓ |
| F60 Specific personality disorders | 38 | 0.01 | 36 | 0.01 | 2 | 0.01 | 19 | 0.02 | 19 | 0.02 | 0 | 0 |  |  | ✓ |
| F79 Unspecified mental retardation | 148 | 0.04 | 137 | 0.04 | 11 | 0.05 | 44 | 0.04 | 41 | 0.04 | 3 | 0.04 |  |  | ✓ |
| F80 Specific developmental disorders of speech and language | 43 | 0.01 | 40 | 0.01 | 3 | 0.01 | 17 | 0.01 | 16 | 0.01 | 1 | 0.01 |  |  | ✓ |
| F82 Specific developmental disorder of motor function | 88 | 0.03 | 81 | 0.03 | 7 | 0.03 | 13 | 0.01 | 12 | 0.01 | 1 | 0.01 |  |  | ✓ |
| F84 Pervasive developmental disorders | 145 | 0.04 | 133 | 0.04 | 12 | 0.05 | 48 | 0.04 | 47 | 0.04 | 1 | 0.01 |  |  | ✓ |
| F89 Unspecified disorder of psychological development | 39 | 0.01 | 34 | 0.01 | 5 | 0.02 | 28 | 0.02 | 26 | 0.02 | 2 | 0.03 |  |  | ✓ |
| F90 Hyperkinetic disorders | 38 | 0.01 | 37 | 0.01 | 1 | 0 | 22 | 0.02 | 22 | 0.02 | 0 | 0 |  |  | ✓ |
| G00 Bacterial meningitis, not elsewhere classified | 139 | 0.04 | 134 | 0.04 | 5 | 0.02 | 39 | 0.03 | 36 | 0.03 | 3 | 0.04 |  |  | ✓ |
| G03 Meningitis due to other and unspecified causes | 319 | 0.09 | 301 | 0.1 | 18 | 0.08 | 96 | 0.08 | 85 | 0.08 | 11 | 0.15 |  |  | ✓ |
| G04 Encephalitis, myelitis and encephalomyelitis | 162 | 0.05 | 150 | 0.05 | 12 | 0.05 | 71 | 0.06 | 70 | 0.06 | 1 | 0.01 |  |  | ✓ |
| G06 Intracranial and intraspinal abscess and granuloma | 83 | 0.02 | 79 | 0.02 | 4 | 0.02 | 24 | 0.02 | 23 | 0.02 | 1 | 0.01 |  |  | ✓ |
| G09 Sequelae of inflammatory diseases of central nervous system | 55 | 0.02 | 47 | 0.01 | 8 | 0.03 | 13 | 0.01 | 8 | 0.01 | 5 | 0.07 |  |  | ✓ |
| G12 Spinal muscular atrophy and related syndromes | 344 | 0.1 | 317 | 0.1 | 27 | 0.12 | 150 | 0.13 | 140 | 0.13 | 10 | 0.13 |  |  | ✓ |
| G20 Parkinson disease | 2439 | 0.72 | 2167 | 0.68 | 272 | 1.18 | 815 | 0.69 | 724 | 0.65 | 91 | 1.21 |  |  | ✓ |
| G21 Secondary parkinsonism | 169 | 0.05 | 155 | 0.05 | 14 | 0.06 | 56 | 0.05 | 54 | 0.05 | 2 | 0.03 |  |  | ✓ |
| G23 Other degenerative diseases of basal ganglia | 153 | 0.05 | 132 | 0.04 | 21 | 0.09 | 58 | 0.05 | 55 | 0.05 | 3 | 0.04 |  |  | ✓ |
| G24 Dystonia | 45 | 0.01 | 40 | 0.01 | 5 | 0.02 | 17 | 0.01 | 16 | 0.01 | 1 | 0.01 |  |  | ✓ |
| G25 Other extrapyramidal and movement disorders | 186 | 0.05 | 169 | 0.05 | 17 | 0.07 | 65 | 0.06 | 57 | 0.05 | 8 | 0.11 |  |  | ✓ |
| G30 Alzheimer disease | 7686 | 2.26 | 6881 | 2.17 | 805 | 3.48 | 2938 | 2.49 | 2626 | 2.38 | 312 | 4.16 |  | ✓ | ✓ |
| G31 Other degenerative diseases of nervous system, not elsewhere classified | 607 | 0.18 | 541 | 0.17 | 66 | 0.29 | 266 | 0.23 | 244 | 0.22 | 22 | 0.29 |  |  | ✓ |
| G35 Multiple sclerosis | 157 | 0.05 | 142 | 0.04 | 15 | 0.06 | 44 | 0.04 | 41 | 0.04 | 3 | 0.04 |  |  | ✓ |
| G36 Other acute disseminated demyelination | 34 | 0.01 | 34 | 0.01 | 0 | 0 | 17 | 0.01 | 16 | 0.01 | 1 | 0.01 |  |  | ✓ |
| G40 Epilepsy | 6868 | 2.02 | 6198 | 1.96 | 670 | 2.9 | 2355 | 1.99 | 2104 | 1.9 | 251 | 3.34 |  | ✓ | ✓ |
| G41 Status epilepticus | 261 | 0.08 | 230 | 0.07 | 31 | 0.13 | 136 | 0.12 | 123 | 0.11 | 13 | 0.17 |  |  | ✓ |
| G43 Migraine | 344 | 0.1 | 331 | 0.1 | 13 | 0.06 | 118 | 0.1 | 108 | 0.1 | 10 | 0.13 |  |  | ✓ |
| G44 Other headache syndromes | 231 | 0.07 | 217 | 0.07 | 14 | 0.06 | 73 | 0.06 | 67 | 0.06 | 6 | 0.08 |  |  | ✓ |
| G45 Transient cerebral ischaemic attacks and related syndromes | 1212 | 0.36 | 1157 | 0.37 | 55 | 0.24 | 420 | 0.36 | 403 | 0.36 | 17 | 0.23 |  |  | ✓ |
| G47 Sleep disorders | 24818 | 7.31 | 22969 | 7.26 | 1849 | 8 | 8578 | 7.26 | 7949 | 7.19 | 629 | 8.38 | ✓ | ✓ | ✓ |
| G50 Disorders of trigeminal nerve | 147 | 0.04 | 135 | 0.04 | 12 | 0.05 | 61 | 0.05 | 58 | 0.05 | 3 | 0.04 |  |  | ✓ |
| G51 Facial nerve disorders | 819 | 0.24 | 800 | 0.25 | 19 | 0.08 | 265 | 0.22 | 260 | 0.24 | 5 | 0.07 |  |  | ✓ |
| G52 Disorders of other cranial nerves | 388 | 0.11 | 351 | 0.11 | 37 | 0.16 | 135 | 0.11 | 124 | 0.11 | 11 | 0.15 |  |  | ✓ |
| G54 Nerve root and plexus disorders | 37 | 0.01 | 36 | 0.01 | 1 | 0 | 17 | 0.01 | 16 | 0.01 | 1 | 0.01 |  |  | ✓ |
| G56 Mononeuropathies of upper limb | 423 | 0.12 | 412 | 0.13 | 11 | 0.05 | 138 | 0.12 | 136 | 0.12 | 2 | 0.03 |  |  | ✓ |
| G57 Mononeuropathies of lower limb | 98 | 0.03 | 96 | 0.03 | 2 | 0.01 | 33 | 0.03 | 32 | 0.03 | 1 | 0.01 |  |  | ✓ |
| G58 Other mononeuropathies | 168 | 0.05 | 157 | 0.05 | 11 | 0.05 | 40 | 0.03 | 37 | 0.03 | 3 | 0.04 |  |  | ✓ |
| G61 Inflammatory polyneuropathy | 209 | 0.06 | 203 | 0.06 | 6 | 0.03 | 87 | 0.07 | 83 | 0.08 | 4 | 0.05 |  |  | ✓ |
| G62 Other polyneuropathies | 5243 | 1.54 | 4958 | 1.57 | 285 | 1.23 | 1689 | 1.43 | 1602 | 1.45 | 87 | 1.16 |  | ✓ | ✓ |
| G64 Other disorders of peripheral nervous system | 4895 | 1.44 | 4492 | 1.42 | 403 | 1.74 | 1494 | 1.27 | 1386 | 1.25 | 108 | 1.44 |  | ✓ | ✓ |
| G70 Myasthenia gravis and other myoneural disorders | 175 | 0.05 | 165 | 0.05 | 10 | 0.04 | 67 | 0.06 | 64 | 0.06 | 3 | 0.04 |  |  | ✓ |
| G71 Primary disorders of muscles | 134 | 0.04 | 119 | 0.04 | 15 | 0.06 | 43 | 0.04 | 37 | 0.03 | 6 | 0.08 |  |  | ✓ |
| G72 Other myopathies | 100 | 0.03 | 89 | 0.03 | 11 | 0.05 | 29 | 0.02 | 29 | 0.03 | 0 | 0 |  |  | ✓ |
| G80 Cerebral palsy | 378 | 0.11 | 322 | 0.1 | 56 | 0.24 | 110 | 0.09 | 92 | 0.08 | 18 | 0.24 |  |  | ✓ |
| G81 Hemiplegia | 1864 | 0.55 | 1765 | 0.56 | 99 | 0.43 | 485 | 0.41 | 459 | 0.42 | 26 | 0.35 |  |  | ✓ |
| G82 Paraplegia and tetraplegia | 122 | 0.04 | 113 | 0.04 | 9 | 0.04 | 43 | 0.04 | 40 | 0.04 | 3 | 0.04 |  |  | ✓ |
| G83 Other paralytic syndromes | 735 | 0.22 | 686 | 0.22 | 49 | 0.21 | 196 | 0.17 | 189 | 0.17 | 7 | 0.09 |  |  | ✓ |
| G90 Disorders of autonomic nervous system | 652 | 0.19 | 617 | 0.2 | 35 | 0.15 | 242 | 0.2 | 226 | 0.2 | 16 | 0.21 |  |  | ✓ |
| G91 Hydrocephalus | 571 | 0.17 | 522 | 0.16 | 49 | 0.21 | 212 | 0.18 | 196 | 0.18 | 16 | 0.21 |  |  | ✓ |
| G93 Other disorders of brain | 482 | 0.14 | 447 | 0.14 | 35 | 0.15 | 130 | 0.11 | 118 | 0.11 | 12 | 0.16 |  |  | ✓ |
| G95 Other diseases of spinal cord | 255 | 0.08 | 244 | 0.08 | 11 | 0.05 | 98 | 0.08 | 90 | 0.08 | 8 | 0.11 |  |  | ✓ |
| G96 Other disorders of central nervous system | 264 | 0.08 | 253 | 0.08 | 11 | 0.05 | 59 | 0.05 | 58 | 0.05 | 1 | 0.01 |  |  | ✓ |
| G97 Postprocedural disorders of nervous system, not elsewhere classified | 69 | 0.02 | 62 | 0.02 | 7 | 0.03 | 29 | 0.02 | 28 | 0.03 | 1 | 0.01 |  |  | ✓ |
| G98 Other disorders of nervous system, not elsewhere classified | 2110 | 0.62 | 1955 | 0.62 | 155 | 0.67 | 710 | 0.6 | 659 | 0.6 | 51 | 0.68 |  |  | ✓ |
| H00 Hordeolum and chalazion | 48 | 0.01 | 47 | 0.01 | 1 | 0 | 11 | 0.01 | 11 | 0.01 | 0 | 0 |  |  | ✓ |
| H01 Other inflammation of eyelid | 133 | 0.04 | 128 | 0.04 | 5 | 0.02 | 49 | 0.04 | 48 | 0.04 | 1 | 0.01 |  |  | ✓ |
| H02 Other disorders of eyelid | 83 | 0.02 | 79 | 0.02 | 4 | 0.02 | 22 | 0.02 | 22 | 0.02 | 0 | 0 |  |  | ✓ |
| H04 Disorders of lacrimal system | 666 | 0.2 | 624 | 0.2 | 42 | 0.18 | 230 | 0.19 | 215 | 0.19 | 15 | 0.2 |  |  | ✓ |
| H05 Disorders of orbit | 58 | 0.02 | 57 | 0.02 | 1 | 0 | 16 | 0.01 | 16 | 0.01 | 0 | 0 |  |  | ✓ |
| H10 Conjunctivitis | 776 | 0.23 | 723 | 0.23 | 53 | 0.23 | 246 | 0.21 | 232 | 0.21 | 14 | 0.19 |  |  | ✓ |
| H11 Other disorders of conjunctiva | 74 | 0.02 | 71 | 0.02 | 3 | 0.01 | 26 | 0.02 | 24 | 0.02 | 2 | 0.03 |  |  | ✓ |
| H16 Keratitis | 322 | 0.09 | 309 | 0.1 | 13 | 0.06 | 76 | 0.06 | 74 | 0.07 | 2 | 0.03 |  |  | ✓ |
| H20 Iridocyclitis | 280 | 0.08 | 264 | 0.08 | 16 | 0.07 | 100 | 0.08 | 95 | 0.09 | 5 | 0.07 |  |  | ✓ |
| H25 Senile cataract | 573 | 0.17 | 551 | 0.17 | 22 | 0.1 | 235 | 0.2 | 225 | 0.2 | 10 | 0.13 |  |  | ✓ |
| H26 Other cataract | 551 | 0.16 | 519 | 0.16 | 32 | 0.14 | 206 | 0.17 | 196 | 0.18 | 10 | 0.13 |  |  | ✓ |
| H33 Retinal detachments and breaks | 325 | 0.1 | 312 | 0.1 | 13 | 0.06 | 94 | 0.08 | 94 | 0.09 | 0 | 0 |  |  | ✓ |
| H34 Retinal vascular occlusions | 108 | 0.03 | 102 | 0.03 | 6 | 0.03 | 27 | 0.02 | 27 | 0.02 | 0 | 0 |  |  | ✓ |
| H35 Other retinal disorders | 285 | 0.08 | 273 | 0.09 | 12 | 0.05 | 90 | 0.08 | 81 | 0.07 | 9 | 0.12 |  |  | ✓ |
| H40 Glaucoma | 1123 | 0.33 | 1044 | 0.33 | 79 | 0.34 | 413 | 0.35 | 389 | 0.35 | 24 | 0.32 |  |  | ✓ |
| H43 Disorders of vitreous body | 158 | 0.05 | 145 | 0.05 | 13 | 0.06 | 49 | 0.04 | 48 | 0.04 | 1 | 0.01 |  |  | ✓ |
| H44 Disorders of globe | 58 | 0.02 | 54 | 0.02 | 4 | 0.02 | 17 | 0.01 | 15 | 0.01 | 2 | 0.03 |  |  | ✓ |
| H46 Optic neuritis | 54 | 0.02 | 52 | 0.02 | 2 | 0.01 | 35 | 0.03 | 33 | 0.03 | 2 | 0.03 |  |  | ✓ |
| H49 Paralytic strabismus | 101 | 0.03 | 98 | 0.03 | 3 | 0.01 | 43 | 0.04 | 43 | 0.04 | 0 | 0 |  |  | ✓ |
| H52 Disorders of refraction and accommodation | 716 | 0.21 | 686 | 0.22 | 30 | 0.13 | 185 | 0.16 | 176 | 0.16 | 9 | 0.12 |  |  | ✓ |
| H53 Visual disturbances | 211 | 0.06 | 203 | 0.06 | 8 | 0.03 | 55 | 0.05 | 54 | 0.05 | 1 | 0.01 |  |  | ✓ |
| H54 Visual impairment including blindness (binocular or monocular) | 49 | 0.01 | 47 | 0.01 | 2 | 0.01 | 15 | 0.01 | 15 | 0.01 | 0 | 0 |  |  | ✓ |
| H55 Nystagmus and other irregular eye movements | 68 | 0.02 | 64 | 0.02 | 4 | 0.02 | 24 | 0.02 | 24 | 0.02 | 0 | 0 |  |  | ✓ |
| H59 Postprocedural disorders of eye and adnexa, not elsewhere classified | 83 | 0.02 | 82 | 0.03 | 1 | 0 | 34 | 0.03 | 32 | 0.03 | 2 | 0.03 |  |  | ✓ |
| H60 Otitis externa | 142 | 0.04 | 141 | 0.04 | 1 | 0 | 51 | 0.04 | 48 | 0.04 | 3 | 0.04 |  |  | ✓ |
| H61 Other disorders of external ear | 389 | 0.11 | 371 | 0.12 | 18 | 0.08 | 153 | 0.13 | 139 | 0.13 | 14 | 0.19 |  |  | ✓ |
| H65 Nonsuppurative otitis media | 463 | 0.14 | 435 | 0.14 | 28 | 0.12 | 156 | 0.13 | 141 | 0.13 | 15 | 0.2 |  |  | ✓ |
| H66 Suppurative and unspecified otitis media | 1540 | 0.45 | 1453 | 0.46 | 87 | 0.38 | 578 | 0.49 | 539 | 0.49 | 39 | 0.52 |  |  | ✓ |
| H71 Cholesteatoma of middle ear | 158 | 0.05 | 154 | 0.05 | 4 | 0.02 | 56 | 0.05 | 55 | 0.05 | 1 | 0.01 |  |  | ✓ |
| H81 Disorders of vestibular function | 3353 | 0.99 | 3195 | 1.01 | 158 | 0.68 | 1327 | 1.12 | 1272 | 1.15 | 55 | 0.73 |  |  | ✓ |
| H83 Other diseases of inner ear | 46 | 0.01 | 44 | 0.01 | 2 | 0.01 | 17 | 0.01 | 17 | 0.02 | 0 | 0 |  |  | ✓ |
| H90 Conductive and sensorineural hearing loss | 366 | 0.11 | 357 | 0.11 | 9 | 0.04 | 128 | 0.11 | 123 | 0.11 | 5 | 0.07 |  |  | ✓ |
| H91 Other hearing loss | 920 | 0.27 | 893 | 0.28 | 27 | 0.12 | 260 | 0.22 | 254 | 0.23 | 6 | 0.08 |  |  | ✓ |
| H92 Otalgia and effusion of ear | 41 | 0.01 | 39 | 0.01 | 2 | 0.01 | 13 | 0.01 | 13 | 0.01 | 0 | 0 |  |  | ✓ |
| I05 Rheumatic mitral valve diseases | 222 | 0.07 | 202 | 0.06 | 20 | 0.09 | 64 | 0.05 | 59 | 0.05 | 5 | 0.07 |  |  | ✓ |
| I07 Rheumatic tricuspid valve diseases | 1327 | 0.39 | 1235 | 0.39 | 92 | 0.4 | 581 | 0.49 | 546 | 0.49 | 35 | 0.47 |  |  | ✓ |
| I08 Multiple valve diseases | 766 | 0.23 | 694 | 0.22 | 72 | 0.31 | 538 | 0.46 | 482 | 0.44 | 56 | 0.75 |  |  | ✓ |
| I10 Essential (primary) hypertension | 106140 | 31.26 | 98472 | 31.12 | 7668 | 33.18 | 38817 | 32.88 | 36235 | 32.77 | 2582 | 34.39 | ✓ | ✓ | ✓ |
| I11 Hypertensive heart disease | 1506 | 0.44 | 1394 | 0.44 | 112 | 0.48 | 580 | 0.49 | 539 | 0.49 | 41 | 0.55 |  |  | ✓ |
| I12 Hypertensive renal disease | 514 | 0.15 | 450 | 0.14 | 64 | 0.28 | 97 | 0.08 | 82 | 0.07 | 15 | 0.2 |  |  | ✓ |
| I15 Secondary hypertension | 128 | 0.04 | 119 | 0.04 | 9 | 0.04 | 34 | 0.03 | 31 | 0.03 | 3 | 0.04 |  |  | ✓ |
| I20 Angina pectoris | 27513 | 8.1 | 25743 | 8.14 | 1770 | 7.66 | 8943 | 7.57 | 8430 | 7.62 | 513 | 6.83 | ✓ | ✓ | ✓ |
| I21 Acute myocardial infarction | 3465 | 1.02 | 3303 | 1.04 | 162 | 0.7 | 1240 | 1.05 | 1190 | 1.08 | 50 | 0.67 |  | ✓ | ✓ |
| I23 Certain current complications following acute myocardial infarction | 21 | 0.01 | 21 | 0.01 | 0 | 0 | 16 | 0.01 | 15 | 0.01 | 1 | 0.01 |  |  | ✓ |
| I24 Other acute ischaemic heart diseases | 617 | 0.18 | 595 | 0.19 | 22 | 0.1 | 186 | 0.16 | 180 | 0.16 | 6 | 0.08 |  |  | ✓ |
| I25 Chronic ischaemic heart disease | 10194 | 3 | 9516 | 3.01 | 678 | 2.93 | 3919 | 3.32 | 3679 | 3.33 | 240 | 3.2 |  | ✓ | ✓ |
| I26 Pulmonary embolism | 1137 | 0.33 | 1052 | 0.33 | 85 | 0.37 | 462 | 0.39 | 423 | 0.38 | 39 | 0.52 |  |  | ✓ |
| I27 Other pulmonary heart diseases | 870 | 0.26 | 782 | 0.25 | 88 | 0.38 | 335 | 0.28 | 302 | 0.27 | 33 | 0.44 |  |  | ✓ |
| I28 Other diseases of pulmonary vessels | 29 | 0.01 | 28 | 0.01 | 1 | 0 | 11 | 0.01 | 11 | 0.01 | 0 | 0 |  |  | ✓ |
| I30 Acute pericarditis | 56 | 0.02 | 55 | 0.02 | 1 | 0 | 32 | 0.03 | 30 | 0.03 | 2 | 0.03 |  |  | ✓ |
| I31 Other diseases of pericardium | 505 | 0.15 | 449 | 0.14 | 56 | 0.24 | 175 | 0.15 | 163 | 0.15 | 12 | 0.16 |  |  | ✓ |
| I33 Acute and subacute endocarditis | 195 | 0.06 | 169 | 0.05 | 26 | 0.11 | 56 | 0.05 | 49 | 0.04 | 7 | 0.09 |  |  | ✓ |
| I34 Nonrheumatic mitral valve disorders | 3431 | 1.01 | 3187 | 1.01 | 244 | 1.06 | 1034 | 0.88 | 956 | 0.86 | 78 | 1.04 |  |  | ✓ |
| I35 Nonrheumatic aortic valve disorders | 4700 | 1.38 | 4296 | 1.36 | 404 | 1.75 | 1864 | 1.58 | 1745 | 1.58 | 119 | 1.59 |  | ✓ | ✓ |
| I37 Pulmonary valve disorders | 31 | 0.01 | 30 | 0.01 | 1 | 0 | 15 | 0.01 | 15 | 0.01 | 0 | 0 |  |  | ✓ |
| I38 Endocarditis, valve unspecified | 467 | 0.14 | 434 | 0.14 | 33 | 0.14 | 145 | 0.12 | 129 | 0.12 | 16 | 0.21 |  |  | ✓ |
| I40 Acute myocarditis | 44 | 0.01 | 41 | 0.01 | 3 | 0.01 | 13 | 0.01 | 13 | 0.01 | 0 | 0 |  |  | ✓ |
| I42 Cardiomyopathy | 1513 | 0.45 | 1363 | 0.43 | 150 | 0.65 | 518 | 0.44 | 480 | 0.43 | 38 | 0.51 |  |  | ✓ |
| I44 Atrioventricular and left bundle-branch block | 3166 | 0.93 | 2992 | 0.95 | 174 | 0.75 | 1197 | 1.01 | 1128 | 1.02 | 69 | 0.92 |  |  | ✓ |
| I45 Other conduction disorders | 706 | 0.21 | 666 | 0.21 | 40 | 0.17 | 258 | 0.22 | 245 | 0.22 | 13 | 0.17 |  |  | ✓ |
| I46 Cardiac arrest | 219 | 0.06 | 188 | 0.06 | 31 | 0.13 | 76 | 0.06 | 72 | 0.07 | 4 | 0.05 |  |  | ✓ |
| I47 Paroxysmal tachycardia | 3709 | 1.09 | 3499 | 1.11 | 210 | 0.91 | 1729 | 1.46 | 1662 | 1.5 | 67 | 0.89 |  | ✓ | ✓ |
| I48 Atrial fibrillation and flutter | 22749 | 6.7 | 20767 | 6.56 | 1982 | 8.58 | 8763 | 7.42 | 8004 | 7.24 | 759 | 10.11 | ✓ | ✓ | ✓ |
| I49 Other cardiac arrhythmias | 5685 | 1.67 | 5376 | 1.7 | 309 | 1.34 | 1800 | 1.52 | 1697 | 1.53 | 103 | 1.37 |  | ✓ | ✓ |
| I50 Heart failure | 33468 | 9.86 | 29941 | 9.46 | 3527 | 15.26 | 12191 | 10.32 | 10968 | 9.92 | 1223 | 16.29 | ✓ | ✓ | ✓ |
| I51 Complications and ill-defined descriptions of heart disease | 1650 | 0.49 | 1577 | 0.5 | 73 | 0.32 | 740 | 0.63 | 712 | 0.64 | 28 | 0.37 |  |  | ✓ |
| I60 Subarachnoid haemorrhage | 557 | 0.16 | 544 | 0.17 | 13 | 0.06 | 196 | 0.17 | 192 | 0.17 | 4 | 0.05 |  |  | ✓ |
| I61 Intracerebral haemorrhage | 1378 | 0.41 | 1308 | 0.41 | 70 | 0.3 | 447 | 0.38 | 424 | 0.38 | 23 | 0.31 |  |  | ✓ |
| I62 Other nontraumatic intracranial haemorrhage | 521 | 0.15 | 463 | 0.15 | 58 | 0.25 | 195 | 0.17 | 185 | 0.17 | 10 | 0.13 |  |  | ✓ |
| I63 Cerebral infarction | 9774 | 2.88 | 9176 | 2.9 | 598 | 2.59 | 3282 | 2.78 | 3066 | 2.77 | 216 | 2.88 |  | ✓ | ✓ |
| I65 Occlusion and stenosis of precerebral arteries, not resulting in cerebral infarction | 2013 | 0.59 | 1920 | 0.61 | 93 | 0.4 | 677 | 0.57 | 650 | 0.59 | 27 | 0.36 |  |  | ✓ |
| I66 Occlusion and stenosis of cerebral arteries, not resulting in cerebral infarction | 722 | 0.21 | 689 | 0.22 | 33 | 0.14 | 178 | 0.15 | 170 | 0.15 | 8 | 0.11 |  |  | ✓ |
| I67 Other cerebrovascular diseases | 1323 | 0.39 | 1272 | 0.4 | 51 | 0.22 | 487 | 0.41 | 472 | 0.43 | 15 | 0.2 |  |  | ✓ |
| I69 Sequelae of cerebrovascular disease | 15287 | 4.5 | 13820 | 4.37 | 1467 | 6.35 | 5197 | 4.4 | 4719 | 4.27 | 478 | 6.37 |  | ✓ | ✓ |
| I70 Atherosclerosis | 5313 | 1.56 | 4981 | 1.57 | 332 | 1.44 | 1915 | 1.62 | 1796 | 1.62 | 119 | 1.59 |  | ✓ | ✓ |
| I71 Aortic aneurysm and dissection | 3625 | 1.07 | 3392 | 1.07 | 233 | 1.01 | 1429 | 1.21 | 1352 | 1.22 | 77 | 1.03 |  | ✓ | ✓ |
| I72 Other aneurysm and dissection | 924 | 0.27 | 877 | 0.28 | 47 | 0.2 | 376 | 0.32 | 356 | 0.32 | 20 | 0.27 |  |  | ✓ |
| I73 Other peripheral vascular diseases | 486 | 0.14 | 470 | 0.15 | 16 | 0.07 | 168 | 0.14 | 159 | 0.14 | 9 | 0.12 |  |  | ✓ |
| I74 Arterial embolism and thrombosis | 2609 | 0.77 | 2438 | 0.77 | 171 | 0.74 | 855 | 0.72 | 791 | 0.72 | 64 | 0.85 |  |  | ✓ |
| I77 Other disorders of arteries and arterioles | 252 | 0.07 | 237 | 0.07 | 15 | 0.06 | 100 | 0.08 | 94 | 0.09 | 6 | 0.08 |  |  | ✓ |
| I78 Diseases of capillaries | 69 | 0.02 | 55 | 0.02 | 14 | 0.06 | 25 | 0.02 | 22 | 0.02 | 3 | 0.04 |  |  | ✓ |
| I80 Phlebitis and thrombophlebitis | 4163 | 1.23 | 3860 | 1.22 | 303 | 1.31 | 1774 | 1.5 | 1655 | 1.5 | 119 | 1.59 |  | ✓ | ✓ |
| I81 Portal vein thrombosis | 206 | 0.06 | 179 | 0.06 | 27 | 0.12 | 78 | 0.07 | 67 | 0.06 | 11 | 0.15 |  |  | ✓ |
| I82 Other venous embolism and thrombosis | 347 | 0.1 | 316 | 0.1 | 31 | 0.13 | 129 | 0.11 | 119 | 0.11 | 10 | 0.13 |  |  | ✓ |
| I83 Varicose veins of lower extremities | 345 | 0.1 | 322 | 0.1 | 23 | 0.1 | 137 | 0.12 | 132 | 0.12 | 5 | 0.07 |  |  | ✓ |
| I85 Oesophageal varices | 1277 | 0.38 | 1116 | 0.35 | 161 | 0.7 | 353 | 0.3 | 314 | 0.28 | 39 | 0.52 |  |  | ✓ |
| I86 Varicose veins of other sites | 221 | 0.07 | 193 | 0.06 | 28 | 0.12 | 73 | 0.06 | 63 | 0.06 | 10 | 0.13 |  |  | ✓ |
| I87 Other disorders of veins | 106 | 0.03 | 87 | 0.03 | 19 | 0.08 | 32 | 0.03 | 30 | 0.03 | 2 | 0.03 |  |  | ✓ |
| I88 Nonspecific lymphadenitis | 172 | 0.05 | 168 | 0.05 | 4 | 0.02 | 57 | 0.05 | 57 | 0.05 | 0 | 0 |  |  | ✓ |
| I89 Other noninfective disorders of lymphatic vessels and lymph nodes | 249 | 0.07 | 224 | 0.07 | 25 | 0.11 | 70 | 0.06 | 65 | 0.06 | 5 | 0.07 |  |  | ✓ |
| I95 Hypotension | 2249 | 0.66 | 2141 | 0.68 | 108 | 0.47 | 652 | 0.55 | 625 | 0.57 | 27 | 0.36 |  |  | ✓ |
| J00 Acute nasopharyngitis [common cold] | 529 | 0.16 | 501 | 0.16 | 28 | 0.12 | 148 | 0.13 | 141 | 0.13 | 7 | 0.09 |  |  | ✓ |
| J01 Acute sinusitis | 368 | 0.11 | 349 | 0.11 | 19 | 0.08 | 118 | 0.1 | 112 | 0.1 | 6 | 0.08 |  |  | ✓ |
| J02 Acute pharyngitis | 4643 | 1.37 | 4393 | 1.39 | 250 | 1.08 | 1692 | 1.43 | 1604 | 1.45 | 88 | 1.17 |  | ✓ | ✓ |
| J03 Acute tonsillitis | 1539 | 0.45 | 1482 | 0.47 | 57 | 0.25 | 518 | 0.44 | 491 | 0.44 | 27 | 0.36 |  |  | ✓ |
| J04 Acute laryngitis and tracheitis | 506 | 0.15 | 475 | 0.15 | 31 | 0.13 | 158 | 0.13 | 151 | 0.14 | 7 | 0.09 |  |  | ✓ |
| J05 Acute obstructive laryngitis [croup] and epiglottitis | 476 | 0.14 | 457 | 0.14 | 19 | 0.08 | 168 | 0.14 | 164 | 0.15 | 4 | 0.05 |  |  | ✓ |
| J06 Acute upper respiratory infections of multiple and unspecified sites | 5492 | 1.62 | 5186 | 1.64 | 306 | 1.32 | 1782 | 1.51 | 1682 | 1.52 | 100 | 1.33 |  | ✓ | ✓ |
| J10 Influenza due to identified seasonal influenza virus | 2385 | 0.7 | 2247 | 0.71 | 138 | 0.6 | 1020 | 0.86 | 965 | 0.87 | 55 | 0.73 |  |  | ✓ |
| J11 Influenza, virus not identified | 673 | 0.2 | 628 | 0.2 | 45 | 0.19 | 226 | 0.19 | 214 | 0.19 | 12 | 0.16 |  |  | ✓ |
| J12 Viral pneumonia, not elsewhere classified | 1806 | 0.53 | 1693 | 0.54 | 113 | 0.49 | 673 | 0.57 | 629 | 0.57 | 44 | 0.59 |  |  | ✓ |
| J13 Pneumonia due to Streptococcus pneumoniae | 1215 | 0.36 | 1110 | 0.35 | 105 | 0.45 | 431 | 0.37 | 394 | 0.36 | 37 | 0.49 |  |  | ✓ |
| J14 Pneumonia due to Haemophilus influenzae | 484 | 0.14 | 428 | 0.14 | 56 | 0.24 | 122 | 0.1 | 107 | 0.1 | 15 | 0.2 |  |  | ✓ |
| J15 Bacterial pneumonia, not elsewhere classified | 8879 | 2.62 | 8003 | 2.53 | 876 | 3.79 | 2687 | 2.28 | 2429 | 2.2 | 258 | 3.44 |  | ✓ | ✓ |
| J18 Pneumonia, organism unspecified | 12087 | 3.56 | 10812 | 3.42 | 1275 | 5.52 | 4100 | 3.47 | 3685 | 3.33 | 415 | 5.53 |  | ✓ | ✓ |
| J20 Acute bronchitis | 11042 | 3.25 | 10310 | 3.26 | 732 | 3.17 | 3978 | 3.37 | 3728 | 3.37 | 250 | 3.33 |  | ✓ | ✓ |
| J21 Acute bronchiolitis | 1234 | 0.36 | 1187 | 0.38 | 47 | 0.2 | 378 | 0.32 | 359 | 0.32 | 19 | 0.25 |  |  | ✓ |
| J30 Vasomotor and allergic rhinitis | 4773 | 1.41 | 4487 | 1.42 | 286 | 1.24 | 1762 | 1.49 | 1644 | 1.49 | 118 | 1.57 |  | ✓ | ✓ |
| J31 Chronic rhinitis, nasopharyngitis and pharyngitis | 275 | 0.08 | 260 | 0.08 | 15 | 0.06 | 61 | 0.05 | 58 | 0.05 | 3 | 0.04 |  |  | ✓ |
| J32 Chronic sinusitis | 1093 | 0.32 | 1029 | 0.33 | 64 | 0.28 | 388 | 0.33 | 365 | 0.33 | 23 | 0.31 |  |  | ✓ |
| J33 Nasal polyp | 52 | 0.02 | 50 | 0.02 | 2 | 0.01 | 16 | 0.01 | 16 | 0.01 | 0 | 0 |  |  | ✓ |
| J34 Other disorders of nose and nasal sinuses | 247 | 0.07 | 238 | 0.08 | 9 | 0.04 | 63 | 0.05 | 62 | 0.06 | 1 | 0.01 |  |  | ✓ |
| J35 Chronic diseases of tonsils and adenoids | 613 | 0.18 | 578 | 0.18 | 35 | 0.15 | 168 | 0.14 | 157 | 0.14 | 11 | 0.15 |  |  | ✓ |
| J36 Peritonsillar abscess | 978 | 0.29 | 951 | 0.3 | 27 | 0.12 | 304 | 0.26 | 293 | 0.26 | 11 | 0.15 |  |  | ✓ |
| J37 Chronic laryngitis and laryngotracheitis | 472 | 0.14 | 443 | 0.14 | 29 | 0.13 | 109 | 0.09 | 100 | 0.09 | 9 | 0.12 |  |  | ✓ |
| J38 Diseases of vocal cords and larynx, not elsewhere classified | 1106 | 0.33 | 1069 | 0.34 | 37 | 0.16 | 370 | 0.31 | 358 | 0.32 | 12 | 0.16 |  |  | ✓ |
| J39 Other diseases of upper respiratory tract | 263 | 0.08 | 245 | 0.08 | 18 | 0.08 | 73 | 0.06 | 69 | 0.06 | 4 | 0.05 |  |  | ✓ |
| J40 Bronchitis, not specified as acute or chronic | 1196 | 0.35 | 1087 | 0.34 | 109 | 0.47 | 397 | 0.34 | 358 | 0.32 | 39 | 0.52 |  |  | ✓ |
| J42 Unspecified chronic bronchitis | 3564 | 1.05 | 3152 | 1 | 412 | 1.78 | 858 | 0.73 | 771 | 0.7 | 87 | 1.16 |  |  | ✓ |
| J43 Emphysema | 2670 | 0.79 | 2426 | 0.77 | 244 | 1.06 | 909 | 0.77 | 825 | 0.75 | 84 | 1.12 |  |  | ✓ |
| J44 Other chronic obstructive pulmonary disease | 7016 | 2.07 | 6030 | 1.91 | 986 | 4.27 | 2458 | 2.08 | 2201 | 1.99 | 257 | 3.42 |  | ✓ | ✓ |
| J45 Asthma | 17785 | 5.24 | 16373 | 5.17 | 1412 | 6.11 | 6189 | 5.24 | 5733 | 5.19 | 456 | 6.07 | ✓ | ✓ | ✓ |
| J46 Status asthmaticus | 3742 | 1.1 | 3492 | 1.1 | 250 | 1.08 | 1018 | 0.86 | 936 | 0.85 | 82 | 1.09 |  |  | ✓ |
| J47 Bronchiectasis | 698 | 0.21 | 606 | 0.19 | 92 | 0.4 | 291 | 0.25 | 254 | 0.23 | 37 | 0.49 |  |  | ✓ |
| J64 Unspecified pneumoconiosis | 81 | 0.02 | 71 | 0.02 | 10 | 0.04 | 24 | 0.02 | 22 | 0.02 | 2 | 0.03 |  |  | ✓ |
| J67 Hypersensitivity pneumonitis due to organic dust | 79 | 0.02 | 73 | 0.02 | 6 | 0.03 | 33 | 0.03 | 28 | 0.03 | 5 | 0.07 |  |  | ✓ |
| J69 Pneumonitis due to solids and liquids | 8557 | 2.52 | 7210 | 2.28 | 1347 | 5.83 | 2784 | 2.36 | 2382 | 2.15 | 402 | 5.36 |  | ✓ | ✓ |
| J70 Respiratory conditions due to other external agents | 415 | 0.12 | 359 | 0.11 | 56 | 0.24 | 206 | 0.17 | 174 | 0.16 | 32 | 0.43 |  |  | ✓ |
| J80 Adult respiratory distress syndrome | 117 | 0.03 | 106 | 0.03 | 11 | 0.05 | 35 | 0.03 | 31 | 0.03 | 4 | 0.05 |  |  | ✓ |
| J81 Pulmonary oedema | 457 | 0.13 | 406 | 0.13 | 51 | 0.22 | 172 | 0.15 | 153 | 0.14 | 19 | 0.25 |  |  | ✓ |
| J82 Pulmonary eosinophilia, not elsewhere classified | 129 | 0.04 | 117 | 0.04 | 12 | 0.05 | 41 | 0.03 | 38 | 0.03 | 3 | 0.04 |  |  | ✓ |
| J84 Other interstitial pulmonary diseases | 3587 | 1.06 | 3064 | 0.97 | 523 | 2.26 | 1295 | 1.1 | 1132 | 1.02 | 163 | 2.17 |  | ✓ | ✓ |
| J85 Abscess of lung and mediastinum | 435 | 0.13 | 398 | 0.13 | 37 | 0.16 | 180 | 0.15 | 167 | 0.15 | 13 | 0.17 |  |  | ✓ |
| J86 Pyothorax | 492 | 0.14 | 444 | 0.14 | 48 | 0.21 | 213 | 0.18 | 185 | 0.17 | 28 | 0.37 |  |  | ✓ |
| J90 Pleural effusion, not elsewhere classified | 7470 | 2.2 | 6394 | 2.02 | 1076 | 4.66 | 2864 | 2.43 | 2484 | 2.25 | 380 | 5.06 |  | ✓ | ✓ |
| J93 Pneumothorax | 1687 | 0.5 | 1512 | 0.48 | 175 | 0.76 | 578 | 0.49 | 521 | 0.47 | 57 | 0.76 |  |  | ✓ |
| J94 Other pleural conditions | 91 | 0.03 | 84 | 0.03 | 7 | 0.03 | 31 | 0.03 | 28 | 0.03 | 3 | 0.04 |  |  | ✓ |
| J95 Postprocedural respiratory disorders, not elsewhere classified | 398 | 0.12 | 365 | 0.12 | 33 | 0.14 | 114 | 0.1 | 103 | 0.09 | 11 | 0.15 |  |  | ✓ |
| J96 Respiratory failure, not elsewhere classified | 15685 | 4.62 | 13846 | 4.38 | 1839 | 7.96 | 4482 | 3.8 | 3991 | 3.61 | 491 | 6.54 |  | ✓ | ✓ |
| J98 Other respiratory disorders | 1293 | 0.38 | 1168 | 0.37 | 125 | 0.54 | 456 | 0.39 | 423 | 0.38 | 33 | 0.44 |  |  | ✓ |
| K04 Diseases of pulp and periapical tissues | 34 | 0.01 | 32 | 0.01 | 2 | 0.01 | 14 | 0.01 | 13 | 0.01 | 1 | 0.01 |  |  | ✓ |
| K05 Gingivitis and periodontal diseases | 132 | 0.04 | 127 | 0.04 | 5 | 0.02 | 109 | 0.09 | 103 | 0.09 | 6 | 0.08 |  |  | ✓ |
| K07 Dentofacial anomalies [including malocclusion] | 30 | 0.01 | 29 | 0.01 | 1 | 0 | 14 | 0.01 | 14 | 0.01 | 0 | 0 |  |  | ✓ |
| K10 Other diseases of jaws | 64 | 0.02 | 56 | 0.02 | 8 | 0.03 | 26 | 0.02 | 23 | 0.02 | 3 | 0.04 |  |  | ✓ |
| K11 Diseases of salivary glands | 312 | 0.09 | 299 | 0.09 | 13 | 0.06 | 86 | 0.07 | 84 | 0.08 | 2 | 0.03 |  |  | ✓ |
| K12 Stomatitis and related lesions | 4282 | 1.26 | 3944 | 1.25 | 338 | 1.46 | 1336 | 1.13 | 1253 | 1.13 | 83 | 1.11 |  | ✓ | ✓ |
| K13 Other diseases of lip and oral mucosa | 93 | 0.03 | 78 | 0.02 | 15 | 0.06 | 36 | 0.03 | 32 | 0.03 | 4 | 0.05 |  |  | ✓ |
| K14 Diseases of tongue | 30 | 0.01 | 26 | 0.01 | 4 | 0.02 | 12 | 0.01 | 12 | 0.01 | 0 | 0 |  |  | ✓ |
| K20 Oesophagitis | 268 | 0.08 | 245 | 0.08 | 23 | 0.1 | 68 | 0.06 | 62 | 0.06 | 6 | 0.08 |  |  | ✓ |
| K21 Gastro-oesophageal reflux disease | 47087 | 13.87 | 42952 | 13.58 | 4135 | 17.89 | 16680 | 14.13 | 15337 | 13.87 | 1343 | 17.89 | ✓ | ✓ | ✓ |
| K22 Other diseases of oesophagus | 1464 | 0.43 | 1311 | 0.41 | 153 | 0.66 | 490 | 0.41 | 457 | 0.41 | 33 | 0.44 |  |  | ✓ |
| K25 Gastric ulcer | 27943 | 8.23 | 25996 | 8.22 | 1947 | 8.43 | 9649 | 8.17 | 9050 | 8.19 | 599 | 7.98 | ✓ | ✓ | ✓ |
| K26 Duodenal ulcer | 2336 | 0.69 | 2164 | 0.68 | 172 | 0.74 | 754 | 0.64 | 708 | 0.64 | 46 | 0.61 |  |  | ✓ |
| K27 Peptic ulcer, site unspecified | 287 | 0.08 | 257 | 0.08 | 30 | 0.13 | 106 | 0.09 | 100 | 0.09 | 6 | 0.08 |  |  | ✓ |
| K28 Gastrojejunal ulcer | 176 | 0.05 | 158 | 0.05 | 18 | 0.08 | 32 | 0.03 | 31 | 0.03 | 1 | 0.01 |  |  | ✓ |
| K29 Gastritis and duodenitis | 22129 | 6.52 | 20684 | 6.54 | 1445 | 6.25 | 6022 | 5.1 | 5697 | 5.15 | 325 | 4.33 | ✓ | ✓ | ✓ |
| K30 Functional dyspepsia | 169 | 0.05 | 145 | 0.05 | 24 | 0.1 | 58 | 0.05 | 51 | 0.05 | 7 | 0.09 |  |  | ✓ |
| K31 Other diseases of stomach and duodenum | 2255 | 0.66 | 1982 | 0.63 | 273 | 1.18 | 743 | 0.63 | 672 | 0.61 | 71 | 0.95 |  |  | ✓ |
| K35Acute appendicitis | 4499 | 1.33 | 4374 | 1.38 | 125 | 0.54 | 1451 | 1.23 | 1421 | 1.29 | 30 | 0.4 |  | ✓ | ✓ |
| K36 Other appendicitis | 179 | 0.05 | 175 | 0.06 | 4 | 0.02 | 72 | 0.06 | 71 | 0.06 | 1 | 0.01 |  |  | ✓ |
| K37 Unspecified appendicitis | 146 | 0.04 | 138 | 0.04 | 8 | 0.03 | 43 | 0.04 | 40 | 0.04 | 3 | 0.04 |  |  | ✓ |
| K38 Other diseases of appendix | 69 | 0.02 | 66 | 0.02 | 3 | 0.01 | 30 | 0.03 | 30 | 0.03 | 0 | 0 |  |  | ✓ |
| K40 Inguinal hernia | 535 | 0.16 | 505 | 0.16 | 30 | 0.13 | 493 | 0.42 | 482 | 0.44 | 11 | 0.15 |  |  | ✓ |
| K41 Femoral hernia | 201 | 0.06 | 192 | 0.06 | 9 | 0.04 | 62 | 0.05 | 59 | 0.05 | 3 | 0.04 |  |  | ✓ |
| K42 Umbilical hernia | 149 | 0.04 | 137 | 0.04 | 12 | 0.05 | 56 | 0.05 | 53 | 0.05 | 3 | 0.04 |  |  | ✓ |
| K43 Ventral hernia | 613 | 0.18 | 579 | 0.18 | 34 | 0.15 | 226 | 0.19 | 215 | 0.19 | 11 | 0.15 |  |  | ✓ |
| K44 Diaphragmatic hernia | 1222 | 0.36 | 1120 | 0.35 | 102 | 0.44 | 402 | 0.34 | 375 | 0.34 | 27 | 0.36 |  |  | ✓ |
| K45 Other abdominal hernia | 124 | 0.04 | 113 | 0.04 | 11 | 0.05 | 54 | 0.05 | 49 | 0.04 | 5 | 0.07 |  |  | ✓ |
| K46 Unspecified abdominal hernia | 68 | 0.02 | 64 | 0.02 | 4 | 0.02 | 25 | 0.02 | 25 | 0.02 | 0 | 0 |  |  | ✓ |
| K50 Crohn disease [regional enteritis] | 325 | 0.1 | 298 | 0.09 | 27 | 0.12 | 94 | 0.08 | 87 | 0.08 | 7 | 0.09 |  |  | ✓ |
| K51 Ulcerative colitis | 853 | 0.25 | 800 | 0.25 | 53 | 0.23 | 311 | 0.26 | 291 | 0.26 | 20 | 0.27 |  |  | ✓ |
| K52 Other noninfective gastroenteritis and colitis | 1434 | 0.42 | 1319 | 0.42 | 115 | 0.5 | 330 | 0.28 | 309 | 0.28 | 21 | 0.28 |  |  | ✓ |
| K55 Vascular disorders of intestine | 2005 | 0.59 | 1916 | 0.61 | 89 | 0.39 | 825 | 0.7 | 789 | 0.71 | 36 | 0.48 |  |  | ✓ |
| K56 Paralytic ileus and intestinal obstruction without hernia | 6893 | 2.03 | 6114 | 1.93 | 779 | 3.37 | 2377 | 2.01 | 2131 | 1.93 | 246 | 3.28 |  | ✓ | ✓ |
| K57 Diverticular disease of intestine | 5184 | 1.53 | 4839 | 1.53 | 345 | 1.49 | 1879 | 1.59 | 1762 | 1.59 | 117 | 1.56 |  | ✓ | ✓ |
| K58 Irritable bowel syndrome | 1098 | 0.32 | 1020 | 0.32 | 78 | 0.34 | 352 | 0.3 | 326 | 0.29 | 26 | 0.35 |  |  | ✓ |
| K59 Other functional intestinal disorders | 45752 | 13.48 | 42324 | 13.38 | 3428 | 14.83 | 15432 | 13.07 | 14326 | 12.96 | 1106 | 14.73 | ✓ | ✓ | ✓ |
| K60 Fissure and fistula of anal and rectal regions | 183 | 0.05 | 173 | 0.05 | 10 | 0.04 | 60 | 0.05 | 57 | 0.05 | 3 | 0.04 |  |  | ✓ |
| K61 Abscess of anal and rectal regions | 237 | 0.07 | 229 | 0.07 | 8 | 0.03 | 73 | 0.06 | 68 | 0.06 | 5 | 0.07 |  |  | ✓ |
| K62 Other diseases of anus and rectum | 953 | 0.28 | 861 | 0.27 | 92 | 0.4 | 338 | 0.29 | 303 | 0.27 | 35 | 0.47 |  |  | ✓ |
| K63 Other diseases of intestine | 2475 | 0.73 | 2336 | 0.74 | 139 | 0.6 | 1224 | 1.04 | 1158 | 1.05 | 66 | 0.88 |  |  | ✓ |
| K64 Haemorrhoids and perianal venous thrombosis | 1889 | 0.56 | 1715 | 0.54 | 174 | 0.75 | 594 | 0.5 | 546 | 0.49 | 48 | 0.64 |  |  | ✓ |
| K65 Peritonitis | 3419 | 1.01 | 3073 | 0.97 | 346 | 1.5 | 1213 | 1.03 | 1096 | 0.99 | 117 | 1.56 |  | ✓ | ✓ |
| K66 Other disorders of peritoneum | 445 | 0.13 | 417 | 0.13 | 28 | 0.12 | 135 | 0.11 | 120 | 0.11 | 15 | 0.2 |  |  | ✓ |
| K70 Alcoholic liver disease | 2968 | 0.87 | 2613 | 0.83 | 355 | 1.54 | 986 | 0.84 | 890 | 0.8 | 96 | 1.28 |  |  | ✓ |
| K71 Toxic liver disease | 522 | 0.15 | 496 | 0.16 | 26 | 0.11 | 190 | 0.16 | 171 | 0.15 | 19 | 0.25 |  |  | ✓ |
| K72 Hepatic failure, not elsewhere classified | 1487 | 0.44 | 1097 | 0.35 | 390 | 1.69 | 456 | 0.39 | 361 | 0.33 | 95 | 1.27 |  |  | ✓ |
| K73 Chronic hepatitis, not elsewhere classified | 696 | 0.2 | 627 | 0.2 | 69 | 0.3 | 139 | 0.12 | 129 | 0.12 | 10 | 0.13 |  |  | ✓ |
| K74 Fibrosis and cirrhosis of liver | 3194 | 0.94 | 2730 | 0.86 | 464 | 2.01 | 957 | 0.81 | 832 | 0.75 | 125 | 1.67 |  |  | ✓ |
| K75 Other inflammatory liver diseases | 1134 | 0.33 | 998 | 0.32 | 136 | 0.59 | 402 | 0.34 | 354 | 0.32 | 48 | 0.64 |  |  | ✓ |
| K76 Other diseases of liver | 5951 | 1.75 | 5477 | 1.73 | 474 | 2.05 | 1957 | 1.66 | 1828 | 1.65 | 129 | 1.72 |  | ✓ | ✓ |
| K80 Cholelithiasis | 15334 | 4.52 | 14394 | 4.55 | 940 | 4.07 | 5376 | 4.55 | 5040 | 4.56 | 336 | 4.48 |  | ✓ | ✓ |
| K81 Cholecystitis | 2787 | 0.82 | 2522 | 0.8 | 265 | 1.15 | 913 | 0.77 | 845 | 0.76 | 68 | 0.91 |  |  | ✓ |
| K82 Other diseases of gallbladder | 516 | 0.15 | 502 | 0.16 | 14 | 0.06 | 195 | 0.17 | 190 | 0.17 | 5 | 0.07 |  |  | ✓ |
| K83 Other diseases of biliary tract | 8826 | 2.6 | 7230 | 2.29 | 1596 | 6.91 | 2984 | 2.53 | 2447 | 2.21 | 537 | 7.15 |  | ✓ | ✓ |
| K85 Acute pancreatitis | 5291 | 1.56 | 4807 | 1.52 | 484 | 2.09 | 1759 | 1.49 | 1591 | 1.44 | 168 | 2.24 |  | ✓ | ✓ |
| K86 Other diseases of pancreas | 3507 | 1.03 | 3096 | 0.98 | 411 | 1.78 | 1423 | 1.21 | 1278 | 1.16 | 145 | 1.93 |  | ✓ | ✓ |
| K90 Intestinal malabsorption | 56 | 0.02 | 43 | 0.01 | 13 | 0.06 | 13 | 0.01 | 11 | 0.01 | 2 | 0.03 |  |  | ✓ |
| K91 Postprocedural disorders of digestive system, not elsewhere classified | 3358 | 0.99 | 2979 | 0.94 | 379 | 1.64 | 1096 | 0.93 | 983 | 0.89 | 113 | 1.51 |  |  | ✓ |
| K92 Other diseases of digestive system | 5857 | 1.73 | 5336 | 1.69 | 521 | 2.25 | 1946 | 1.65 | 1784 | 1.61 | 162 | 2.16 |  | ✓ | ✓ |
| L00 Staphylococcal scalded skin syndrome | 37 | 0.01 | 34 | 0.01 | 3 | 0.01 | 10 | 0.01 | 10 | 0.01 | 0 | 0 |  |  | ✓ |
| L01 Impetigo | 184 | 0.05 | 171 | 0.05 | 13 | 0.06 | 52 | 0.04 | 52 | 0.05 | 0 | 0 |  |  | ✓ |
| L02 Cutaneous abscess, furuncle and carbuncle | 683 | 0.2 | 645 | 0.2 | 38 | 0.16 | 208 | 0.18 | 197 | 0.18 | 11 | 0.15 |  |  | ✓ |
| L03 Cellulitis | 3279 | 0.97 | 2990 | 0.94 | 289 | 1.25 | 1193 | 1.01 | 1095 | 0.99 | 98 | 1.31 |  |  | ✓ |
| L04 Acute lymphadenitis | 508 | 0.15 | 490 | 0.15 | 18 | 0.08 | 177 | 0.15 | 172 | 0.16 | 5 | 0.07 |  |  | ✓ |
| L08 Other local infections of skin and subcutaneous tissue | 746 | 0.22 | 710 | 0.22 | 36 | 0.16 | 240 | 0.2 | 228 | 0.21 | 12 | 0.16 |  |  | ✓ |
| L12 Pemphigoid | 169 | 0.05 | 154 | 0.05 | 15 | 0.06 | 63 | 0.05 | 53 | 0.05 | 10 | 0.13 |  |  | ✓ |
| L13 Other bullous disorders | 36 | 0.01 | 31 | 0.01 | 5 | 0.02 | 12 | 0.01 | 11 | 0.01 | 1 | 0.01 |  |  | ✓ |
| L20 Atopic dermatitis | 993 | 0.29 | 933 | 0.29 | 60 | 0.26 | 357 | 0.3 | 328 | 0.3 | 29 | 0.39 |  |  | ✓ |
| L21 Seborrhoeic dermatitis | 173 | 0.05 | 159 | 0.05 | 14 | 0.06 | 60 | 0.05 | 56 | 0.05 | 4 | 0.05 |  |  | ✓ |
| L22 Diaper [napkin] dermatitis | 768 | 0.23 | 708 | 0.22 | 60 | 0.26 | 171 | 0.14 | 156 | 0.14 | 15 | 0.2 |  |  | ✓ |
| L23 Allergic contact dermatitis | 94 | 0.03 | 87 | 0.03 | 7 | 0.03 | 16 | 0.01 | 16 | 0.01 | 0 | 0 |  |  | ✓ |
| L25 Unspecified contact dermatitis | 448 | 0.13 | 419 | 0.13 | 29 | 0.13 | 206 | 0.17 | 194 | 0.18 | 12 | 0.16 |  |  | ✓ |
| L27 Dermatitis due to substances taken internally | 2231 | 0.66 | 2077 | 0.66 | 154 | 0.67 | 746 | 0.63 | 686 | 0.62 | 60 | 0.8 |  |  | ✓ |
| L28 Lichen simplex chronicus and prurigo | 82 | 0.02 | 78 | 0.02 | 4 | 0.02 | 12 | 0.01 | 11 | 0.01 | 1 | 0.01 |  |  | ✓ |
| L29 Pruritus | 2570 | 0.76 | 2360 | 0.75 | 210 | 0.91 | 833 | 0.71 | 769 | 0.7 | 64 | 0.85 |  |  | ✓ |
| L30 Other dermatitis | 3799 | 1.12 | 3523 | 1.11 | 276 | 1.19 | 1139 | 0.96 | 1072 | 0.97 | 67 | 0.89 |  |  | ✓ |
| L40 Psoriasis | 342 | 0.1 | 313 | 0.1 | 29 | 0.13 | 119 | 0.1 | 111 | 0.1 | 8 | 0.11 |  |  | ✓ |
| L50 Urticaria | 1427 | 0.42 | 1349 | 0.43 | 78 | 0.34 | 406 | 0.34 | 384 | 0.35 | 22 | 0.29 |  |  | ✓ |
| L51 Erythema multiforme | 213 | 0.06 | 192 | 0.06 | 21 | 0.09 | 79 | 0.07 | 72 | 0.07 | 7 | 0.09 |  |  | ✓ |
| L52 Erythema nodosum | 39 | 0.01 | 38 | 0.01 | 1 | 0 | 13 | 0.01 | 11 | 0.01 | 2 | 0.03 |  |  | ✓ |
| L53 Other erythematous conditions | 78 | 0.02 | 72 | 0.02 | 6 | 0.03 | 27 | 0.02 | 25 | 0.02 | 2 | 0.03 |  |  | ✓ |
| L58 Radiodermatitis | 207 | 0.06 | 191 | 0.06 | 16 | 0.07 | 72 | 0.06 | 71 | 0.06 | 1 | 0.01 |  |  | ✓ |
| L60 Nail disorders | 74 | 0.02 | 69 | 0.02 | 5 | 0.02 | 36 | 0.03 | 35 | 0.03 | 1 | 0.01 |  |  | ✓ |
| L70 Acne | 242 | 0.07 | 234 | 0.07 | 8 | 0.03 | 69 | 0.06 | 60 | 0.05 | 9 | 0.12 |  |  | ✓ |
| L72 Follicular cysts of skin and subcutaneous tissue | 181 | 0.05 | 172 | 0.05 | 9 | 0.04 | 51 | 0.04 | 49 | 0.04 | 2 | 0.03 |  |  | ✓ |
| L73 Other follicular disorders | 39 | 0.01 | 35 | 0.01 | 4 | 0.02 | 20 | 0.02 | 20 | 0.02 | 0 | 0 |  |  | ✓ |
| L74 Eccrine sweat disorders | 71 | 0.02 | 63 | 0.02 | 8 | 0.03 | 14 | 0.01 | 13 | 0.01 | 1 | 0.01 |  |  | ✓ |
| L81 Other disorders of pigmentation | 136 | 0.04 | 126 | 0.04 | 10 | 0.04 | 53 | 0.04 | 51 | 0.05 | 2 | 0.03 |  |  | ✓ |
| L82 Seborrhoeic keratosis | 53 | 0.02 | 52 | 0.02 | 1 | 0 | 15 | 0.01 | 14 | 0.01 | 1 | 0.01 |  |  | ✓ |
| L84 Corns and callosities | 50 | 0.01 | 50 | 0.02 | 0 | 0 | 15 | 0.01 | 15 | 0.01 | 0 | 0 |  |  | ✓ |
| L85 Other epidermal thickening | 3779 | 1.11 | 3488 | 1.1 | 291 | 1.26 | 1560 | 1.32 | 1465 | 1.32 | 95 | 1.27 |  | ✓ | ✓ |
| L89 Decubitus ulcer and pressure area | 1854 | 0.55 | 1578 | 0.5 | 276 | 1.19 | 600 | 0.51 | 523 | 0.47 | 77 | 1.03 |  |  | ✓ |
| L90 Atrophic disorders of skin | 58 | 0.02 | 56 | 0.02 | 2 | 0.01 | 20 | 0.02 | 20 | 0.02 | 0 | 0 |  |  | ✓ |
| L91 Hypertrophic disorders of skin | 125 | 0.04 | 121 | 0.04 | 4 | 0.02 | 51 | 0.04 | 49 | 0.04 | 2 | 0.03 |  |  | ✓ |
| L94 Other localized connective tissue disorders | 13 | 0 | 10 | 0 | 3 | 0.01 | 10 | 0.01 | 8 | 0.01 | 2 | 0.03 |  |  | ✓ |
| L97 Ulcer of lower limb, not elsewhere classified | 868 | 0.26 | 801 | 0.25 | 67 | 0.29 | 356 | 0.3 | 328 | 0.3 | 28 | 0.37 |  |  | ✓ |
| L98 Other disorders of skin and subcutaneous tissue, not elsewhere classified | 1134 | 0.33 | 1052 | 0.33 | 82 | 0.35 | 385 | 0.33 | 357 | 0.32 | 28 | 0.37 |  |  | ✓ |
| M00 Pyogenic arthritis | 339 | 0.1 | 310 | 0.1 | 29 | 0.13 | 112 | 0.09 | 108 | 0.1 | 4 | 0.05 |  |  | ✓ |
| M05 Seropositive rheumatoid arthritis | 316 | 0.09 | 281 | 0.09 | 35 | 0.15 | 107 | 0.09 | 95 | 0.09 | 12 | 0.16 |  |  | ✓ |
| M06 Other rheumatoid arthritis | 3267 | 0.96 | 2991 | 0.95 | 276 | 1.19 | 1255 | 1.06 | 1166 | 1.05 | 89 | 1.19 |  |  | ✓ |
| M10 Gout | 1142 | 0.34 | 1072 | 0.34 | 70 | 0.3 | 430 | 0.36 | 408 | 0.37 | 22 | 0.29 |  |  | ✓ |
| M11 Other crystal arthropathies | 944 | 0.28 | 855 | 0.27 | 89 | 0.39 | 351 | 0.3 | 321 | 0.29 | 30 | 0.4 |  |  | ✓ |
| M13 Other arthritis | 700 | 0.21 | 653 | 0.21 | 47 | 0.2 | 212 | 0.18 | 199 | 0.18 | 13 | 0.17 |  |  | ✓ |
| M16 Coxarthrosis [arthrosis of hip] | 1233 | 0.36 | 1203 | 0.38 | 30 | 0.13 | 639 | 0.54 | 628 | 0.57 | 11 | 0.15 |  |  | ✓ |
| M17 Gonarthrosis [arthrosis of knee] | 3040 | 0.9 | 2910 | 0.92 | 130 | 0.56 | 1271 | 1.08 | 1227 | 1.11 | 44 | 0.59 |  |  | ✓ |
| M18 Arthrosis of first carpometacarpal joint | 31 | 0.01 | 30 | 0.01 | 1 | 0 | 10 | 0.01 | 9 | 0.01 | 1 | 0.01 |  |  | ✓ |
| M19 Other arthrosis | 518 | 0.15 | 483 | 0.15 | 35 | 0.15 | 225 | 0.19 | 213 | 0.19 | 12 | 0.16 |  |  | ✓ |
| M20 Acquired deformities of fingers and toes | 86 | 0.03 | 84 | 0.03 | 2 | 0.01 | 31 | 0.03 | 30 | 0.03 | 1 | 0.01 |  |  | ✓ |
| M21 Other acquired deformities of limbs | 90 | 0.03 | 88 | 0.03 | 2 | 0.01 | 31 | 0.03 | 30 | 0.03 | 1 | 0.01 |  |  | ✓ |
| M23 Internal derangement of knee | 84 | 0.02 | 84 | 0.03 | 0 | 0 | 33 | 0.03 | 33 | 0.03 | 0 | 0 |  |  | ✓ |
| M24 Other specific joint derangements | 383 | 0.11 | 373 | 0.12 | 10 | 0.04 | 283 | 0.24 | 278 | 0.25 | 5 | 0.07 |  |  | ✓ |
| M25 Other joint disorders, not elsewhere classified | 1255 | 0.37 | 1193 | 0.38 | 62 | 0.27 | 406 | 0.34 | 388 | 0.35 | 18 | 0.24 |  |  | ✓ |
| M30 Polyarteritis nodosa and related conditions | 1070 | 0.32 | 1031 | 0.33 | 39 | 0.17 | 392 | 0.33 | 380 | 0.34 | 12 | 0.16 |  |  | ✓ |
| M31 Other necrotizing vasculopathies | 631 | 0.19 | 552 | 0.17 | 79 | 0.34 | 217 | 0.18 | 187 | 0.17 | 30 | 0.4 |  |  | ✓ |
| M32 Systemic lupus erythematosus | 562 | 0.17 | 516 | 0.16 | 46 | 0.2 | 152 | 0.13 | 139 | 0.13 | 13 | 0.17 |  |  | ✓ |
| M33 Dermatopolymyositis | 316 | 0.09 | 295 | 0.09 | 21 | 0.09 | 71 | 0.06 | 68 | 0.06 | 3 | 0.04 |  |  | ✓ |
| M34 Systemic sclerosis | 260 | 0.08 | 228 | 0.07 | 32 | 0.14 | 103 | 0.09 | 93 | 0.08 | 10 | 0.13 |  |  | ✓ |
| M35 Other systemic involvement of connective tissue | 1133 | 0.33 | 1044 | 0.33 | 89 | 0.39 | 479 | 0.41 | 451 | 0.41 | 28 | 0.37 |  |  | ✓ |
| M40 Kyphosis and lordosis | 38 | 0.01 | 36 | 0.01 | 2 | 0.01 | 21 | 0.02 | 21 | 0.02 | 0 | 0 |  |  | ✓ |
| M41 Scoliosis | 199 | 0.06 | 194 | 0.06 | 5 | 0.02 | 102 | 0.09 | 98 | 0.09 | 4 | 0.05 |  |  | ✓ |
| M43 Other deforming dorsopathies | 1051 | 0.31 | 1013 | 0.32 | 38 | 0.16 | 381 | 0.32 | 366 | 0.33 | 15 | 0.2 |  |  | ✓ |
| M45 Ankylosing spondylitis | 32 | 0.01 | 29 | 0.01 | 3 | 0.01 | 15 | 0.01 | 14 | 0.01 | 1 | 0.01 |  |  | ✓ |
| M46 Other inflammatory spondylopathies | 392 | 0.12 | 355 | 0.11 | 37 | 0.16 | 118 | 0.1 | 113 | 0.1 | 5 | 0.07 |  |  | ✓ |
| M47 Spondylosis | 2375 | 0.7 | 2262 | 0.71 | 113 | 0.49 | 790 | 0.67 | 759 | 0.69 | 31 | 0.41 |  |  | ✓ |
| M48 Other spondylopathies | 6520 | 1.92 | 6243 | 1.97 | 277 | 1.2 | 2242 | 1.9 | 2150 | 1.94 | 92 | 1.23 |  | ✓ | ✓ |
| M50 Cervical disc disorders | 177 | 0.05 | 172 | 0.05 | 5 | 0.02 | 57 | 0.05 | 56 | 0.05 | 1 | 0.01 |  |  | ✓ |
| M51 Other intervertebral disc disorders | 2004 | 0.59 | 1951 | 0.62 | 53 | 0.23 | 762 | 0.65 | 743 | 0.67 | 19 | 0.25 |  |  | ✓ |
| M53 Other dorsopathies, not elsewhere classified | 469 | 0.14 | 456 | 0.14 | 13 | 0.06 | 118 | 0.1 | 111 | 0.1 | 7 | 0.09 |  |  | ✓ |
| M54 Dorsalgia | 10471 | 3.08 | 9590 | 3.03 | 881 | 3.81 | 3249 | 2.75 | 3008 | 2.72 | 241 | 3.21 |  | ✓ | ✓ |
| M60 Myositis | 78 | 0.02 | 72 | 0.02 | 6 | 0.03 | 23 | 0.02 | 23 | 0.02 | 0 | 0 |  |  | ✓ |
| M62 Other disorders of muscle | 18122 | 5.34 | 15567 | 4.92 | 2555 | 11.06 | 7952 | 6.73 | 7021 | 6.35 | 931 | 12.4 | ✓ | ✓ | ✓ |
| M65 Synovitis and tenosynovitis | 277 | 0.08 | 267 | 0.08 | 10 | 0.04 | 115 | 0.1 | 112 | 0.1 | 3 | 0.04 |  |  | ✓ |
| M67 Other disorders of synovium and tendon | 67 | 0.02 | 67 | 0.02 | 0 | 0 | 23 | 0.02 | 23 | 0.02 | 0 | 0 |  |  | ✓ |
| M70 Soft tissue disorders related to use, overuse and pressure | 81 | 0.02 | 77 | 0.02 | 4 | 0.02 | 27 | 0.02 | 25 | 0.02 | 2 | 0.03 |  |  | ✓ |
| M71 Other bursopathies | 54 | 0.02 | 51 | 0.02 | 3 | 0.01 | 26 | 0.02 | 23 | 0.02 | 3 | 0.04 |  |  | ✓ |
| M72 Fibroblastic disorders | 125 | 0.04 | 115 | 0.04 | 10 | 0.04 | 53 | 0.04 | 47 | 0.04 | 6 | 0.08 |  |  | ✓ |
| M75 Shoulder lesions | 790 | 0.23 | 741 | 0.23 | 49 | 0.21 | 295 | 0.25 | 277 | 0.25 | 18 | 0.24 |  |  | ✓ |
| M77 Other enthesopathies | 42 | 0.01 | 41 | 0.01 | 1 | 0 | 18 | 0.02 | 16 | 0.01 | 2 | 0.03 |  |  | ✓ |
| M79 Other soft tissue disorders, not elsewhere classified | 854 | 0.25 | 788 | 0.25 | 66 | 0.29 | 245 | 0.21 | 223 | 0.2 | 22 | 0.29 |  |  | ✓ |
| M80 Osteoporosis with pathological fracture | 958 | 0.28 | 909 | 0.29 | 49 | 0.21 | 417 | 0.35 | 393 | 0.36 | 24 | 0.32 |  |  | ✓ |
| M81 Osteoporosis without pathological fracture | 11925 | 3.51 | 11082 | 3.5 | 843 | 3.65 | 4381 | 3.71 | 4124 | 3.73 | 257 | 3.42 |  | ✓ | ✓ |
| M84 Disorders of continuity of bone | 471 | 0.14 | 441 | 0.14 | 30 | 0.13 | 136 | 0.12 | 128 | 0.12 | 8 | 0.11 |  |  | ✓ |
| M85 Other disorders of bone density and structure | 35 | 0.01 | 35 | 0.01 | 0 | 0 | 10 | 0.01 | 10 | 0.01 | 0 | 0 |  |  | ✓ |
| M86 Osteomyelitis | 378 | 0.11 | 355 | 0.11 | 23 | 0.1 | 146 | 0.12 | 133 | 0.12 | 13 | 0.17 |  |  | ✓ |
| M87 Osteonecrosis | 338 | 0.1 | 322 | 0.1 | 16 | 0.07 | 148 | 0.13 | 142 | 0.13 | 6 | 0.08 |  |  | ✓ |
| M89 Other disorders of bone | 48 | 0.01 | 47 | 0.01 | 1 | 0 | 16 | 0.01 | 16 | 0.01 | 0 | 0 |  |  | ✓ |
| M93 Other osteochondropathies | 49 | 0.01 | 48 | 0.02 | 1 | 0 | 12 | 0.01 | 12 | 0.01 | 0 | 0 |  |  | ✓ |
| M96 Postprocedural musculoskeletal disorders, not elsewhere classified | 106 | 0.03 | 95 | 0.03 | 11 | 0.05 | 75 | 0.06 | 69 | 0.06 | 6 | 0.08 |  |  | ✓ |
| M99 Biomechanical lesions, not elsewhere classified | 157 | 0.05 | 152 | 0.05 | 5 | 0.02 | 80 | 0.07 | 79 | 0.07 | 1 | 0.01 |  |  | ✓ |
| N00 Acute nephritic syndrome | 88 | 0.03 | 84 | 0.03 | 4 | 0.02 | 30 | 0.03 | 30 | 0.03 | 0 | 0 |  |  | ✓ |
| N01 Rapidly progressive nephritic syndrome | 221 | 0.07 | 201 | 0.06 | 20 | 0.09 | 75 | 0.06 | 63 | 0.06 | 12 | 0.16 |  |  | ✓ |
| N02 Recurrent and persistent haematuria | 667 | 0.2 | 645 | 0.2 | 22 | 0.1 | 296 | 0.25 | 285 | 0.26 | 11 | 0.15 |  |  | ✓ |
| N03 Chronic nephritic syndrome | 353 | 0.1 | 344 | 0.11 | 9 | 0.04 | 133 | 0.11 | 127 | 0.11 | 6 | 0.08 |  |  | ✓ |
| N04 Nephrotic syndrome | 1717 | 0.51 | 1575 | 0.5 | 142 | 0.61 | 626 | 0.53 | 565 | 0.51 | 61 | 0.81 |  |  | ✓ |
| N05 Unspecified nephritic syndrome | 330 | 0.1 | 310 | 0.1 | 20 | 0.09 | 98 | 0.08 | 88 | 0.08 | 10 | 0.13 |  |  | ✓ |
| N10 Acute tubulo-interstitial nephritis | 5110 | 1.51 | 4578 | 1.45 | 532 | 2.3 | 1740 | 1.47 | 1567 | 1.42 | 173 | 2.3 |  | ✓ | ✓ |
| N11 Chronic tubulo-interstitial nephritis | 124 | 0.04 | 110 | 0.03 | 14 | 0.06 | 52 | 0.04 | 44 | 0.04 | 8 | 0.11 |  |  | ✓ |
| N12 Tubulo-interstitial nephritis, not specified as acute or chronic | 674 | 0.2 | 604 | 0.19 | 70 | 0.3 | 226 | 0.19 | 206 | 0.19 | 20 | 0.27 |  |  | ✓ |
| N13 Obstructive and reflux uropathy | 3834 | 1.13 | 3396 | 1.07 | 438 | 1.9 | 1591 | 1.35 | 1417 | 1.28 | 174 | 2.32 |  | ✓ | ✓ |
| N14 Drug- and heavy-metal-induced tubulo-interstitial and tubular conditions | 76 | 0.02 | 71 | 0.02 | 5 | 0.02 | 23 | 0.02 | 20 | 0.02 | 3 | 0.04 |  |  | ✓ |
| N15 Other renal tubulo-interstitial diseases | 117 | 0.03 | 109 | 0.03 | 8 | 0.03 | 27 | 0.02 | 24 | 0.02 | 3 | 0.04 |  |  | ✓ |
| N17 Acute renal failure | 2622 | 0.77 | 2326 | 0.74 | 296 | 1.28 | 1013 | 0.86 | 920 | 0.83 | 93 | 1.24 |  |  | ✓ |
| N18 Chronic kidney disease | 17642 | 5.2 | 15834 | 5 | 1808 | 7.82 | 7039 | 5.96 | 6330 | 5.73 | 709 | 9.44 | ✓ | ✓ | ✓ |
| N19 Unspecified kidney failure | 3132 | 0.92 | 2769 | 0.88 | 363 | 1.57 | 1116 | 0.95 | 1002 | 0.91 | 114 | 1.52 |  |  | ✓ |
| N20 Calculus of kidney and ureter | 2088 | 0.61 | 1984 | 0.63 | 104 | 0.45 | 921 | 0.78 | 874 | 0.79 | 47 | 0.63 |  |  | ✓ |
| N21 Calculus of lower urinary tract | 326 | 0.1 | 310 | 0.1 | 16 | 0.07 | 177 | 0.15 | 166 | 0.15 | 11 | 0.15 |  |  | ✓ |
| N25 Disorders resulting from impaired renal tubular function | 59 | 0.02 | 55 | 0.02 | 4 | 0.02 | 25 | 0.02 | 22 | 0.02 | 3 | 0.04 |  |  | ✓ |
| N26 Unspecified contracted kidney | 125 | 0.04 | 119 | 0.04 | 6 | 0.03 | 39 | 0.03 | 36 | 0.03 | 3 | 0.04 |  |  | ✓ |
| N28 Other disorders of kidney and ureter, not elsewhere classified | 2481 | 0.73 | 2268 | 0.72 | 213 | 0.92 | 956 | 0.81 | 879 | 0.79 | 77 | 1.03 |  |  | ✓ |
| N30 Cystitis | 1532 | 0.45 | 1403 | 0.44 | 129 | 0.56 | 574 | 0.49 | 528 | 0.48 | 46 | 0.61 |  |  | ✓ |
| N31 Neuromuscular dysfunction of bladder, not elsewhere classified | 2630 | 0.77 | 2383 | 0.75 | 247 | 1.07 | 1003 | 0.85 | 918 | 0.83 | 85 | 1.13 |  |  | ✓ |
| N32 Other disorders of bladder | 2628 | 0.77 | 2411 | 0.76 | 217 | 0.94 | 1065 | 0.9 | 975 | 0.88 | 90 | 1.2 |  |  | ✓ |
| N34 Urethritis and urethral syndrome | 39 | 0.01 | 31 | 0.01 | 8 | 0.03 | 11 | 0.01 | 11 | 0.01 | 0 | 0 |  |  | ✓ |
| N35 Urethral stricture | 170 | 0.05 | 159 | 0.05 | 11 | 0.05 | 68 | 0.06 | 59 | 0.05 | 9 | 0.12 |  |  | ✓ |
| N36 Other disorders of urethra | 63 | 0.02 | 59 | 0.02 | 4 | 0.02 | 22 | 0.02 | 19 | 0.02 | 3 | 0.04 |  |  | ✓ |
| N39 Other disorders of urinary system | 8233 | 2.42 | 7363 | 2.33 | 870 | 3.76 | 2950 | 2.5 | 2654 | 2.4 | 296 | 3.94 |  | ✓ | ✓ |
| N40 Hyperplasia of prostate | 11730 | 3.45 | 10717 | 3.39 | 1013 | 4.38 | 4281 | 3.63 | 3935 | 3.56 | 346 | 4.61 |  | ✓ | ✓ |
| N41 Inflammatory diseases of prostate | 566 | 0.17 | 516 | 0.16 | 50 | 0.22 | 193 | 0.16 | 182 | 0.16 | 11 | 0.15 |  |  | ✓ |
| N42 Other disorders of prostate | 34 | 0.01 | 31 | 0.01 | 3 | 0.01 | 24 | 0.02 | 22 | 0.02 | 2 | 0.03 |  |  | ✓ |
| N43 Hydrocele and spermatocele | 126 | 0.04 | 120 | 0.04 | 6 | 0.03 | 38 | 0.03 | 37 | 0.03 | 1 | 0.01 |  |  | ✓ |
| N44 Torsion of testis | 53 | 0.02 | 53 | 0.02 | 0 | 0 | 17 | 0.01 | 17 | 0.02 | 0 | 0 |  |  | ✓ |
| N45 Orchitis and epididymitis | 156 | 0.05 | 145 | 0.05 | 11 | 0.05 | 49 | 0.04 | 48 | 0.04 | 1 | 0.01 |  |  | ✓ |
| N47 Redundant prepuce, phimosis and paraphimosis | 42 | 0.01 | 40 | 0.01 | 2 | 0.01 | 22 | 0.02 | 20 | 0.02 | 2 | 0.03 |  |  | ✓ |
| N48 Other disorders of penis | 50 | 0.01 | 46 | 0.01 | 4 | 0.02 | 20 | 0.02 | 20 | 0.02 | 0 | 0 |  |  | ✓ |
| N49 Inflammatory disorders of male genital organs, not elsewhere classified | 23 | 0.01 | 22 | 0.01 | 1 | 0 | 10 | 0.01 | 9 | 0.01 | 1 | 0.01 |  |  | ✓ |
| N63 Unspecified lump in breast | 68 | 0.02 | 66 | 0.02 | 2 | 0.01 | 18 | 0.02 | 15 | 0.01 | 3 | 0.04 |  |  | ✓ |
| N64 Other disorders of breast | 60 | 0.02 | 58 | 0.02 | 2 | 0.01 | 12 | 0.01 | 10 | 0.01 | 2 | 0.03 |  |  | ✓ |
| N70 Salpingitis and oophoritis | 265 | 0.08 | 262 | 0.08 | 3 | 0.01 | 93 | 0.08 | 90 | 0.08 | 3 | 0.04 |  |  | ✓ |
| N71 Inflammatory disease of uterus, except cervix | 92 | 0.03 | 84 | 0.03 | 8 | 0.03 | 33 | 0.03 | 33 | 0.03 | 0 | 0 |  |  | ✓ |
| N72 Inflammatory disease of cervix uteri | 27 | 0.01 | 26 | 0.01 | 1 | 0 | 19 | 0.02 | 18 | 0.02 | 1 | 0.01 |  |  | ✓ |
| N73 Other female pelvic inflammatory diseases | 353 | 0.1 | 342 | 0.11 | 11 | 0.05 | 172 | 0.15 | 162 | 0.15 | 10 | 0.13 |  |  | ✓ |
| N76 Other inflammation of vagina and vulva | 122 | 0.04 | 114 | 0.04 | 8 | 0.03 | 31 | 0.03 | 30 | 0.03 | 1 | 0.01 |  |  | ✓ |
| N80 Endometriosis | 1375 | 0.4 | 1338 | 0.42 | 37 | 0.16 | 469 | 0.4 | 462 | 0.42 | 7 | 0.09 |  |  | ✓ |
| N81 Female genital prolapse | 759 | 0.22 | 743 | 0.23 | 16 | 0.07 | 239 | 0.2 | 237 | 0.21 | 2 | 0.03 |  |  | ✓ |
| N82 Fistulae involving female genital tract | 72 | 0.02 | 62 | 0.02 | 10 | 0.04 | 13 | 0.01 | 11 | 0.01 | 2 | 0.03 |  |  | ✓ |
| N83 Noninflammatory disorders of ovary, fallopian tube and broad ligament | 337 | 0.1 | 332 | 0.1 | 5 | 0.02 | 169 | 0.14 | 166 | 0.15 | 3 | 0.04 |  |  | ✓ |
| N84 Polyp of female genital tract | 236 | 0.07 | 235 | 0.07 | 1 | 0 | 99 | 0.08 | 99 | 0.09 | 0 | 0 |  |  | ✓ |
| N85 Other noninflammatory disorders of uterus, except cervix | 264 | 0.08 | 259 | 0.08 | 5 | 0.02 | 115 | 0.1 | 112 | 0.1 | 3 | 0.04 |  |  | ✓ |
| N86 Erosion and ectropion of cervix uteri | 234 | 0.07 | 217 | 0.07 | 17 | 0.07 | 69 | 0.06 | 65 | 0.06 | 4 | 0.05 |  |  | ✓ |
| N87 Dysplasia of cervix uteri | 470 | 0.14 | 466 | 0.15 | 4 | 0.02 | 271 | 0.23 | 265 | 0.24 | 6 | 0.08 |  |  | ✓ |
| N92 Excessive, frequent and irregular menstruation | 231 | 0.07 | 218 | 0.07 | 13 | 0.06 | 85 | 0.07 | 80 | 0.07 | 5 | 0.07 |  |  | ✓ |
| N93 Other abnormal uterine and vaginal bleeding | 199 | 0.06 | 184 | 0.06 | 15 | 0.06 | 62 | 0.05 | 57 | 0.05 | 5 | 0.07 |  |  | ✓ |
| N94 Pain and other conditions associated with female genital organs and menstrual cycle | 146 | 0.04 | 142 | 0.04 | 4 | 0.02 | 47 | 0.04 | 45 | 0.04 | 2 | 0.03 |  |  | ✓ |
| N95 Menopausal and other perimenopausal disorders | 140 | 0.04 | 136 | 0.04 | 4 | 0.02 | 54 | 0.05 | 54 | 0.05 | 0 | 0 |  |  | ✓ |
| N98 Complications associated with artificial fertilization | 18 | 0.01 | 18 | 0.01 | 0 | 0 | 10 | 0.01 | 10 | 0.01 | 0 | 0 |  |  | ✓ |
| N99 Postprocedural disorders of genitourinary system, not elsewhere classified | 129 | 0.04 | 123 | 0.04 | 6 | 0.03 | 66 | 0.06 | 62 | 0.06 | 4 | 0.05 |  |  | ✓ |
| O20 Haemorrhage in early pregnancy | 54 | 0.02 | 50 | 0.02 | 4 | 0.02 | 13 | 0.01 | 12 | 0.01 | 1 | 0.01 |  |  | ✓ |
| O99 Other maternal diseases classifiable elsewhere but complicating pregnancy, childbirth and the puerperium | 26 | 0.01 | 26 | 0.01 | 0 | 0 | 12 | 0.01 | 11 | 0.01 | 1 | 0.01 |  |  | ✓ |
| P27 Chronic respiratory disease originating in the perinatal period | 28 | 0.01 | 26 | 0.01 | 2 | 0.01 | 13 | 0.01 | 12 | 0.01 | 1 | 0.01 |  |  | ✓ |
| P61 Other perinatal haematological disorders | 13 | 0 | 10 | 0 | 3 | 0.01 | 10 | 0.01 | 10 | 0.01 | 0 | 0 |  |  | ✓ |
| P81 Other disturbances of temperature regulation of newborn | 21 | 0.01 | 20 | 0.01 | 1 | 0 | 12 | 0.01 | 12 | 0.01 | 0 | 0 |  |  | ✓ |
| P92 Feeding problems of newborn | 31 | 0.01 | 31 | 0.01 | 0 | 0 | 16 | 0.01 | 16 | 0.01 | 0 | 0 |  |  | ✓ |
| Q04 Other congenital malformations of brain | 48 | 0.01 | 31 | 0.01 | 17 | 0.07 | 12 | 0.01 | 11 | 0.01 | 1 | 0.01 |  |  | ✓ |
| Q05 Spina bifida | 16 | 0 | 14 | 0 | 2 | 0.01 | 14 | 0.01 | 11 | 0.01 | 3 | 0.04 |  |  | ✓ |
| Q20 Congenital malformations of cardiac chambers and connections | 67 | 0.02 | 58 | 0.02 | 9 | 0.04 | 28 | 0.02 | 24 | 0.02 | 4 | 0.05 |  |  | ✓ |
| Q21 Congenital malformations of cardiac septa | 542 | 0.16 | 497 | 0.16 | 45 | 0.19 | 166 | 0.14 | 149 | 0.13 | 17 | 0.23 |  |  | ✓ |
| Q22 Congenital malformations of pulmonary and tricuspid valves | 21 | 0.01 | 19 | 0.01 | 2 | 0.01 | 16 | 0.01 | 15 | 0.01 | 1 | 0.01 |  |  | ✓ |
| Q23 Congenital malformations of aortic and mitral valves | 70 | 0.02 | 63 | 0.02 | 7 | 0.03 | 21 | 0.02 | 19 | 0.02 | 2 | 0.03 |  |  | ✓ |
| Q24 Other congenital malformations of heart | 28 | 0.01 | 24 | 0.01 | 4 | 0.02 | 14 | 0.01 | 14 | 0.01 | 0 | 0 |  |  | ✓ |
| Q25 Congenital malformations of great arteries | 175 | 0.05 | 164 | 0.05 | 11 | 0.05 | 72 | 0.06 | 65 | 0.06 | 7 | 0.09 |  |  | ✓ |
| Q28 Other congenital malformations of circulatory system | 68 | 0.02 | 68 | 0.02 | 0 | 0 | 16 | 0.01 | 15 | 0.01 | 1 | 0.01 |  |  | ✓ |
| Q31 Congenital malformations of larynx | 25 | 0.01 | 21 | 0.01 | 4 | 0.02 | 16 | 0.01 | 13 | 0.01 | 3 | 0.04 |  |  | ✓ |
| Q43 Other congenital malformations of intestine | 86 | 0.03 | 83 | 0.03 | 3 | 0.01 | 32 | 0.03 | 30 | 0.03 | 2 | 0.03 |  |  | ✓ |
| Q44 Congenital malformations of gallbladder, bile ducts and liver | 125 | 0.04 | 108 | 0.03 | 17 | 0.07 | 45 | 0.04 | 39 | 0.04 | 6 | 0.08 |  |  | ✓ |
| Q45 Other congenital malformations of digestive system | 17 | 0.01 | 16 | 0.01 | 1 | 0 | 11 | 0.01 | 9 | 0.01 | 2 | 0.03 |  |  | ✓ |
| Q61 Cystic kidney disease | 135 | 0.04 | 123 | 0.04 | 12 | 0.05 | 87 | 0.07 | 78 | 0.07 | 9 | 0.12 |  |  | ✓ |
| Q62 Congenital obstructive defects of renal pelvis and congenital malformations of ureter | 112 | 0.03 | 100 | 0.03 | 12 | 0.05 | 39 | 0.03 | 38 | 0.03 | 1 | 0.01 |  |  | ✓ |
| Q64 Other congenital malformations of urinary system | 18 | 0.01 | 18 | 0.01 | 0 | 0 | 11 | 0.01 | 10 | 0.01 | 1 | 0.01 |  |  | ✓ |
| Q85 Phakomatoses, not elsewhere classified | 65 | 0.02 | 58 | 0.02 | 7 | 0.03 | 21 | 0.02 | 20 | 0.02 | 1 | 0.01 |  |  | ✓ |
| Q87 Other specified congenital malformation syndromes affecting multiple systems | 98 | 0.03 | 87 | 0.03 | 11 | 0.05 | 38 | 0.03 | 33 | 0.03 | 5 | 0.07 |  |  | ✓ |
| Q89 Other congenital malformations, not elsewhere classified | 43 | 0.01 | 33 | 0.01 | 10 | 0.04 | 17 | 0.01 | 17 | 0.02 | 0 | 0 |  |  | ✓ |
| Q90 Down syndrome | 222 | 0.07 | 179 | 0.06 | 43 | 0.19 | 91 | 0.08 | 79 | 0.07 | 12 | 0.16 |  |  | ✓ |
| R00 Abnormalities of heart beat | 2637 | 0.78 | 2476 | 0.78 | 161 | 0.7 | 954 | 0.81 | 905 | 0.82 | 49 | 0.65 |  |  | ✓ |
| R02 Gangrene, not elsewhere classified | 263 | 0.08 | 240 | 0.08 | 23 | 0.1 | 91 | 0.08 | 82 | 0.07 | 9 | 0.12 |  |  | ✓ |
| R03 Abnormal blood-pressure reading, without diagnosis | 1996 | 0.59 | 1908 | 0.6 | 88 | 0.38 | 1079 | 0.91 | 1016 | 0.92 | 63 | 0.84 |  |  | ✓ |
| R04 Haemorrhage from respiratory passages | 1241 | 0.37 | 1098 | 0.35 | 143 | 0.62 | 358 | 0.3 | 326 | 0.29 | 32 | 0.43 |  |  | ✓ |
| R05 Cough | 371 | 0.11 | 334 | 0.11 | 37 | 0.16 | 121 | 0.1 | 111 | 0.1 | 10 | 0.13 |  |  | ✓ |
| R06 Abnormalities of breathing | 564 | 0.17 | 488 | 0.15 | 76 | 0.33 | 235 | 0.2 | 194 | 0.18 | 41 | 0.55 |  |  | ✓ |
| R07 Pain in throat and chest | 301 | 0.09 | 289 | 0.09 | 12 | 0.05 | 118 | 0.1 | 108 | 0.1 | 10 | 0.13 |  |  | ✓ |
| R09 Other symptoms and signs involving the circulatory and respiratory systems | 2579 | 0.76 | 2382 | 0.75 | 197 | 0.85 | 722 | 0.61 | 652 | 0.59 | 70 | 0.93 |  |  | ✓ |
| R10 Abdominal and pelvic pain | 1879 | 0.55 | 1756 | 0.55 | 123 | 0.53 | 688 | 0.58 | 647 | 0.59 | 41 | 0.55 |  |  | ✓ |
| R11 Nausea and vomiting | 19087 | 5.62 | 17824 | 5.63 | 1263 | 5.47 | 6057 | 5.13 | 5638 | 5.1 | 419 | 5.58 | ✓ | ✓ | ✓ |
| R13 Dysphagia | 5858 | 1.73 | 5068 | 1.6 | 790 | 3.42 | 2200 | 1.86 | 1949 | 1.76 | 251 | 3.34 |  | ✓ | ✓ |
| R14 Flatulence and related conditions | 2560 | 0.75 | 2365 | 0.75 | 195 | 0.84 | 1015 | 0.86 | 943 | 0.85 | 72 | 0.96 |  |  | ✓ |
| R16 Hepatomegaly and splenomegaly, not elsewhere classified | 103 | 0.03 | 95 | 0.03 | 8 | 0.03 | 38 | 0.03 | 37 | 0.03 | 1 | 0.01 |  |  | ✓ |
| R17 Hyperbilirubinaemia, with or without jaundice, not elsewhere classified | 158 | 0.05 | 138 | 0.04 | 20 | 0.09 | 62 | 0.05 | 57 | 0.05 | 5 | 0.07 |  |  | ✓ |
| R18 Ascites | 2099 | 0.62 | 1608 | 0.51 | 491 | 2.12 | 603 | 0.51 | 490 | 0.44 | 113 | 1.51 |  |  | ✓ |
| R19 Other symptoms and signs involving the digestive system and abdomen | 2115 | 0.62 | 1880 | 0.59 | 235 | 1.02 | 562 | 0.48 | 508 | 0.46 | 54 | 0.72 |  |  | ✓ |
| R20 Disturbances of skin sensation | 237 | 0.07 | 229 | 0.07 | 8 | 0.03 | 77 | 0.07 | 73 | 0.07 | 4 | 0.05 |  |  | ✓ |
| R21 Rash and other nonspecific skin eruption | 132 | 0.04 | 124 | 0.04 | 8 | 0.03 | 44 | 0.04 | 41 | 0.04 | 3 | 0.04 |  |  | ✓ |
| R22 Localized swelling, mass and lump of skin and subcutaneous tissue | 191 | 0.06 | 180 | 0.06 | 11 | 0.05 | 49 | 0.04 | 44 | 0.04 | 5 | 0.07 |  |  | ✓ |
| R23 Other skin changes | 91 | 0.03 | 79 | 0.02 | 12 | 0.05 | 26 | 0.02 | 25 | 0.02 | 1 | 0.01 |  |  | ✓ |
| R25 Abnormal involuntary movements | 299 | 0.09 | 264 | 0.08 | 35 | 0.15 | 93 | 0.08 | 85 | 0.08 | 8 | 0.11 |  |  | ✓ |
| R26 Abnormalities of gait and mobility | 2832 | 0.83 | 2513 | 0.79 | 319 | 1.38 | 456 | 0.39 | 425 | 0.38 | 31 | 0.41 |  |  | ✓ |
| R29 Other symptoms and signs involving the nervous and musculoskeletal systems | 365 | 0.11 | 344 | 0.11 | 21 | 0.09 | 86 | 0.07 | 76 | 0.07 | 10 | 0.13 |  |  | ✓ |
| R31 Unspecified haematuria | 811 | 0.24 | 730 | 0.23 | 81 | 0.35 | 278 | 0.24 | 252 | 0.23 | 26 | 0.35 |  |  | ✓ |
| R32 Unspecified urinary incontinence | 15 | 0 | 14 | 0 | 1 | 0 | 31 | 0.03 | 30 | 0.03 | 1 | 0.01 |  |  | ✓ |
| R33 Retention of urine | 705 | 0.21 | 637 | 0.2 | 68 | 0.29 | 320 | 0.27 | 281 | 0.25 | 39 | 0.52 |  |  | ✓ |
| R34 Anuria and oliguria | 32 | 0.01 | 29 | 0.01 | 3 | 0.01 | 20 | 0.02 | 18 | 0.02 | 2 | 0.03 |  |  | ✓ |
| R35 Polyuria | 212 | 0.06 | 201 | 0.06 | 11 | 0.05 | 68 | 0.06 | 64 | 0.06 | 4 | 0.05 |  |  | ✓ |
| R39 Other symptoms and signs involving the urinary system | 842 | 0.25 | 782 | 0.25 | 60 | 0.26 | 409 | 0.35 | 390 | 0.35 | 19 | 0.25 |  |  | ✓ |
| R40 Somnolence, stupor and coma | 5780 | 1.7 | 5318 | 1.68 | 462 | 2 | 1661 | 1.41 | 1541 | 1.39 | 120 | 1.6 |  | ✓ | ✓ |
| R41 Other symptoms and signs involving cognitive functions and awareness | 114 | 0.03 | 111 | 0.04 | 3 | 0.01 | 39 | 0.03 | 36 | 0.03 | 3 | 0.04 |  |  | ✓ |
| R42 Dizziness and giddiness | 1499 | 0.44 | 1423 | 0.45 | 76 | 0.33 | 473 | 0.4 | 458 | 0.41 | 15 | 0.2 |  |  | ✓ |
| R45 Symptoms and signs involving emotional state | 157 | 0.05 | 130 | 0.04 | 27 | 0.12 | 41 | 0.03 | 39 | 0.04 | 2 | 0.03 |  |  | ✓ |
| R46 Symptoms and signs involving appearance and behaviour | 75 | 0.02 | 73 | 0.02 | 2 | 0.01 | 28 | 0.02 | 28 | 0.03 | 0 | 0 |  |  | ✓ |
| R47 Speech disturbances, not elsewhere classified | 873 | 0.26 | 846 | 0.27 | 27 | 0.12 | 187 | 0.16 | 182 | 0.16 | 5 | 0.07 |  |  | ✓ |
| R49 Voice disturbances | 71 | 0.02 | 67 | 0.02 | 4 | 0.02 | 14 | 0.01 | 13 | 0.01 | 1 | 0.01 |  |  | ✓ |
| R50 Fever of other and unknown origin | 1640 | 0.48 | 1485 | 0.47 | 155 | 0.67 | 516 | 0.44 | 481 | 0.44 | 35 | 0.47 |  |  | ✓ |
| R51 Headache | 1472 | 0.43 | 1400 | 0.44 | 72 | 0.31 | 494 | 0.42 | 481 | 0.44 | 13 | 0.17 |  |  | ✓ |
| R52 Pain, not elsewhere classified | 12095 | 3.56 | 9859 | 3.12 | 2236 | 9.68 | 3902 | 3.3 | 3266 | 2.95 | 636 | 8.47 |  | ✓ | ✓ |
| R53 Malaise and fatigue | 137 | 0.04 | 114 | 0.04 | 23 | 0.1 | 119 | 0.1 | 102 | 0.09 | 17 | 0.23 |  |  | ✓ |
| R54 Senility | 48 | 0.01 | 43 | 0.01 | 5 | 0.02 | 10 | 0.01 | 8 | 0.01 | 2 | 0.03 |  |  | ✓ |
| R55 Syncope and collapse | 352 | 0.1 | 337 | 0.11 | 15 | 0.06 | 147 | 0.12 | 140 | 0.13 | 7 | 0.09 |  |  | ✓ |
| R56 Convulsions, not elsewhere classified | 2574 | 0.76 | 2430 | 0.77 | 144 | 0.62 | 903 | 0.76 | 851 | 0.77 | 52 | 0.69 |  |  | ✓ |
| R57 Shock, not elsewhere classified | 10750 | 3.17 | 10088 | 3.19 | 662 | 2.86 | 3659 | 3.1 | 3458 | 3.13 | 201 | 2.68 |  | ✓ | ✓ |
| R58 Haemorrhage, not elsewhere classified | 1173 | 0.35 | 1117 | 0.35 | 56 | 0.24 | 469 | 0.4 | 454 | 0.41 | 15 | 0.2 |  |  | ✓ |
| R59 Enlarged lymph nodes | 409 | 0.12 | 385 | 0.12 | 24 | 0.1 | 113 | 0.1 | 107 | 0.1 | 6 | 0.08 |  |  | ✓ |
| R60 Oedema, not elsewhere classified | 1989 | 0.59 | 1663 | 0.53 | 326 | 1.41 | 701 | 0.59 | 616 | 0.56 | 85 | 1.13 |  |  | ✓ |
| R62 Lack of expected normal physiological development | 86 | 0.03 | 77 | 0.02 | 9 | 0.04 | 36 | 0.03 | 33 | 0.03 | 3 | 0.04 |  |  | ✓ |
| R63 Symptoms and signs concerning food and fluid intake | 2666 | 0.79 | 2265 | 0.72 | 401 | 1.74 | 920 | 0.78 | 808 | 0.73 | 112 | 1.49 |  |  | ✓ |
| R65 Systemic Inflammatory Response Syndrome [SIRS] | 164 | 0.05 | 146 | 0.05 | 18 | 0.08 | 20 | 0.02 | 19 | 0.02 | 1 | 0.01 |  |  | ✓ |
| R68 Other general symptoms and signs | 94 | 0.03 | 76 | 0.02 | 18 | 0.08 | 43 | 0.04 | 35 | 0.03 | 8 | 0.11 |  |  | ✓ |
| R73 Elevated blood glucose level | 834 | 0.25 | 800 | 0.25 | 34 | 0.15 | 261 | 0.22 | 252 | 0.23 | 9 | 0.12 |  |  | ✓ |
| R74 Abnormal serum enzyme levels | 171 | 0.05 | 159 | 0.05 | 12 | 0.05 | 78 | 0.07 | 74 | 0.07 | 4 | 0.05 |  |  | ✓ |
| R76 Other abnormal immunological findings in serum | 105 | 0.03 | 98 | 0.03 | 7 | 0.03 | 66 | 0.06 | 62 | 0.06 | 4 | 0.05 |  |  | ✓ |
| R77 Other abnormalities of plasma proteins | 31 | 0.01 | 30 | 0.01 | 1 | 0 | 11 | 0.01 | 10 | 0.01 | 1 | 0.01 |  |  | ✓ |
| R79 Other abnormal findings of blood chemistry | 139 | 0.04 | 126 | 0.04 | 13 | 0.06 | 31 | 0.03 | 29 | 0.03 | 2 | 0.03 |  |  | ✓ |
| R80 Isolated proteinuria | 115 | 0.03 | 108 | 0.03 | 7 | 0.03 | 31 | 0.03 | 29 | 0.03 | 2 | 0.03 |  |  | ✓ |
| R82 Other abnormal findings in urine | 190 | 0.06 | 179 | 0.06 | 11 | 0.05 | 57 | 0.05 | 53 | 0.05 | 4 | 0.05 |  |  | ✓ |
| R91 Abnormal findings on diagnostic imaging of lung | 166 | 0.05 | 159 | 0.05 | 7 | 0.03 | 53 | 0.04 | 53 | 0.05 | 0 | 0 |  |  | ✓ |
| R93 Abnormal findings on diagnostic imaging of other body structures | 32 | 0.01 | 28 | 0.01 | 4 | 0.02 | 14 | 0.01 | 13 | 0.01 | 1 | 0.01 |  |  | ✓ |
| R94 Abnormal results of function studies | 367 | 0.11 | 341 | 0.11 | 26 | 0.11 | 131 | 0.11 | 122 | 0.11 | 9 | 0.12 |  |  | ✓ |
| S00 Superficial injury of head | 1472 | 0.43 | 1400 | 0.44 | 72 | 0.31 | 450 | 0.38 | 428 | 0.39 | 22 | 0.29 |  |  | ✓ |
| S01 Open wound of head | 1144 | 0.34 | 1089 | 0.34 | 55 | 0.24 | 349 | 0.3 | 336 | 0.3 | 13 | 0.17 |  |  | ✓ |
| S02 Fracture of skull and facial bones | 610 | 0.18 | 590 | 0.19 | 20 | 0.09 | 178 | 0.15 | 175 | 0.16 | 3 | 0.04 |  |  | ✓ |
| S03 Dislocation, sprain and strain of joints and ligaments of head | 18 | 0.01 | 16 | 0.01 | 2 | 0.01 | 10 | 0.01 | 9 | 0.01 | 1 | 0.01 |  |  | ✓ |
| S04 Injury of cranial nerves | 55 | 0.02 | 54 | 0.02 | 1 | 0 | 16 | 0.01 | 16 | 0.01 | 0 | 0 |  |  | ✓ |
| S05 Injury of eye and orbit | 90 | 0.03 | 88 | 0.03 | 2 | 0.01 | 25 | 0.02 | 25 | 0.02 | 0 | 0 |  |  | ✓ |
| S06 Intracranial injury | 2944 | 0.87 | 2735 | 0.86 | 209 | 0.9 | 1077 | 0.91 | 1016 | 0.92 | 61 | 0.81 |  |  | ✓ |
| S09 Other and unspecified injuries of head | 170 | 0.05 | 163 | 0.05 | 7 | 0.03 | 47 | 0.04 | 44 | 0.04 | 3 | 0.04 |  |  | ✓ |
| S10 Superficial injury of neck | 40 | 0.01 | 39 | 0.01 | 1 | 0 | 15 | 0.01 | 15 | 0.01 | 0 | 0 |  |  | ✓ |
| S12 Fracture of neck | 197 | 0.06 | 192 | 0.06 | 5 | 0.02 | 62 | 0.05 | 62 | 0.06 | 0 | 0 |  |  | ✓ |
| S13 Dislocation, sprain and strain of joints and ligaments at neck level | 273 | 0.08 | 263 | 0.08 | 10 | 0.04 | 60 | 0.05 | 58 | 0.05 | 2 | 0.03 |  |  | ✓ |
| S14 Injury of nerves and spinal cord at neck level | 296 | 0.09 | 287 | 0.09 | 9 | 0.04 | 96 | 0.08 | 95 | 0.09 | 1 | 0.01 |  |  | ✓ |
| S20 Superficial injury of thorax | 193 | 0.06 | 183 | 0.06 | 10 | 0.04 | 54 | 0.05 | 53 | 0.05 | 1 | 0.01 |  |  | ✓ |
| S22 Fracture of rib(s), sternum and thoracic spine | 2187 | 0.64 | 2069 | 0.65 | 118 | 0.51 | 864 | 0.73 | 816 | 0.74 | 48 | 0.64 |  |  | ✓ |
| S24 Injury of nerves and spinal cord at thorax level | 27 | 0.01 | 25 | 0.01 | 2 | 0.01 | 13 | 0.01 | 11 | 0.01 | 2 | 0.03 |  |  | ✓ |
| S27 Injury of other and unspecified intrathoracic organs | 672 | 0.2 | 646 | 0.2 | 26 | 0.11 | 202 | 0.17 | 193 | 0.17 | 9 | 0.12 |  |  | ✓ |
| S29 Other and unspecified injuries of thorax | 21 | 0.01 | 21 | 0.01 | 0 | 0 | 13 | 0.01 | 13 | 0.01 | 0 | 0 |  |  | ✓ |
| S30 Superficial injury of abdomen, lower back and pelvis | 423 | 0.12 | 408 | 0.13 | 15 | 0.06 | 150 | 0.13 | 143 | 0.13 | 7 | 0.09 |  |  | ✓ |
| S31 Open wound of abdomen, lower back and pelvis | 95 | 0.03 | 90 | 0.03 | 5 | 0.02 | 12 | 0.01 | 12 | 0.01 | 0 | 0 |  |  | ✓ |
| S32 Fracture of lumbar spine and pelvis | 3117 | 0.92 | 2910 | 0.92 | 207 | 0.9 | 1256 | 1.06 | 1177 | 1.06 | 79 | 1.05 |  |  | ✓ |
| S36 Injury of intra-abdominal organs | 268 | 0.08 | 257 | 0.08 | 11 | 0.05 | 76 | 0.06 | 75 | 0.07 | 1 | 0.01 |  |  | ✓ |
| S37 Injury of urinary and pelvic organs | 190 | 0.06 | 184 | 0.06 | 6 | 0.03 | 68 | 0.06 | 62 | 0.06 | 6 | 0.08 |  |  | ✓ |
| S39 Other and unspecified injuries of abdomen, lower back and pelvis | 33 | 0.01 | 33 | 0.01 | 0 | 0 | 24 | 0.02 | 24 | 0.02 | 0 | 0 |  |  | ✓ |
| S40 Superficial injury of shoulder and upper arm | 138 | 0.04 | 135 | 0.04 | 3 | 0.01 | 54 | 0.05 | 49 | 0.04 | 5 | 0.07 |  |  | ✓ |
| S42 Fracture of shoulder and upper arm | 2086 | 0.61 | 2013 | 0.64 | 73 | 0.32 | 765 | 0.65 | 744 | 0.67 | 21 | 0.28 |  |  | ✓ |
| S43 Dislocation, sprain and strain of joints and ligaments of shoulder girdle | 124 | 0.04 | 119 | 0.04 | 5 | 0.02 | 41 | 0.03 | 41 | 0.04 | 0 | 0 |  |  | ✓ |
| S46 Injury of muscle and tendon at shoulder and upper arm level | 444 | 0.13 | 436 | 0.14 | 8 | 0.03 | 213 | 0.18 | 210 | 0.19 | 3 | 0.04 |  |  | ✓ |
| S50 Superficial injury of forearm | 125 | 0.04 | 121 | 0.04 | 4 | 0.02 | 38 | 0.03 | 37 | 0.03 | 1 | 0.01 |  |  | ✓ |
| S51 Open wound of forearm | 132 | 0.04 | 124 | 0.04 | 8 | 0.03 | 46 | 0.04 | 43 | 0.04 | 3 | 0.04 |  |  | ✓ |
| S52 Fracture of forearm | 2052 | 0.6 | 2029 | 0.64 | 23 | 0.1 | 717 | 0.61 | 705 | 0.64 | 12 | 0.16 |  |  | ✓ |
| S53 Dislocation, sprain and strain of joints and ligaments of elbow | 50 | 0.01 | 50 | 0.02 | 0 | 0 | 22 | 0.02 | 22 | 0.02 | 0 | 0 |  |  | ✓ |
| S54 Injury of nerves at forearm level | 48 | 0.01 | 48 | 0.02 | 0 | 0 | 23 | 0.02 | 23 | 0.02 | 0 | 0 |  |  | ✓ |
| S56 Injury of muscle and tendon at forearm level | 193 | 0.06 | 193 | 0.06 | 0 | 0 | 62 | 0.05 | 61 | 0.06 | 1 | 0.01 |  |  | ✓ |
| S60 Superficial injury of wrist and hand | 83 | 0.02 | 78 | 0.02 | 5 | 0.02 | 25 | 0.02 | 25 | 0.02 | 0 | 0 |  |  | ✓ |
| S61 Open wound of wrist and hand | 243 | 0.07 | 236 | 0.07 | 7 | 0.03 | 76 | 0.06 | 76 | 0.07 | 0 | 0 |  |  | ✓ |
| S62 Fracture at wrist and hand level | 530 | 0.16 | 522 | 0.16 | 8 | 0.03 | 189 | 0.16 | 188 | 0.17 | 1 | 0.01 |  |  | ✓ |
| S63 Dislocation, sprain and strain of joints and ligaments at wrist and hand level | 63 | 0.02 | 63 | 0.02 | 0 | 0 | 18 | 0.02 | 18 | 0.02 | 0 | 0 |  |  | ✓ |
| S64 Injury of nerves at wrist and hand level | 37 | 0.01 | 37 | 0.01 | 0 | 0 | 10 | 0.01 | 10 | 0.01 | 0 | 0 |  |  | ✓ |
| S67 Crushing injury of wrist and hand | 44 | 0.01 | 43 | 0.01 | 1 | 0 | 11 | 0.01 | 11 | 0.01 | 0 | 0 |  |  | ✓ |
| S68 Traumatic amputation of wrist and hand | 110 | 0.03 | 109 | 0.03 | 1 | 0 | 29 | 0.02 | 29 | 0.03 | 0 | 0 |  |  | ✓ |
| S70 Superficial injury of hip and thigh | 152 | 0.04 | 138 | 0.04 | 14 | 0.06 | 49 | 0.04 | 47 | 0.04 | 2 | 0.03 |  |  | ✓ |
| S71 Open wound of hip and thigh | 42 | 0.01 | 42 | 0.01 | 0 | 0 | 18 | 0.02 | 18 | 0.02 | 0 | 0 |  |  | ✓ |
| S72 Fracture of femur | 4318 | 1.27 | 4097 | 1.29 | 221 | 0.96 | 1620 | 1.37 | 1555 | 1.41 | 65 | 0.87 |  | ✓ | ✓ |
| S73 Dislocation, sprain and strain of joint and ligaments of hip | 39 | 0.01 | 35 | 0.01 | 4 | 0.02 | 22 | 0.02 | 20 | 0.02 | 2 | 0.03 |  |  | ✓ |
| S76 Injury of muscle and tendon at hip and thigh level | 49 | 0.01 | 47 | 0.01 | 2 | 0.01 | 27 | 0.02 | 27 | 0.02 | 0 | 0 |  |  | ✓ |
| S80 Superficial injury of lower leg | 347 | 0.1 | 332 | 0.1 | 15 | 0.06 | 118 | 0.1 | 114 | 0.1 | 4 | 0.05 |  |  | ✓ |
| S81 Open wound of lower leg | 233 | 0.07 | 224 | 0.07 | 9 | 0.04 | 77 | 0.07 | 72 | 0.07 | 5 | 0.07 |  |  | ✓ |
| S82 Fracture of lower leg, including ankle | 2277 | 0.67 | 2237 | 0.71 | 40 | 0.17 | 784 | 0.66 | 769 | 0.7 | 15 | 0.2 |  |  | ✓ |
| S83 Dislocation, sprain and strain of joints and ligaments of knee | 664 | 0.2 | 658 | 0.21 | 6 | 0.03 | 282 | 0.24 | 277 | 0.25 | 5 | 0.07 |  |  | ✓ |
| S86 Injury of muscle and tendon at lower leg level | 338 | 0.1 | 332 | 0.1 | 6 | 0.03 | 102 | 0.09 | 99 | 0.09 | 3 | 0.04 |  |  | ✓ |
| S89 Other and unspecified injuries of lower leg | 30 | 0.01 | 29 | 0.01 | 1 | 0 | 12 | 0.01 | 12 | 0.01 | 0 | 0 |  |  | ✓ |
| S90 Superficial injury of ankle and foot | 34 | 0.01 | 33 | 0.01 | 1 | 0 | 19 | 0.02 | 19 | 0.02 | 0 | 0 |  |  | ✓ |
| S91 Open wound of ankle and foot | 95 | 0.03 | 94 | 0.03 | 1 | 0 | 56 | 0.05 | 54 | 0.05 | 2 | 0.03 |  |  | ✓ |
| S92 Fracture of foot, except ankle | 612 | 0.18 | 599 | 0.19 | 13 | 0.06 | 190 | 0.16 | 186 | 0.17 | 4 | 0.05 |  |  | ✓ |
| S93 Dislocation, sprain and strain of joints and ligaments at ankle and foot level | 165 | 0.05 | 161 | 0.05 | 4 | 0.02 | 49 | 0.04 | 49 | 0.04 | 0 | 0 |  |  | ✓ |
| S96 Injury of muscle and tendon at ankle and foot level | 23 | 0.01 | 23 | 0.01 | 0 | 0 | 12 | 0.01 | 12 | 0.01 | 0 | 0 |  |  | ✓ |
| S98 Traumatic amputation of ankle and foot | 23 | 0.01 | 23 | 0.01 | 0 | 0 | 10 | 0.01 | 8 | 0.01 | 2 | 0.03 |  |  | ✓ |
| T00 Superficial injuries involving multiple body regions | 247 | 0.07 | 243 | 0.08 | 4 | 0.02 | 75 | 0.06 | 71 | 0.06 | 4 | 0.05 |  |  | ✓ |
| T02 Fractures involving multiple body regions | 479 | 0.14 | 445 | 0.14 | 34 | 0.15 | 206 | 0.17 | 188 | 0.17 | 18 | 0.24 |  |  | ✓ |
| T07 Unspecified multiple injuries | 59 | 0.02 | 58 | 0.02 | 1 | 0 | 13 | 0.01 | 13 | 0.01 | 0 | 0 |  |  | ✓ |
| T08 Fracture of spine, level unspecified | 117 | 0.03 | 100 | 0.03 | 17 | 0.07 | 32 | 0.03 | 30 | 0.03 | 2 | 0.03 |  |  | ✓ |
| T09 Other injuries of spine and trunk, level unspecified | 134 | 0.04 | 122 | 0.04 | 12 | 0.05 | 32 | 0.03 | 31 | 0.03 | 1 | 0.01 |  |  | ✓ |
| T13 Other injuries of lower limb, level unspecified | 35 | 0.01 | 33 | 0.01 | 2 | 0.01 | 12 | 0.01 | 11 | 0.01 | 1 | 0.01 |  |  | ✓ |
| T14 Injury of unspecified body region | 1216 | 0.36 | 1142 | 0.36 | 74 | 0.32 | 351 | 0.3 | 332 | 0.3 | 19 | 0.25 |  |  | ✓ |
| T17 Foreign body in respiratory tract | 121 | 0.04 | 115 | 0.04 | 6 | 0.03 | 44 | 0.04 | 40 | 0.04 | 4 | 0.05 |  |  | ✓ |
| T18 Foreign body in alimentary tract | 256 | 0.08 | 238 | 0.08 | 18 | 0.08 | 104 | 0.09 | 101 | 0.09 | 3 | 0.04 |  |  | ✓ |
| T19 Foreign body in genitourinary tract | 17 | 0.01 | 17 | 0.01 | 0 | 0 | 11 | 0.01 | 11 | 0.01 | 0 | 0 |  |  | ✓ |
| T20 Burn and corrosion of head and neck | 54 | 0.02 | 50 | 0.02 | 4 | 0.02 | 15 | 0.01 | 15 | 0.01 | 0 | 0 |  |  | ✓ |
| T21 Burn and corrosion of trunk | 85 | 0.03 | 80 | 0.03 | 5 | 0.02 | 29 | 0.02 | 29 | 0.03 | 0 | 0 |  |  | ✓ |
| T22 Burn and corrosion of shoulder and upper limb, except wrist and hand | 71 | 0.02 | 66 | 0.02 | 5 | 0.02 | 14 | 0.01 | 14 | 0.01 | 0 | 0 |  |  | ✓ |
| T23 Burn and corrosion of wrist and hand | 41 | 0.01 | 39 | 0.01 | 2 | 0.01 | 14 | 0.01 | 14 | 0.01 | 0 | 0 |  |  | ✓ |
| T24 Burn and corrosion of hip and lower limb, except ankle and foot | 113 | 0.03 | 106 | 0.03 | 7 | 0.03 | 33 | 0.03 | 32 | 0.03 | 1 | 0.01 |  |  | ✓ |
| T25 Burn and corrosion of ankle and foot | 55 | 0.02 | 51 | 0.02 | 4 | 0.02 | 12 | 0.01 | 12 | 0.01 | 0 | 0 |  |  | ✓ |
| T29 Burns and corrosions of multiple body regions | 70 | 0.02 | 68 | 0.02 | 2 | 0.01 | 17 | 0.01 | 17 | 0.02 | 0 | 0 |  |  | ✓ |
| T30 Burn and corrosion, body region unspecified | 24 | 0.01 | 22 | 0.01 | 2 | 0.01 | 12 | 0.01 | 12 | 0.01 | 0 | 0 |  |  | ✓ |
| T42 Poisoning by antiepileptic, sedative-hypnotic and antiparkinsonism drugs | 216 | 0.06 | 201 | 0.06 | 15 | 0.06 | 47 | 0.04 | 46 | 0.04 | 1 | 0.01 |  |  | ✓ |
| T43 Poisoning by psychotropic drugs, not elsewhere classified | 45 | 0.01 | 43 | 0.01 | 2 | 0.01 | 16 | 0.01 | 15 | 0.01 | 1 | 0.01 |  |  | ✓ |
| T46 Poisoning by agents primarily affecting the cardiovascular system | 29 | 0.01 | 26 | 0.01 | 3 | 0.01 | 12 | 0.01 | 11 | 0.01 | 1 | 0.01 |  |  | ✓ |
| T50 Poisoning by diuretics and other and unspecified drugs, medicaments and biological substances | 346 | 0.1 | 333 | 0.11 | 13 | 0.06 | 111 | 0.09 | 108 | 0.1 | 3 | 0.04 |  |  | ✓ |
| T58 Toxic effect of carbon monoxide | 71 | 0.02 | 68 | 0.02 | 3 | 0.01 | 20 | 0.02 | 20 | 0.02 | 0 | 0 |  |  | ✓ |
| T63 Toxic effect of contact with venomous animals | 283 | 0.08 | 279 | 0.09 | 4 | 0.02 | 63 | 0.05 | 61 | 0.06 | 2 | 0.03 |  |  | ✓ |
| T67 Effects of heat and light | 672 | 0.2 | 634 | 0.2 | 38 | 0.16 | 364 | 0.31 | 344 | 0.31 | 20 | 0.27 |  |  | ✓ |
| T68 Hypothermia | 178 | 0.05 | 164 | 0.05 | 14 | 0.06 | 66 | 0.06 | 60 | 0.05 | 6 | 0.08 |  |  | ✓ |
| T75 Effects of other external causes | 71 | 0.02 | 66 | 0.02 | 5 | 0.02 | 18 | 0.02 | 18 | 0.02 | 0 | 0 |  |  | ✓ |
| T78 Adverse effects, not elsewhere classified | 1037 | 0.31 | 992 | 0.31 | 45 | 0.19 | 336 | 0.28 | 330 | 0.3 | 6 | 0.08 |  |  | ✓ |
| T79 Certain early complications of trauma, not elsewhere classified | 323 | 0.1 | 308 | 0.1 | 15 | 0.06 | 131 | 0.11 | 128 | 0.12 | 3 | 0.04 |  |  | ✓ |
| T80 Complications following infusion, transfusion and therapeutic injection | 117 | 0.03 | 98 | 0.03 | 19 | 0.08 | 51 | 0.04 | 43 | 0.04 | 8 | 0.11 |  |  | ✓ |
| T81 Complications of procedures, not elsewhere classified | 13502 | 3.98 | 12785 | 4.04 | 717 | 3.1 | 4743 | 4.02 | 4511 | 4.08 | 232 | 3.09 |  | ✓ | ✓ |
| T82 Complications of cardiac and vascular prosthetic devices, implants and grafts | 2313 | 0.68 | 2141 | 0.68 | 172 | 0.74 | 722 | 0.61 | 665 | 0.6 | 57 | 0.76 |  |  | ✓ |
| T83 Complications of genitourinary prosthetic devices, implants and grafts | 155 | 0.05 | 131 | 0.04 | 24 | 0.1 | 48 | 0.04 | 40 | 0.04 | 8 | 0.11 |  |  | ✓ |
| T84 Complications of internal orthopaedic prosthetic devices, implants and grafts | 302 | 0.09 | 289 | 0.09 | 13 | 0.06 | 83 | 0.07 | 76 | 0.07 | 7 | 0.09 |  |  | ✓ |
| T85 Complications of other internal prosthetic devices, implants and grafts | 348 | 0.1 | 286 | 0.09 | 62 | 0.27 | 100 | 0.08 | 77 | 0.07 | 23 | 0.31 |  |  | ✓ |
| T86 Failure and rejection of transplanted organs and tissues | 80 | 0.02 | 66 | 0.02 | 14 | 0.06 | 35 | 0.03 | 29 | 0.03 | 6 | 0.08 |  |  | ✓ |
| T88 Other complications of surgical and medical care, not elsewhere classified | 12454 | 3.67 | 12068 | 3.81 | 386 | 1.67 | 5507 | 4.66 | 5359 | 4.85 | 148 | 1.97 |  | ✓ | ✓ |
| T90 Sequelae of injuries of head | 191 | 0.06 | 176 | 0.06 | 15 | 0.06 | 43 | 0.04 | 39 | 0.04 | 4 | 0.05 |  |  | ✓ |
| T91 Sequelae of injuries of neck and trunk | 407 | 0.12 | 377 | 0.12 | 30 | 0.13 | 142 | 0.12 | 135 | 0.12 | 7 | 0.09 |  |  | ✓ |
| Z08 Follow-up examination after treatment for malignant neoplasms | 79 | 0.02 | 74 | 0.02 | 5 | 0.02 | 35 | 0.03 | 34 | 0.03 | 1 | 0.01 |  |  | ✓ |
| Z22 Carrier of infectious disease | 276 | 0.08 | 260 | 0.08 | 16 | 0.07 | 99 | 0.08 | 91 | 0.08 | 8 | 0.11 |  |  | ✓ |
| Z33 Pregnant state, incidental | 76 | 0.02 | 71 | 0.02 | 5 | 0.02 | 16 | 0.01 | 16 | 0.01 | 0 | 0 |  |  | ✓ |
| Z90 Acquired absence of organs, not elsewhere classified | 747 | 0.22 | 668 | 0.21 | 79 | 0.34 | 261 | 0.22 | 232 | 0.21 | 29 | 0.39 |  |  | ✓ |
| Z92 Personal history of medical treatment | 142 | 0.04 | 138 | 0.04 | 4 | 0.02 | 52 | 0.04 | 50 | 0.05 | 2 | 0.03 |  |  | ✓ |
| Z93 Artificial opening status | 2398 | 0.71 | 2089 | 0.66 | 309 | 1.34 | 882 | 0.75 | 783 | 0.71 | 99 | 1.32 |  |  | ✓ |
| Z94 Transplanted organ and tissue status | 251 | 0.07 | 231 | 0.07 | 20 | 0.09 | 110 | 0.09 | 97 | 0.09 | 13 | 0.17 |  |  | ✓ |
| Z95 Presence of cardiac and vascular implants and grafts | 4478 | 1.32 | 4186 | 1.32 | 292 | 1.26 | 1796 | 1.52 | 1693 | 1.53 | 103 | 1.37 |  | ✓ | ✓ |
| Z96 Presence of other functional implants | 477 | 0.14 | 459 | 0.15 | 18 | 0.08 | 226 | 0.19 | 214 | 0.19 | 12 | 0.16 |  |  | ✓ |
| Z98 Other postsurgical states | 84 | 0.02 | 77 | 0.02 | 7 | 0.03 | 48 | 0.04 | 45 | 0.04 | 3 | 0.04 |  |  | ✓ |
