## Supplementary Table S2 for "Comparison of machine-learning and logistic regression models to predict 30-day unplanned readmission: a development and validation study"

| **Supplementary Table S2. Details and frequency of surgery codes (Japanese original codes)** | | | | | | | | | | | | | | | |
| --- | --- | --- | --- | --- | --- | --- | --- | --- | --- | --- | --- | --- | --- | --- | --- |
| Variables | Derivation dataset (N=339,513) | | | | | | Validation dataset (N=118,074) | | | | | | Use of the variable | | |
|  | Total  N=339,513 | | Outcome (-)  N=316,405 | | Outcome (+)  N=23,108 | | Total  N=118,074 | | Outcome (-)  N=110,567 | | Outcome (+)  N=7,507 | | Data pattern  1&2 | Data pattern  3&4 | Data pattern  5&6 |
|  | n | % | n | % | n | % | n | % | n | % | n | % |  |  |  |
| K000 Wound treatment | 26274 | 7.74 | 25039 | 7.91 | 1235 | 5.34 | 9623 | 8.15 | 9234 | 8.35 | 389 | 5.18 | ✓ | ✓ | ✓ |
| K001 Dermatoplasty | 900 | 0.27 | 837 | 0.26 | 63 | 0.27 | 236 | 0.2 | 222 | 0.2 | 14 | 0.19 |  |  | ✓ |
| K002 Debridement | 430 | 0.13 | 415 | 0.13 | 15 | 0.06 | 131 | 0.11 | 128 | 0.12 | 3 | 0.04 |  |  | ✓ |
| K005 Extraction of skin and subcutaneous tumors (the exposed area) | 118 | 0.03 | 113 | 0.04 | 5 | 0.02 | 34 | 0.03 | 34 | 0.03 | 0 | 0 |  |  | ✓ |
| K006 Extraction of skin and subcutaneous tumors (outside the exposed area) | 208 | 0.06 | 200 | 0.06 | 8 | 0.03 | 82 | 0.07 | 80 | 0.07 | 2 | 0.03 |  |  | ✓ |
| K007 Excision of cutaneous malignancy | 314 | 0.09 | 307 | 0.1 | 7 | 0.03 | 101 | 0.09 | 101 | 0.09 | 0 | 0 |  |  | ✓ |
| K010 Scar constriction surgery | 28 | 0.01 | 28 | 0.01 | 0 | 0 | 14 | 0.01 | 14 | 0.01 | 0 | 0 |  |  | ✓ |
| K013 Split-Layer Implant | 675 | 0.2 | 656 | 0.21 | 19 | 0.08 | 202 | 0.17 | 200 | 0.18 | 2 | 0.03 |  |  | ✓ |
| K015 Skin valve creation, migration, amputation, prolonged valvuloplasty | 237 | 0.07 | 227 | 0.07 | 10 | 0.04 | 70 | 0.06 | 67 | 0.06 | 3 | 0.04 |  |  | ✓ |
| K016 Arterial (skin) and muscular (skin) valves | 142 | 0.04 | 138 | 0.04 | 4 | 0.02 | 46 | 0.04 | 41 | 0.04 | 5 | 0.07 |  |  | ✓ |
| K017 Free skin valves (with vascular pattern under microscope) | 133 | 0.04 | 127 | 0.04 | 6 | 0.03 | 52 | 0.04 | 52 | 0.05 | 0 | 0 |  |  | ✓ |
| K020 Autologous free composite tissue graft with vascular pattern under microscope | 59 | 0.02 | 54 | 0.02 | 5 | 0.02 | 17 | 0.01 | 16 | 0.01 | 1 | 0.01 |  |  | ✓ |
| K022 Reconstructive surgery with tissue expander (per series) | 112 | 0.03 | 110 | 0.03 | 2 | 0.01 | 34 | 0.03 | 30 | 0.03 | 4 | 0.05 |  |  | ✓ |
| K028 Tendon sheath dissection (including the one under the arthroscope) | 37 | 0.01 | 36 | 0.01 | 1 | 0 | 11 | 0.01 | 11 | 0.01 | 0 | 0 |  |  | ✓ |
| K030 Extraction of soft tumor of limbs and trunk | 222 | 0.07 | 218 | 0.07 | 4 | 0.02 | 80 | 0.07 | 80 | 0.07 | 0 | 0 |  |  | ✓ |
| K031 Surgery for soft torso cancers of limbs and trunk | 240 | 0.07 | 236 | 0.07 | 4 | 0.02 | 104 | 0.09 | 100 | 0.09 | 4 | 0.05 |  |  | ✓ |
| K037 Tendon suture | 435 | 0.13 | 429 | 0.14 | 6 | 0.03 | 129 | 0.11 | 126 | 0.11 | 3 | 0.04 |  |  | ✓ |
| K040 Tendon transposition | 45 | 0.01 | 45 | 0.01 | 0 | 0 | 12 | 0.01 | 12 | 0.01 | 0 | 0 |  |  | ✓ |
| K043 Bone crawling | 53 | 0.02 | 52 | 0.02 | 1 | 0 | 21 | 0.02 | 20 | 0.02 | 1 | 0.01 |  |  | ✓ |
| K044 Noninvasive fracture repair | 897 | 0.26 | 862 | 0.27 | 35 | 0.15 | 305 | 0.26 | 292 | 0.26 | 13 | 0.17 |  |  | ✓ |
| K045 Transcutaneous steel wire Stabbing and fixation for Fracture | 753 | 0.22 | 740 | 0.23 | 13 | 0.06 | 225 | 0.19 | 224 | 0.2 | 1 | 0.01 |  |  | ✓ |
| K046 Invasive fracture surgery | 5576 | 1.64 | 5421 | 1.71 | 155 | 0.67 | 2001 | 1.69 | 1951 | 1.76 | 50 | 0.67 |  | ✓ | ✓ |
| K047 Electromagnetic Wave Electrotherapy for Refractory Fractures(per series) | 597 | 0.18 | 583 | 0.18 | 14 | 0.06 | 311 | 0.26 | 303 | 0.27 | 8 | 0.11 |  |  | ✓ |
| K048 Intraosseous foreign body (inserted object is included.) Removal | 1122 | 0.33 | 1113 | 0.35 | 9 | 0.04 | 362 | 0.31 | 356 | 0.32 | 6 | 0.08 |  |  | ✓ |
| K050 Removal of rotten bone | 37 | 0.01 | 36 | 0.01 | 1 | 0 | 30 | 0.03 | 24 | 0.02 | 6 | 0.08 |  |  | ✓ |
| K052 Bone tumor Excision | 104 | 0.03 | 103 | 0.03 | 1 | 0 | 34 | 0.03 | 33 | 0.03 | 1 | 0.01 |  |  | ✓ |
| K053 Bone cancer surgery | 59 | 0.02 | 57 | 0.02 | 2 | 0.01 | 23 | 0.02 | 22 | 0.02 | 1 | 0.01 |  |  | ✓ |
| K054 Osteotomy | 88 | 0.03 | 88 | 0.03 | 0 | 0 | 89 | 0.08 | 86 | 0.08 | 3 | 0.04 |  |  | ✓ |
| K056 Pseudo-joint surgery | 73 | 0.02 | 73 | 0.02 | 0 | 0 | 30 | 0.03 | 30 | 0.03 | 0 | 0 |  |  | ✓ |
| K059 Bone grafting (including cartilage grafting.) | 1942 | 0.57 | 1898 | 0.6 | 44 | 0.19 | 602 | 0.51 | 587 | 0.53 | 15 | 0.2 |  |  | ✓ |
| K060 Calcaneal dissection | 141 | 0.04 | 131 | 0.04 | 10 | 0.04 | 49 | 0.04 | 47 | 0.04 | 2 | 0.03 |  |  | ✓ |
| K061 Noninvasive Reconstruction of dislocated joint | 246 | 0.07 | 230 | 0.07 | 16 | 0.07 | 78 | 0.07 | 78 | 0.07 | 0 | 0 |  |  | ✓ |
| K063 Invasive Reconstruction of dislocated joints | 118 | 0.03 | 114 | 0.04 | 4 | 0.02 | 41 | 0.03 | 39 | 0.04 | 2 | 0.03 |  |  | ✓ |
| K065 Intra-articular foreign body (inserted object is included.) Removal surgery | 110 | 0.03 | 110 | 0.03 | 0 | 0 | 44 | 0.04 | 44 | 0.04 | 0 | 0 |  |  | ✓ |
| K066 Synovectomy | 228 | 0.07 | 225 | 0.07 | 3 | 0.01 | 65 | 0.06 | 63 | 0.06 | 2 | 0.03 |  |  | ✓ |
| K067 Joint mouse removal surgery | 32 | 0.01 | 32 | 0.01 | 0 | 0 | 19 | 0.02 | 19 | 0.02 | 0 | 0 |  |  | ✓ |
| K068 Meniscectomy | 173 | 0.05 | 172 | 0.05 | 1 | 0 | 71 | 0.06 | 69 | 0.06 | 2 | 0.03 |  |  | ✓ |
| K069 Meniscus suture | 143 | 0.04 | 143 | 0.05 | 0 | 0 | 62 | 0.05 | 61 | 0.06 | 1 | 0.01 |  |  | ✓ |
| K073 Invasive Intra-articular fracture surgery | 1143 | 0.34 | 1129 | 0.36 | 14 | 0.06 | 352 | 0.3 | 343 | 0.31 | 9 | 0.12 |  |  | ✓ |
| K074 Ligament fracture suture | 89 | 0.03 | 89 | 0.03 | 0 | 0 | 30 | 0.03 | 30 | 0.03 | 0 | 0 |  |  | ✓ |
| K076 Invasive capsular release | 61 | 0.02 | 60 | 0.02 | 1 | 0 | 10 | 0.01 | 10 | 0.01 | 0 | 0 |  |  | ✓ |
| K078 Invasive fixation of joint | 72 | 0.02 | 72 | 0.02 | 0 | 0 | 25 | 0.02 | 24 | 0.02 | 1 | 0.01 |  |  | ✓ |
| K079 Ligament rupture Formation surgery | 223 | 0.07 | 223 | 0.07 | 0 | 0 | 139 | 0.12 | 138 | 0.12 | 1 | 0.01 |  |  | ✓ |
| K080 Joint formation surgery | 441 | 0.13 | 438 | 0.14 | 3 | 0.01 | 235 | 0.2 | 231 | 0.21 | 4 | 0.05 |  |  | ✓ |
| K081 Artificial bone head insertion | 1081 | 0.32 | 1034 | 0.33 | 47 | 0.2 | 440 | 0.37 | 420 | 0.38 | 20 | 0.27 |  |  | ✓ |
| K082 Artificial joint replacement | 2631 | 0.77 | 2578 | 0.81 | 53 | 0.23 | 1324 | 1.12 | 1301 | 1.18 | 23 | 0.31 |  |  | ✓ |
| K083 Direct retraction by steel wire (first day.) (The manual technique is included in the case of invasive operation) (per 1 inning) | 165 | 0.05 | 161 | 0.05 | 4 | 0.02 | 52 | 0.04 | 50 | 0.05 | 2 | 0.03 |  |  | ✓ |
| K084 Limb amputation (upper wrist, fore wrist, hand, thigh, lower leg, foot) | 372 | 0.11 | 350 | 0.11 | 22 | 0.1 | 128 | 0.11 | 126 | 0.11 | 2 | 0.03 |  |  | ✓ |
| K087 Amputation formation (requiring bone formation) | 102 | 0.03 | 98 | 0.03 | 4 | 0.02 | 41 | 0.03 | 38 | 0.03 | 3 | 0.04 |  |  | ✓ |
| K091 Eruption of claw hand | 24 | 0.01 | 22 | 0.01 | 2 | 0.01 | 15 | 0.01 | 15 | 0.01 | 0 | 0 |  |  | ✓ |
| K093 Hand root canal opening | 128 | 0.04 | 127 | 0.04 | 1 | 0 | 52 | 0.04 | 52 | 0.05 | 0 | 0 |  |  | ✓ |
| K110 Fourth digit dystrophy surgery | 27 | 0.01 | 26 | 0.01 | 1 | 0 | 10 | 0.01 | 10 | 0.01 | 0 | 0 |  |  | ✓ |
| K116 Spinal and pelvic bone curettage | 42 | 0.01 | 42 | 0.01 | 0 | 0 | 10 | 0.01 | 9 | 0.01 | 1 | 0.01 |  |  | ✓ |
| K126 Spinal and pelvic bone (cartilage) | 31 | 0.01 | 31 | 0.01 | 0 | 0 | 11 | 0.01 | 9 | 0.01 | 2 | 0.03 |  |  | ✓ |
| K128 Removal of foreign body (insert) from spine and pelvis | 94 | 0.03 | 92 | 0.03 | 2 | 0.01 | 45 | 0.04 | 45 | 0.04 | 0 | 0 |  |  | ✓ |
| K131 Laminectomy | 129 | 0.04 | 124 | 0.04 | 5 | 0.02 | 20 | 0.02 | 20 | 0.02 | 0 | 0 |  |  | ✓ |
| K134 Laminotomy | 684 | 0.2 | 676 | 0.21 | 8 | 0.03 | 275 | 0.23 | 270 | 0.24 | 5 | 0.07 |  |  | ✓ |
| K136 Spinal and pelvic cancer surgery | 19 | 0.01 | 18 | 0.01 | 1 | 0 | 10 | 0.01 | 9 | 0.01 | 1 | 0.01 |  |  | ✓ |
| K142 Spinal immobilization, laminectomy, and arch formation (Multiple intervertebral and multiple arch cases are included.) | 3781 | 1.11 | 3701 | 1.17 | 80 | 0.35 | 1369 | 1.16 | 1340 | 1.21 | 29 | 0.39 |  | ✓ | ✓ |
| K144 External Spinal Fixation | 24 | 0.01 | 23 | 0.01 | 1 | 0 | 11 | 0.01 | 11 | 0.01 | 0 | 0 |  |  | ✓ |
| K145 Perforated ventricular drainage | 28 | 0.01 | 27 | 0.01 | 1 | 0 | 17 | 0.01 | 16 | 0.01 | 1 | 0.01 |  |  | ✓ |
| K160 Cerebral neurosurgery (craniotomy) | 63 | 0.02 | 63 | 0.02 | 0 | 0 | 27 | 0.02 | 27 | 0.02 | 0 | 0 |  |  | ✓ |
| K164 Intracranial hematoma removal (craniotomy) | 1337 | 0.39 | 1216 | 0.38 | 121 | 0.52 | 498 | 0.42 | 463 | 0.42 | 35 | 0.47 |  |  | ✓ |
| K169 Intracranial tumor resection | 384 | 0.11 | 365 | 0.12 | 19 | 0.08 | 126 | 0.11 | 122 | 0.11 | 4 | 0.05 |  |  | ✓ |
| K171 Transnasal pituitary tumor resection | 59 | 0.02 | 57 | 0.02 | 2 | 0.01 | 30 | 0.03 | 30 | 0.03 | 0 | 0 |  |  | ✓ |
| K174 Hydrocephalus surgery | 178 | 0.05 | 167 | 0.05 | 11 | 0.05 | 59 | 0.05 | 55 | 0.05 | 4 | 0.05 |  |  | ✓ |
| K177 Cerebral aneurysm neck clipping | 330 | 0.1 | 328 | 0.1 | 2 | 0.01 | 117 | 0.1 | 114 | 0.1 | 3 | 0.04 |  |  | ✓ |
| K178 Endovascular cerebral surgery | 328 | 0.1 | 322 | 0.1 | 6 | 0.03 | 126 | 0.11 | 120 | 0.11 | 6 | 0.08 |  |  | ✓ |
| K182 Nerve suture | 82 | 0.02 | 82 | 0.03 | 0 | 0 | 26 | 0.02 | 25 | 0.02 | 1 | 0.01 |  |  | ✓ |
| K188 Nerve dissection | 28 | 0.01 | 28 | 0.01 | 0 | 0 | 14 | 0.01 | 14 | 0.01 | 0 | 0 |  |  | ✓ |
| K189 Spinal cord drainage | 121 | 0.04 | 119 | 0.04 | 2 | 0.01 | 54 | 0.05 | 53 | 0.05 | 1 | 0.01 |  |  | ✓ |
| K191 Spinal cord tumor resection | 31 | 0.01 | 31 | 0.01 | 0 | 0 | 17 | 0.01 | 17 | 0.02 | 0 | 0 |  |  | ✓ |
| K197 Nerve migration | 130 | 0.04 | 130 | 0.04 | 0 | 0 | 38 | 0.03 | 38 | 0.03 | 0 | 0 |  |  | ✓ |
| K276 Retinal photocoagulation | 128 | 0.04 | 124 | 0.04 | 4 | 0.02 | 47 | 0.04 | 45 | 0.04 | 2 | 0.03 |  |  | ✓ |
| K280 Vitreous stalk dissection under microscope | 366 | 0.11 | 349 | 0.11 | 17 | 0.07 | 109 | 0.09 | 107 | 0.1 | 2 | 0.03 |  |  | ✓ |
| K282 Lens reconstruction | 337 | 0.1 | 324 | 0.1 | 13 | 0.06 | 141 | 0.12 | 133 | 0.12 | 8 | 0.11 |  |  | ✓ |
| K300 Tympanotomy | 481 | 0.14 | 449 | 0.14 | 32 | 0.14 | 141 | 0.12 | 126 | 0.11 | 15 | 0.2 |  |  | ✓ |
| K305 Papillotomy | 194 | 0.06 | 193 | 0.06 | 1 | 0 | 64 | 0.05 | 63 | 0.06 | 1 | 0.01 |  |  | ✓ |
| K309 Tympanic membrane (fluid drainage, air exchange) insertion | 56 | 0.02 | 52 | 0.02 | 4 | 0.02 | 11 | 0.01 | 9 | 0.01 | 2 | 0.03 |  |  | ✓ |
| K319 Tympanoplasty | 221 | 0.07 | 220 | 0.07 | 1 | 0 | 72 | 0.06 | 71 | 0.06 | 1 | 0.01 |  |  | ✓ |
| K331 nasal mucosa cautery | 246 | 0.07 | 228 | 0.07 | 18 | 0.08 | 78 | 0.07 | 64 | 0.06 | 14 | 0.19 |  |  | ✓ |
| K333 Fixation of nasal bone fracture | 51 | 0.02 | 50 | 0.02 | 1 | 0 | 12 | 0.01 | 12 | 0.01 | 0 | 0 |  |  | ✓ |
| K340 Nasal velum extraction | 244 | 0.07 | 239 | 0.08 | 5 | 0.02 | 74 | 0.06 | 73 | 0.07 | 1 | 0.01 |  |  | ✓ |
| K347 Septum correction | 158 | 0.05 | 157 | 0.05 | 1 | 0 | 46 | 0.04 | 45 | 0.04 | 1 | 0.01 |  |  | ✓ |
| K368 Perineural abscess incision | 477 | 0.14 | 463 | 0.15 | 14 | 0.06 | 170 | 0.14 | 163 | 0.15 | 7 | 0.09 |  |  | ✓ |
| K372 Middle pharyngeal tumor extraction | 23 | 0.01 | 22 | 0.01 | 1 | 0 | 10 | 0.01 | 10 | 0.01 | 0 | 0 |  |  | ✓ |
| K374 Pharyngeal head cancer surgery (soft mouth cap cancer surgery is included.) | 47 | 0.01 | 45 | 0.01 | 2 | 0.01 | 10 | 0.01 | 9 | 0.01 | 1 | 0.01 |  |  | ✓ |
| K377 Lenticule surgery | 565 | 0.17 | 532 | 0.17 | 33 | 0.14 | 149 | 0.13 | 139 | 0.13 | 10 | 0.13 |  |  | ✓ |
| K384 Laryngeal abscess incision | 54 | 0.02 | 52 | 0.02 | 2 | 0.01 | 22 | 0.02 | 21 | 0.02 | 1 | 0.01 |  |  | ✓ |
| K386 Tracheotomy | 412 | 0.12 | 377 | 0.12 | 35 | 0.15 | 127 | 0.11 | 122 | 0.11 | 5 | 0.07 |  |  | ✓ |
| K389 Laryngeal and vocal cord pump excision | 37 | 0.01 | 36 | 0.01 | 1 | 0 | 13 | 0.01 | 13 | 0.01 | 0 | 0 |  |  | ✓ |
| K393 Laryngeal tumor resection | 132 | 0.04 | 131 | 0.04 | 1 | 0 | 39 | 0.03 | 38 | 0.03 | 1 | 0.01 |  |  | ✓ |
| K394 Surgery for malignant tumor of larynx | 84 | 0.02 | 84 | 0.03 | 0 | 0 | 22 | 0.02 | 21 | 0.02 | 1 | 0.01 |  |  | ✓ |
| K395 Surgery for malignant tumor of larynx and hypopharynx (including reconstruction by manipulation of neck, chest, abdomen, etc.) | 56 | 0.02 | 51 | 0.02 | 5 | 0.02 | 18 | 0.02 | 15 | 0.01 | 3 | 0.04 |  |  | ✓ |
| K396 Closure of tracheostomy | 69 | 0.02 | 68 | 0.02 | 1 | 0 | 13 | 0.01 | 13 | 0.01 | 0 | 0 |  |  | ✓ |
| K400 Laryngoplasty | 25 | 0.01 | 24 | 0.01 | 1 | 0 | 11 | 0.01 | 10 | 0.01 | 1 | 0.01 |  |  | ✓ |
| K413 Tongue tumor Extraction | 12 | 0 | 11 | 0 | 1 | 0 | 12 | 0.01 | 12 | 0.01 | 0 | 0 |  |  | ✓ |
| K415 Surgery for lingual tumor | 92 | 0.03 | 90 | 0.03 | 2 | 0.01 | 43 | 0.04 | 42 | 0.04 | 1 | 0.01 |  |  | ✓ |
| K425 Excision of aphthous cancers of the mouth, jaw and face | 15 | 0 | 14 | 0 | 1 | 0 | 10 | 0.01 | 10 | 0.01 | 0 | 0 |  |  | ✓ |
| K454 Submandibular gland extraction | 57 | 0.02 | 57 | 0.02 | 0 | 0 | 17 | 0.01 | 17 | 0.02 | 0 | 0 |  |  | ✓ |
| K457 Extraction of subauricular gland tumor | 260 | 0.08 | 257 | 0.08 | 3 | 0.01 | 76 | 0.06 | 76 | 0.07 | 0 | 0 |  |  | ✓ |
| K458 Subauricular gland cancer surgery | 59 | 0.02 | 58 | 0.02 | 1 | 0 | 15 | 0.01 | 15 | 0.01 | 0 | 0 |  |  | ✓ |
| K461 Partial thyroidectomy, thyroidectomy | 311 | 0.09 | 310 | 0.1 | 1 | 0 | 100 | 0.08 | 97 | 0.09 | 3 | 0.04 |  |  | ✓ |
| K462 Baceldo total thyroid excision (sub-total excision) (both lobes) | 70 | 0.02 | 70 | 0.02 | 0 | 0 | 14 | 0.01 | 13 | 0.01 | 1 | 0.01 |  |  | ✓ |
| K463 Surgery for thyroid cancer | 627 | 0.18 | 622 | 0.2 | 5 | 0.02 | 214 | 0.18 | 213 | 0.19 | 1 | 0.01 |  |  | ✓ |
| K464 Parathyroid (epithelial body) adenomatous overgrowth surgery | 101 | 0.03 | 99 | 0.03 | 2 | 0.01 | 28 | 0.02 | 28 | 0.03 | 0 | 0 |  |  | ✓ |
| K469 Cervical fistulotomy | 134 | 0.04 | 132 | 0.04 | 2 | 0.01 | 37 | 0.03 | 36 | 0.03 | 1 | 0.01 |  |  | ✓ |
| K474 Breast Tumor Extraction | 115 | 0.03 | 114 | 0.04 | 1 | 0 | 31 | 0.03 | 31 | 0.03 | 0 | 0 |  |  | ✓ |
| K476 Mammary tumor surgery | 3523 | 1.04 | 3479 | 1.1 | 44 | 0.19 | 1081 | 0.92 | 1071 | 0.97 | 10 | 0.13 |  |  | ✓ |
| K484 Extraction of chest wall cancer | 34 | 0.01 | 33 | 0.01 | 1 | 0 | 11 | 0.01 | 11 | 0.01 | 0 | 0 |  |  | ✓ |
| K488 Experimental thoracotomy | 154 | 0.05 | 146 | 0.05 | 8 | 0.03 | 65 | 0.06 | 60 | 0.05 | 5 | 0.07 |  |  | ✓ |
| K496 Pseudopleural, pleural callus excision | 95 | 0.03 | 87 | 0.03 | 8 | 0.03 | 40 | 0.03 | 38 | 0.03 | 2 | 0.03 |  |  | ✓ |
| K502 vertical septal tumor and thymus removal | 76 | 0.02 | 71 | 0.02 | 5 | 0.02 | 13 | 0.01 | 13 | 0.01 | 0 | 0 |  |  | ✓ |
| K504 Vertical septal cancer surgery | 90 | 0.03 | 89 | 0.03 | 1 | 0 | 33 | 0.03 | 32 | 0.03 | 1 | 0.01 |  |  | ✓ |
| K509 Bronchial foreign body removal | 26 | 0.01 | 24 | 0.01 | 2 | 0.01 | 12 | 0.01 | 11 | 0.01 | 1 | 0.01 |  |  | ✓ |
| K511 Pneumonectomy | 33 | 0.01 | 31 | 0.01 | 2 | 0.01 | 19 | 0.02 | 17 | 0.02 | 2 | 0.03 |  |  | ✓ |
| K513 Thoracoscopic pneumonectomy | 1069 | 0.31 | 1029 | 0.33 | 40 | 0.17 | 353 | 0.3 | 340 | 0.31 | 13 | 0.17 |  |  | ✓ |
| K514 Pulmonary cancer surgery | 3045 | 0.9 | 2956 | 0.93 | 89 | 0.39 | 1147 | 0.97 | 1106 | 1 | 41 | 0.55 |  |  | ✓ |
| K522 Esophageal stenosis extension | 452 | 0.13 | 371 | 0.12 | 81 | 0.35 | 149 | 0.13 | 128 | 0.12 | 21 | 0.28 |  |  | ✓ |
| K526 Esophageal tumor Extraction | 574 | 0.17 | 567 | 0.18 | 7 | 0.03 | 248 | 0.21 | 244 | 0.22 | 4 | 0.05 |  |  | ✓ |
| K529 Surgery for oesophageal cancer(combined with reconstruction of the alimentary canal) | 365 | 0.11 | 337 | 0.11 | 28 | 0.12 | 129 | 0.11 | 120 | 0.11 | 9 | 0.12 |  |  | ✓ |
| K533 Sclerotherapy for esophageal and gastric varices (endoscopic) (as a series) | 892 | 0.26 | 832 | 0.26 | 60 | 0.26 | 296 | 0.25 | 276 | 0.25 | 20 | 0.27 |  |  | ✓ |
| K537 Hiatal hiatal hernia surgery | 31 | 0.01 | 28 | 0.01 | 3 | 0.01 | 12 | 0.01 | 11 | 0.01 | 1 | 0.01 |  |  | ✓ |
| K546 Percutaneous coronary angioplasty | 1845 | 0.54 | 1765 | 0.56 | 80 | 0.35 | 674 | 0.57 | 648 | 0.59 | 26 | 0.35 |  |  | ✓ |
| K547 Percutaneous coronary atherectomy | 82 | 0.02 | 80 | 0.03 | 2 | 0.01 | 64 | 0.05 | 62 | 0.06 | 2 | 0.03 |  |  | ✓ |
| K548 Percutaneous coronary angioplasty (with special catheter) | 362 | 0.11 | 338 | 0.11 | 24 | 0.1 | 139 | 0.12 | 132 | 0.12 | 7 | 0.09 |  |  | ✓ |
| K549 Percutaneous coronary stenting | 7913 | 2.33 | 7626 | 2.41 | 287 | 1.24 | 2635 | 2.23 | 2550 | 2.31 | 85 | 1.13 |  | ✓ | ✓ |
| K550 Intracoronary thrombolysis | 384 | 0.11 | 372 | 0.12 | 12 | 0.05 | 107 | 0.09 | 104 | 0.09 | 3 | 0.04 |  |  | ✓ |
| K552 Coronary artery and aortic bypass grafting | 898 | 0.26 | 853 | 0.27 | 45 | 0.19 | 262 | 0.22 | 250 | 0.23 | 12 | 0.16 |  |  | ✓ |
| K553 Resection of ventricular aneurysm (including resection of infarction) | 26 | 0.01 | 25 | 0.01 | 1 | 0 | 14 | 0.01 | 13 | 0.01 | 1 | 0.01 |  |  | ✓ |
| K554 Valvuloplasty | 360 | 0.11 | 345 | 0.11 | 15 | 0.06 | 83 | 0.07 | 78 | 0.07 | 5 | 0.07 |  |  | ✓ |
| K555 Valve replacement | 931 | 0.27 | 871 | 0.28 | 60 | 0.26 | 375 | 0.32 | 357 | 0.32 | 18 | 0.24 |  |  | ✓ |
| K556 Aortic valve stenosis direct incision | 38 | 0.01 | 37 | 0.01 | 1 | 0 | 19 | 0.02 | 19 | 0.02 | 0 | 0 |  |  | ✓ |
| K560 Aortic aneurysmectomy (including anastomosis or transplantation) | 827 | 0.24 | 795 | 0.25 | 32 | 0.14 | 301 | 0.25 | 293 | 0.26 | 8 | 0.11 |  |  | ✓ |
| K561 Stent graft insertion | 645 | 0.19 | 607 | 0.19 | 38 | 0.16 | 266 | 0.23 | 254 | 0.23 | 12 | 0.16 |  |  | ✓ |
| K594 Arrhythmia surgery | 270 | 0.08 | 259 | 0.08 | 11 | 0.05 | 88 | 0.07 | 83 | 0.08 | 5 | 0.07 |  |  | ✓ |
| K595 Percutaneous catheter myocardial ablation | 2979 | 0.88 | 2922 | 0.92 | 57 | 0.25 | 943 | 0.8 | 927 | 0.84 | 16 | 0.21 |  |  | ✓ |
| K596 Extracorporeal pacemaking | 891 | 0.26 | 840 | 0.27 | 51 | 0.22 | 296 | 0.25 | 273 | 0.25 | 23 | 0.31 |  |  | ✓ |
| K597 Pacemaker implantation | 2628 | 0.77 | 2528 | 0.8 | 100 | 0.43 | 881 | 0.75 | 850 | 0.77 | 31 | 0.41 |  |  | ✓ |
| K598 Biventricular pacemaker implantation | 44 | 0.01 | 42 | 0.01 | 2 | 0.01 | 15 | 0.01 | 15 | 0.01 | 0 | 0 |  |  | ✓ |
| K599 Implantable cardioverter defibrillator | 435 | 0.13 | 416 | 0.13 | 19 | 0.08 | 150 | 0.13 | 140 | 0.13 | 10 | 0.13 |  |  | ✓ |
| K600 Aortic balloon pumping (IABP) (per day) | 789 | 0.23 | 742 | 0.23 | 47 | 0.2 | 227 | 0.19 | 213 | 0.19 | 14 | 0.19 |  |  | ✓ |
| K601 Artificial heart lung (per day) | 1817 | 0.54 | 1716 | 0.54 | 101 | 0.44 | 608 | 0.51 | 580 | 0.52 | 28 | 0.37 |  |  | ✓ |
| K602 Percutaneous cardiopulmonary support (per day) | 42 | 0.01 | 42 | 0.01 | 0 | 0 | 24 | 0.02 | 22 | 0.02 | 2 | 0.03 |  |  | ✓ |
| K607 Vascular ligation | 124 | 0.04 | 114 | 0.04 | 10 | 0.04 | 51 | 0.04 | 49 | 0.04 | 2 | 0.03 |  |  | ✓ |
| K608 Arterial embolization | 229 | 0.07 | 211 | 0.07 | 18 | 0.08 | 62 | 0.05 | 56 | 0.05 | 6 | 0.08 |  |  | ✓ |
| K609 Arterial thromboendarterectomy | 372 | 0.11 | 357 | 0.11 | 15 | 0.06 | 126 | 0.11 | 120 | 0.11 | 6 | 0.08 |  |  | ✓ |
| K610 Arterioplasty, anastomosis | 1634 | 0.48 | 1516 | 0.48 | 118 | 0.51 | 507 | 0.43 | 474 | 0.43 | 33 | 0.44 |  |  | ✓ |
| K611 Placement of implantable catheter for continuous infusion of antineoplastic agent into artery, vein or abdominal cavity | 2131 | 0.63 | 1893 | 0.6 | 238 | 1.03 | 776 | 0.66 | 677 | 0.61 | 99 | 1.32 |  |  | ✓ |
| K614 Vascular grafting, bypass grafting | 448 | 0.13 | 411 | 0.13 | 37 | 0.16 | 145 | 0.12 | 137 | 0.12 | 8 | 0.11 |  |  | ✓ |
| K615 Vascular embolization (head, thoracic, intra-abdominal vessels, etc.) | 3258 | 0.96 | 3090 | 0.98 | 168 | 0.73 | 1021 | 0.86 | 973 | 0.88 | 48 | 0.64 |  |  | ✓ |
| K616 Vasodilatation and thrombectomy of extremities | 1841 | 0.54 | 1731 | 0.55 | 110 | 0.48 | 736 | 0.62 | 712 | 0.64 | 24 | 0.32 |  |  | ✓ |
| K617 Varicose vein surgery | 42 | 0.01 | 40 | 0.01 | 2 | 0.01 | 28 | 0.02 | 28 | 0.03 | 0 | 0 |  |  | ✓ |
| K618 Placement of an implantable catheter for central venous injection | 419 | 0.12 | 340 | 0.11 | 79 | 0.34 | 125 | 0.11 | 114 | 0.1 | 11 | 0.15 |  |  | ✓ |
| K620 Inferior vena cava filter | 243 | 0.07 | 228 | 0.07 | 15 | 0.06 | 47 | 0.04 | 46 | 0.04 | 1 | 0.01 |  |  | ✓ |
| K626 Lymphadenectomy | 392 | 0.12 | 370 | 0.12 | 22 | 0.1 | 102 | 0.09 | 98 | 0.09 | 4 | 0.05 |  |  | ✓ |
| K627 Lymph node group dissection | 156 | 0.05 | 149 | 0.05 | 7 | 0.03 | 42 | 0.04 | 41 | 0.04 | 1 | 0.01 |  |  | ✓ |
| K633 Hernia surgery | 841 | 0.25 | 809 | 0.26 | 32 | 0.14 | 413 | 0.35 | 404 | 0.37 | 9 | 0.12 |  |  | ✓ |
| K634 Laparoscopic Hernia surgery (both sides)(peritoneum, retroperitoneum, mesentery, omentum) | 43 | 0.01 | 41 | 0.01 | 2 | 0.01 | 178 | 0.15 | 178 | 0.16 | 0 | 0 |  |  | ✓ |
| K635 thoracic fluid and ascites filtration and concentration re-infusion method | 609 | 0.18 | 425 | 0.13 | 184 | 0.8 | 170 | 0.14 | 117 | 0.11 | 53 | 0.71 |  |  | ✓ |
| K636 Experimental open abdominal surgery | 447 | 0.13 | 418 | 0.13 | 29 | 0.13 | 164 | 0.14 | 156 | 0.14 | 8 | 0.11 |  |  | ✓ |
| K637 Limited abdominal abscess surgery | 372 | 0.11 | 339 | 0.11 | 33 | 0.14 | 128 | 0.11 | 116 | 0.1 | 12 | 0.16 |  |  | ✓ |
| K639 Acute generalized peritonitis surgery | 384 | 0.11 | 359 | 0.11 | 25 | 0.11 | 135 | 0.11 | 124 | 0.11 | 11 | 0.15 |  |  | ✓ |
| K642 Large mesh, mesenteric and retroperitoneal tumor extraction | 116 | 0.03 | 111 | 0.04 | 5 | 0.02 | 29 | 0.02 | 28 | 0.03 | 1 | 0.01 |  |  | ✓ |
| K643 Retroperitoneal cancer surgery | 90 | 0.03 | 87 | 0.03 | 3 | 0.01 | 36 | 0.03 | 33 | 0.03 | 3 | 0.04 |  |  | ✓ |
| K647 Gastric suturing (including large mesh filling or cover operation ) | 231 | 0.07 | 218 | 0.07 | 13 | 0.06 | 65 | 0.06 | 61 | 0.06 | 4 | 0.05 |  |  | ✓ |
| K651 Endoscopic gastric and duodenal stenting | 191 | 0.06 | 136 | 0.04 | 55 | 0.24 | 71 | 0.06 | 50 | 0.05 | 21 | 0.28 |  |  | ✓ |
| K653 Endoscopic gastroduodenal plication and mucosal resection | 3031 | 0.89 | 2928 | 0.93 | 103 | 0.45 | 1045 | 0.89 | 1013 | 0.92 | 32 | 0.43 |  |  | ✓ |
| K654 Endoscopic hemostasis of the alimentary canal | 3357 | 0.99 | 3163 | 1 | 194 | 0.84 | 1096 | 0.93 | 1034 | 0.94 | 62 | 0.83 |  |  | ✓ |
| K655 Gastrectomy | 2072 | 0.61 | 1969 | 0.62 | 103 | 0.45 | 646 | 0.55 | 618 | 0.56 | 28 | 0.37 |  |  | ✓ |
| K657 Total gastric resection | 949 | 0.28 | 860 | 0.27 | 89 | 0.39 | 223 | 0.19 | 212 | 0.19 | 11 | 0.15 |  |  | ✓ |
| K662 Gastrointestinal anastomosis (Blanco-anastomosis included.) | 215 | 0.06 | 176 | 0.06 | 39 | 0.17 | 78 | 0.07 | 68 | 0.06 | 10 | 0.13 |  |  | ✓ |
| K664 Gastrostomy (including endoscopic gastrostomy and laparoscopic gastrostomy) | 846 | 0.25 | 743 | 0.23 | 103 | 0.45 | 299 | 0.25 | 273 | 0.25 | 26 | 0.35 |  |  | ✓ |
| K671 Choledochotomy for stone extraction (with insertion of chambers) | 175 | 0.05 | 166 | 0.05 | 9 | 0.04 | 58 | 0.05 | 54 | 0.05 | 4 | 0.05 |  |  | ✓ |
| K672 Choledochotomy | 6919 | 2.04 | 6743 | 2.13 | 176 | 0.76 | 2328 | 1.97 | 2278 | 2.06 | 50 | 0.67 |  | ✓ | ✓ |
| K675 Surgery for biliary cystic cancer | 88 | 0.03 | 87 | 0.03 | 1 | 0 | 50 | 0.04 | 46 | 0.04 | 4 | 0.05 |  |  | ✓ |
| K677 Surgery for bile duct cancer | 81 | 0.02 | 74 | 0.02 | 7 | 0.03 | 49 | 0.04 | 44 | 0.04 | 5 | 0.07 |  |  | ✓ |
| K680 General bile duct gastrointestinal anastomosis | 85 | 0.03 | 74 | 0.02 | 11 | 0.05 | 33 | 0.03 | 29 | 0.03 | 4 | 0.05 |  |  | ✓ |
| K681 External gallbladder fistula construction | 448 | 0.13 | 379 | 0.12 | 69 | 0.3 | 134 | 0.11 | 122 | 0.11 | 12 | 0.16 |  |  | ✓ |
| K682 External biliary fistula construction | 1486 | 0.44 | 1243 | 0.39 | 243 | 1.05 | 503 | 0.43 | 431 | 0.39 | 72 | 0.96 |  |  | ✓ |
| K685 Endoscopic biliary stone removal | 1342 | 0.4 | 1239 | 0.39 | 103 | 0.45 | 518 | 0.44 | 486 | 0.44 | 32 | 0.43 |  |  | ✓ |
| K686 Endoscopic biliary tract extension | 286 | 0.08 | 264 | 0.08 | 22 | 0.1 | 168 | 0.14 | 157 | 0.14 | 11 | 0.15 |  |  | ✓ |
| K687 Endoscopic papillotomy | 3529 | 1.04 | 3303 | 1.04 | 226 | 0.98 | 1195 | 1.01 | 1123 | 1.02 | 72 | 0.96 |  | ✓ | ✓ |
| K688 Endoscopic biliary stenting | 4695 | 1.38 | 3830 | 1.21 | 865 | 3.74 | 1699 | 1.44 | 1418 | 1.28 | 281 | 3.74 |  | ✓ | ✓ |
| K689 Transcutaneous transhepatic insertion of bile ducts | 75 | 0.02 | 53 | 0.02 | 22 | 0.1 | 30 | 0.03 | 22 | 0.02 | 8 | 0.11 |  |  | ✓ |
| K691 Hepatic abscess dissection | 265 | 0.08 | 246 | 0.08 | 19 | 0.08 | 92 | 0.08 | 76 | 0.07 | 16 | 0.21 |  |  | ✓ |
| K692 Hepatocystectomy or suture | 26 | 0.01 | 26 | 0.01 | 0 | 0 | 18 | 0.02 | 17 | 0.02 | 1 | 0.01 |  |  | ✓ |
| K695 Hepatectomy | 1119 | 0.33 | 1074 | 0.34 | 45 | 0.19 | 362 | 0.31 | 343 | 0.31 | 19 | 0.25 |  |  | ✓ |
| K697 Intrahepatic fistula construction | 931 | 0.27 | 909 | 0.29 | 22 | 0.1 | 279 | 0.24 | 270 | 0.24 | 9 | 0.12 |  |  | ✓ |
| K702 Caudal pancreatectomy | 339 | 0.1 | 315 | 0.1 | 24 | 0.1 | 115 | 0.1 | 109 | 0.1 | 6 | 0.08 |  |  | ✓ |
| K703 Cephalocele resection | 712 | 0.21 | 650 | 0.21 | 62 | 0.27 | 257 | 0.22 | 235 | 0.21 | 22 | 0.29 |  |  | ✓ |
| K704 Total pancreatectomy | 43 | 0.01 | 41 | 0.01 | 2 | 0.01 | 23 | 0.02 | 21 | 0.02 | 2 | 0.03 |  |  | ✓ |
| K708 External pancreas fistula construction | 521 | 0.15 | 470 | 0.15 | 51 | 0.22 | 198 | 0.17 | 183 | 0.17 | 15 | 0.2 |  |  | ✓ |
| K711 Splenectomy | 198 | 0.06 | 181 | 0.06 | 17 | 0.07 | 40 | 0.03 | 38 | 0.03 | 2 | 0.03 |  |  | ✓ |
| K714 Intestinal tube adhesion surgery | 724 | 0.21 | 660 | 0.21 | 64 | 0.28 | 263 | 0.22 | 248 | 0.22 | 15 | 0.2 |  |  | ✓ |
| K715 Intestinal hyperplasia revision | 263 | 0.08 | 234 | 0.07 | 29 | 0.13 | 57 | 0.05 | 53 | 0.05 | 4 | 0.05 |  |  | ✓ |
| K716 Small bowel resection | 786 | 0.23 | 734 | 0.23 | 52 | 0.23 | 244 | 0.21 | 227 | 0.21 | 17 | 0.23 |  |  | ✓ |
| K718 appendectomy | 3143 | 0.93 | 3073 | 0.97 | 70 | 0.3 | 943 | 0.8 | 931 | 0.84 | 12 | 0.16 |  |  | ✓ |
| K719 Colpotomy | 4308 | 1.27 | 4123 | 1.3 | 185 | 0.8 | 1542 | 1.31 | 1483 | 1.34 | 59 | 0.79 |  | ✓ | ✓ |
| K721 Endoscopic colorectal polyp and mucosal resection | 2096 | 0.62 | 2026 | 0.64 | 70 | 0.3 | 1332 | 1.13 | 1286 | 1.16 | 46 | 0.61 |  |  | ✓ |
| K722 Endoscopic hemostasis of small intestine | 825 | 0.24 | 768 | 0.24 | 57 | 0.25 | 309 | 0.26 | 287 | 0.26 | 22 | 0.29 |  |  | ✓ |
| K724 Intestinal anastomosis | 82 | 0.02 | 62 | 0.02 | 20 | 0.09 | 39 | 0.03 | 34 | 0.03 | 5 | 0.07 |  |  | ✓ |
| K725 Bowel fistula construction, appendicostomy | 132 | 0.04 | 116 | 0.04 | 16 | 0.07 | 50 | 0.04 | 46 | 0.04 | 4 | 0.05 |  |  | ✓ |
| K726 Colostomy | 590 | 0.17 | 530 | 0.17 | 60 | 0.26 | 223 | 0.19 | 192 | 0.17 | 31 | 0.41 |  |  | ✓ |
| K732 Artificial anus Closure | 519 | 0.15 | 504 | 0.16 | 15 | 0.06 | 142 | 0.12 | 135 | 0.12 | 7 | 0.09 |  |  | ✓ |
| K735 Congenital megacolon surgery | 476 | 0.14 | 429 | 0.14 | 47 | 0.2 | 197 | 0.17 | 181 | 0.16 | 16 | 0.21 |  |  | ✓ |
| K739 Removal of rectal tumor(Polyp extraction included.) | 66 | 0.02 | 65 | 0.02 | 1 | 0 | 19 | 0.02 | 17 | 0.02 | 2 | 0.03 |  |  | ✓ |
| K740 Rectal resection and dissection | 2102 | 0.62 | 2019 | 0.64 | 83 | 0.36 | 679 | 0.58 | 663 | 0.6 | 16 | 0.21 |  |  | ✓ |
| K742 Rectal detachment Surgery | 94 | 0.03 | 88 | 0.03 | 6 | 0.03 | 33 | 0.03 | 31 | 0.03 | 2 | 0.03 |  |  | ✓ |
| K743 Hemorrhoid surgery (Prolapse included.) | 113 | 0.03 | 106 | 0.03 | 7 | 0.03 | 36 | 0.03 | 33 | 0.03 | 3 | 0.04 |  |  | ✓ |
| K745 Perianal abscess Incision | 119 | 0.04 | 117 | 0.04 | 2 | 0.01 | 34 | 0.03 | 33 | 0.03 | 1 | 0.01 |  |  | ✓ |
| K746 Hemorrhoid fistula Radical surgery | 57 | 0.02 | 56 | 0.02 | 1 | 0 | 15 | 0.01 | 15 | 0.01 | 0 | 0 |  |  | ✓ |
| K754 Paranephrectomy (Partial nephrectomy included.) | 59 | 0.02 | 57 | 0.02 | 2 | 0.01 | 15 | 0.01 | 15 | 0.01 | 0 | 0 |  |  | ✓ |
| K764 Percutaneous urinary tract stone removal (Percutaneous fistulostomy included.) | 61 | 0.02 | 60 | 0.02 | 1 | 0 | 14 | 0.01 | 12 | 0.01 | 2 | 0.03 |  |  | ✓ |
| K768 Excavation of renal and ureteral stones by extracorporeal impulse wave (per one series) | 181 | 0.05 | 170 | 0.05 | 11 | 0.05 | 34 | 0.03 | 33 | 0.03 | 1 | 0.01 |  |  | ✓ |
| K772 renal extraction | 46 | 0.01 | 44 | 0.01 | 2 | 0.01 | 27 | 0.02 | 27 | 0.02 | 0 | 0 |  |  | ✓ |
| K773 Renal (ureteral) ulcer surgery | 1053 | 0.31 | 1030 | 0.33 | 23 | 0.1 | 377 | 0.32 | 361 | 0.33 | 16 | 0.21 |  |  | ✓ |
| K775 Transcutaneous renal (pelvic) fistulotomy | 413 | 0.12 | 335 | 0.11 | 78 | 0.34 | 138 | 0.12 | 114 | 0.1 | 24 | 0.32 |  |  | ✓ |
| K781 Urethral stone removal via urethra | 635 | 0.19 | 618 | 0.2 | 17 | 0.07 | 361 | 0.31 | 355 | 0.32 | 6 | 0.08 |  |  | ✓ |
| K783 Urethral stricture extension via urethra | 1600 | 0.47 | 1421 | 0.45 | 179 | 0.77 | 759 | 0.64 | 676 | 0.61 | 83 | 1.11 |  |  | ✓ |
| K797 Removal of intravesical coagulation | 86 | 0.03 | 80 | 0.03 | 6 | 0.03 | 37 | 0.03 | 29 | 0.03 | 8 | 0.11 |  |  | ✓ |
| K798 Removal of bladder stones and foreign bodies | 193 | 0.06 | 189 | 0.06 | 4 | 0.02 | 104 | 0.09 | 97 | 0.09 | 7 | 0.09 |  |  | ✓ |
| K800 Bladder diverticulectomy | 180 | 0.05 | 162 | 0.05 | 18 | 0.08 | 131 | 0.11 | 122 | 0.11 | 9 | 0.12 |  |  | ✓ |
| K802 Removal of bladder ulcer | 104 | 0.03 | 103 | 0.03 | 1 | 0 | 28 | 0.02 | 26 | 0.02 | 2 | 0.03 |  |  | ✓ |
| K803 Bladder ulcer surgery | 2636 | 0.78 | 2542 | 0.8 | 94 | 0.41 | 919 | 0.78 | 889 | 0.8 | 30 | 0.4 |  |  | ✓ |
| K804 Ureteral tube extraction | 28 | 0.01 | 28 | 0.01 | 0 | 0 | 12 | 0.01 | 11 | 0.01 | 1 | 0.01 |  |  | ✓ |
| K805 Cystostomy | 84 | 0.02 | 74 | 0.02 | 10 | 0.04 | 27 | 0.02 | 24 | 0.02 | 3 | 0.04 |  |  | ✓ |
| K821 Endoscopic urethral stricture surgery | 72 | 0.02 | 68 | 0.02 | 4 | 0.02 | 41 | 0.03 | 37 | 0.03 | 4 | 0.05 |  |  | ✓ |
| K828 Circumcision Surgery | 27 | 0.01 | 26 | 0.01 | 1 | 0 | 13 | 0.01 | 11 | 0.01 | 2 | 0.03 |  |  | ✓ |
| K830 Spermatophore extraction | 105 | 0.03 | 105 | 0.03 | 0 | 0 | 33 | 0.03 | 31 | 0.03 | 2 | 0.03 |  |  | ✓ |
| K833 Spermatogonial ulcer surgery | 110 | 0.03 | 110 | 0.03 | 0 | 0 | 36 | 0.03 | 36 | 0.03 | 0 | 0 |  |  | ✓ |
| K835 Vaginal Vesicle Oedema Surgery | 67 | 0.02 | 64 | 0.02 | 3 | 0.01 | 24 | 0.02 | 24 | 0.02 | 0 | 0 |  |  | ✓ |
| K838 Spermatic cord twisting surgery | 39 | 0.01 | 39 | 0.01 | 0 | 0 | 13 | 0.01 | 13 | 0.01 | 0 | 0 |  |  | ✓ |
| K841 Prostate gland surgery via urethra | 619 | 0.18 | 605 | 0.19 | 14 | 0.06 | 265 | 0.22 | 258 | 0.23 | 7 | 0.09 |  |  | ✓ |
| K843 Surgery for prostate cancer | 1245 | 0.37 | 1220 | 0.39 | 25 | 0.11 | 534 | 0.45 | 526 | 0.48 | 8 | 0.11 |  |  | ✓ |
| K860 Vaginal wall Formation surgery | 36 | 0.01 | 34 | 0.01 | 2 | 0.01 | 13 | 0.01 | 13 | 0.01 | 0 | 0 |  |  | ✓ |
| K861 Endometrial curettage | 117 | 0.03 | 116 | 0.04 | 1 | 0 | 23 | 0.02 | 22 | 0.02 | 1 | 0.01 |  |  | ✓ |
| K863 Laparoscopic Endometriosis removal | 25 | 0.01 | 25 | 0.01 | 0 | 0 | 12 | 0.01 | 12 | 0.01 | 0 | 0 |  |  | ✓ |
| K865 Uterine prolapse surgery | 458 | 0.13 | 450 | 0.14 | 8 | 0.03 | 161 | 0.14 | 160 | 0.14 | 1 | 0.01 |  |  | ✓ |
| K867 Cervical Excision | 25 | 0.01 | 25 | 0.01 | 0 | 0 | 55 | 0.05 | 53 | 0.05 | 2 | 0.03 |  |  | ✓ |
| K872 Removal of uterine polyp (nucleus pulposus) | 742 | 0.22 | 736 | 0.23 | 6 | 0.03 | 279 | 0.24 | 279 | 0.25 | 0 | 0 |  |  | ✓ |
| K877 Total uterine extraction | 2892 | 0.85 | 2834 | 0.9 | 58 | 0.25 | 1155 | 0.98 | 1138 | 1.03 | 17 | 0.23 |  |  | ✓ |
| K879 Uterine cancer ulcer surgery | 1291 | 0.38 | 1236 | 0.39 | 55 | 0.24 | 463 | 0.39 | 447 | 0.4 | 16 | 0.21 |  |  | ✓ |
| K886 Uterine appendage adhesion excision (both sides) | 71 | 0.02 | 70 | 0.02 | 1 | 0 | 41 | 0.03 | 41 | 0.04 | 0 | 0 |  |  | ✓ |
| K888 Extraction of uterine appendage ulcer (both sides) | 3476 | 1.02 | 3429 | 1.08 | 47 | 0.2 | 1187 | 1.01 | 1177 | 1.06 | 10 | 0.13 |  | ✓ | ✓ |
| K889 Surgery for malignant ulcer of uterine appendages (both sides) | 628 | 0.18 | 609 | 0.19 | 19 | 0.08 | 232 | 0.2 | 226 | 0.2 | 6 | 0.08 |  |  | ✓ |
| K920 Blood transfusion | 27705 | 8.16 | 24458 | 7.73 | 3247 | 14.05 | 8915 | 7.55 | 7969 | 7.21 | 946 | 12.6 | ✓ | ✓ | ✓ |
| K921 Hematopoietic stem cell collection (per series) | 148 | 0.04 | 146 | 0.05 | 2 | 0.01 | 28 | 0.02 | 27 | 0.02 | 1 | 0.01 |  |  | ✓ |
| K923 Intraoperative autologous blood collection (with autologous blood collection device) | 2976 | 0.88 | 2838 | 0.9 | 138 | 0.6 | 986 | 0.84 | 948 | 0.86 | 38 | 0.51 |  |  | ✓ |
| K930 Additional charge for Spinal cord induced potential measurement and other | 1217 | 0.36 | 1182 | 0.37 | 35 | 0.15 | 589 | 0.5 | 580 | 0.52 | 9 | 0.12 |  |  | ✓ |
| K931 Additional charge for ultrasonic coagulation and incision device | 27062 | 7.97 | 26072 | 8.24 | 990 | 4.28 | 9860 | 8.35 | 9542 | 8.63 | 318 | 4.24 | ✓ | ✓ | ✓ |
| K932 Additional charge for external fixator | 118 | 0.03 | 116 | 0.04 | 2 | 0.01 | 20 | 0.02 | 19 | 0.02 | 1 | 0.01 |  |  | ✓ |
| K933 Additional charge for the Iontoforehead | 148 | 0.04 | 140 | 0.04 | 8 | 0.03 | 26 | 0.02 | 25 | 0.02 | 1 | 0.01 |  |  | ✓ |
| K934 Additional charge for endoscope for paranasal surgery | 185 | 0.05 | 182 | 0.06 | 3 | 0.01 | 63 | 0.05 | 62 | 0.06 | 1 | 0.01 |  |  | ✓ |
| K935 Additional charge for Heating coagulation incision device for hemostasis | 499 | 0.15 | 492 | 0.16 | 7 | 0.03 | 142 | 0.12 | 142 | 0.13 | 0 | 0 |  |  | ✓ |
| K936 Additional charge foe Automatic Suture Clutch | 15040 | 4.43 | 14323 | 4.53 | 717 | 3.1 | 5318 | 4.5 | 5098 | 4.61 | 220 | 2.93 |  | ✓ | ✓ |
| K937 Additional charge for the machine for coronary artery and aorta bypass graft under heart beat | 661 | 0.19 | 633 | 0.2 | 28 | 0.12 | 249 | 0.21 | 238 | 0.22 | 11 | 0.15 |  |  | ✓ |
| K938 Additional charge for external impulse wave consuming electrodes | 161 | 0.05 | 153 | 0.05 | 8 | 0.03 | 24 | 0.02 | 23 | 0.02 | 1 | 0.01 |  |  | ✓ |
| K939 Additional charge for Surgical support such as imaging | 3833 | 1.13 | 3673 | 1.16 | 160 | 0.69 | 1786 | 1.51 | 1722 | 1.56 | 64 | 0.85 |  | ✓ | ✓ |
