## Supplementary Table S3 for "Comparison of machine-learning and logistic regression models to predict 30-day unplanned readmission: a development and validation study"

| **Supplementary Table S3. Details and frequency of procedure codes (Japanese original codes)** | | | | | | | | | | | | | | | |
| --- | --- | --- | --- | --- | --- | --- | --- | --- | --- | --- | --- | --- | --- | --- | --- |
| Variables | Derivation dataset (N=339,513) | | | | | | Validation dataset (N=118,074) | | | | | | Use of the variable | | |
|  | Total  N=339,513 | | Outcome (-)  N=316,405 | | Outcome (+)  N=23,108 | | Total  N=118,074 | | Outcome (-)  N=110,567 | | Outcome (+)  N=7,507 | | Data pattern  1&2 | Data pattern  3&4 | Data pattern  5&6 |
|  | n | % | n | % | n | % | n | % | n | % | n | % |  |  |  |
| J000 Wound care | 67913 | 20 | 64901 | 20.51 | 3012 | 13.03 | 23943 | 20.28 | 22836 | 20.65 | 1107 | 14.75 | ✓ | ✓ | ✓ |
| J001 Burn treatment | 1004 | 0.3 | 929 | 0.29 | 75 | 0.32 | 323 | 0.27 | 306 | 0.28 | 17 | 0.23 |  |  | ✓ |
| J002 Drainage | 53902 | 15.88 | 50875 | 16.08 | 3027 | 13.1 | 17978 | 15.23 | 17097 | 15.46 | 881 | 11.74 | ✓ | ✓ | ✓ |
| J003 Local negative pressure closure procedure (inpatient) | 683 | 0.2 | 646 | 0.2 | 37 | 0.16 | 234 | 0.2 | 221 | 0.2 | 13 | 0.17 |  |  | ✓ |
| J007 Cervical, thoracic, or lumbar puncture | 263 | 0.08 | 247 | 0.08 | 16 | 0.07 | 68 | 0.06 | 65 | 0.06 | 3 | 0.04 |  |  | ✓ |
| J008 Thoracentesis (including washing, injection and drainage) | 1824 | 0.54 | 1545 | 0.49 | 279 | 1.21 | 668 | 0.57 | 564 | 0.51 | 104 | 1.39 |  |  | ✓ |
| J010 Abdominocentesis (including artificial insufflation, washing, infusion, and drainage) | 1201 | 0.35 | 860 | 0.27 | 341 | 1.48 | 412 | 0.35 | 319 | 0.29 | 93 | 1.24 |  |  | ✓ |
| J011 Bone marrow puncture | 113 | 0.03 | 102 | 0.03 | 11 | 0.05 | 111 | 0.09 | 95 | 0.09 | 16 | 0.21 |  |  | ✓ |
| J016 Lymph node puncture | 29 | 0.01 | 28 | 0.01 | 1 | 0 | 11 | 0.01 | 11 | 0.01 | 0 | 0 |  |  | ✓ |
| J017 Local injection of ethanol | 67 | 0.02 | 65 | 0.02 | 2 | 0.01 | 14 | 0.01 | 14 | 0.01 | 0 | 0 |  |  | ✓ |
| J018 Sputum suction (per day) | 19883 | 5.86 | 17689 | 5.59 | 2194 | 9.49 | 7446 | 6.31 | 6667 | 6.03 | 779 | 10.38 | ✓ | ✓ | ✓ |
| J019 Continuous thoracic drainage (first day) | 3080 | 0.91 | 2744 | 0.87 | 336 | 1.45 | 1019 | 0.86 | 906 | 0.82 | 113 | 1.51 |  |  | ✓ |
| J020 Continuous gastric drainage (first day) | 5155 | 1.52 | 4632 | 1.46 | 523 | 2.26 | 1484 | 1.26 | 1355 | 1.23 | 129 | 1.72 |  | ✓ | ✓ |
| J021 Continuous abdominal drainage (first day) | 279 | 0.08 | 222 | 0.07 | 57 | 0.25 | 81 | 0.07 | 73 | 0.07 | 8 | 0.11 |  |  | ✓ |
| J022 Elevated enema, high-pressure enema, bowel wash | 14204 | 4.18 | 12556 | 3.97 | 1648 | 7.13 | 5172 | 4.38 | 4681 | 4.23 | 491 | 6.54 |  |  | ✓ |
| J024 Oxygen inhalation (per day) | 94767 | 27.91 | 87066 | 27.52 | 7701 | 33.33 | 32164 | 27.24 | 29629 | 26.8 | 2535 | 33.77 | ✓ | ✓ | ✓ |
| J026 Intermittent positive pressure inhalation (per day) | 1188 | 0.35 | 1041 | 0.33 | 147 | 0.64 | 663 | 0.56 | 601 | 0.54 | 62 | 0.83 |  |  | ✓ |
| J027 Hyperbaric oxygen therapy (per day) | 410 | 0.12 | 394 | 0.12 | 16 | 0.07 | 113 | 0.1 | 110 | 0.1 | 3 | 0.04 |  |  | ✓ |
| J034 Long tube insertion for ileus | 2312 | 0.68 | 2108 | 0.67 | 204 | 0.88 | 838 | 0.71 | 748 | 0.68 | 90 | 1.2 |  |  | ✓ |
| J036 Manual repair of irreducible hernia | 130 | 0.04 | 119 | 0.04 | 11 | 0.05 | 52 | 0.04 | 50 | 0.05 | 2 | 0.03 |  |  | ✓ |
| J038 Artificial kidney (per day) | 8200 | 2.42 | 7496 | 2.37 | 704 | 3.05 | 2858 | 2.42 | 2598 | 2.35 | 260 | 3.46 |  | ✓ | ✓ |
| J039 Plasma exchange (per day) | 104 | 0.03 | 94 | 0.03 | 10 | 0.04 | 49 | 0.04 | 47 | 0.04 | 2 | 0.03 |  |  | ✓ |
| J040 Regional perfusion (per day) | 12 | 0 | 11 | 0 | 1 | 0 | 16 | 0.01 | 15 | 0.01 | 1 | 0.01 |  |  | ✓ |
| J041 Adsorptive hemodiafiltration (per day) | 239 | 0.07 | 225 | 0.07 | 14 | 0.06 | 59 | 0.05 | 55 | 0.05 | 4 | 0.05 |  |  | ✓ |
| J042 Peritoneal perfusion (per day) | 337 | 0.1 | 288 | 0.09 | 49 | 0.21 | 154 | 0.13 | 130 | 0.12 | 24 | 0.32 |  |  | ✓ |
| J043 Phototherapy for neonatal hyperbilirubinemia (per day) | 1095 | 0.32 | 922 | 0.29 | 173 | 0.75 | 342 | 0.29 | 298 | 0.27 | 44 | 0.59 |  |  | ✓ |
| J044 Endotracheal intubation for life-saving | 1071 | 0.32 | 988 | 0.31 | 83 | 0.36 | 321 | 0.27 | 294 | 0.27 | 27 | 0.36 |  |  | ✓ |
| J045 Artificial respiration | 6902 | 2.03 | 6155 | 1.95 | 747 | 3.23 | 2282 | 1.93 | 2063 | 1.87 | 219 | 2.92 |  | ✓ | ✓ |
| J046 closed chest cardiac massage | 193 | 0.06 | 173 | 0.05 | 20 | 0.09 | 59 | 0.05 | 57 | 0.05 | 2 | 0.03 |  |  | ✓ |
| J047 Countershock (per day) | 908 | 0.27 | 857 | 0.27 | 51 | 0.22 | 250 | 0.21 | 230 | 0.21 | 20 | 0.27 |  |  | ✓ |
| J048 Pericardiocentesis | 124 | 0.04 | 111 | 0.04 | 13 | 0.06 | 52 | 0.04 | 49 | 0.04 | 3 | 0.04 |  |  | ✓ |
| J051 Gastric lavage | 201 | 0.06 | 183 | 0.06 | 18 | 0.08 | 39 | 0.03 | 38 | 0.03 | 1 | 0.01 |  |  | ✓ |
| J053 Dermatological ointment procedures | 2593 | 0.76 | 2348 | 0.74 | 245 | 1.06 | 1316 | 1.11 | 1194 | 1.08 | 122 | 1.63 |  |  | ✓ |
| J056 Cryocoagulation of warts | 118 | 0.03 | 107 | 0.03 | 11 | 0.05 | 39 | 0.03 | 37 | 0.03 | 2 | 0.03 |  |  | ✓ |
| J057 Excision of molluscum contagiosum | 173 | 0.05 | 165 | 0.05 | 8 | 0.03 | 71 | 0.06 | 67 | 0.06 | 4 | 0.05 |  |  | ✓ |
| J059 Scrotal edema puncture | 128 | 0.04 | 117 | 0.04 | 11 | 0.05 | 34 | 0.03 | 33 | 0.03 | 1 | 0.01 |  |  | ✓ |
| J060 Cystourethral lavage (per day) | 2741 | 0.81 | 2476 | 0.78 | 265 | 1.15 | 1084 | 0.92 | 982 | 0.89 | 102 | 1.36 |  |  | ✓ |
| J061 Perineal pelvic lavage (unilateral) | 343 | 0.1 | 276 | 0.09 | 67 | 0.29 | 78 | 0.07 | 68 | 0.06 | 10 | 0.13 |  |  | ✓ |
| J063 Placement of indwelling catheter | 34223 | 10.08 | 30879 | 9.76 | 3344 | 14.47 | 12319 | 10.43 | 11212 | 10.14 | 1107 | 14.75 | ✓ | ✓ | ✓ |
| J064 Urinary drainage (requiring urethral dilatation) | 6021 | 1.77 | 5451 | 1.72 | 570 | 2.47 | 2439 | 2.07 | 2226 | 2.01 | 213 | 2.84 |  | ✓ | ✓ |
| J065 Intermittent urinary drainage (per day) | 434 | 0.13 | 398 | 0.13 | 36 | 0.16 | 254 | 0.22 | 238 | 0.22 | 16 | 0.21 |  |  | ✓ |
| J066 Urethral dilatation | 57 | 0.02 | 53 | 0.02 | 4 | 0.02 | 20 | 0.02 | 17 | 0.02 | 3 | 0.04 |  |  | ✓ |
| J073 Uterine cavity lavage (including drug infusion) | 119 | 0.04 | 117 | 0.04 | 2 | 0.01 | 53 | 0.04 | 53 | 0.05 | 0 | 0 |  |  | ✓ |
| J080 Cervical dilation and delivery induction | 56 | 0.02 | 55 | 0.02 | 1 | 0 | 20 | 0.02 | 20 | 0.02 | 0 | 0 |  |  | ✓ |
| J082 Noninvasive repair of uterine prolapse (pessary) | 48 | 0.01 | 44 | 0.01 | 4 | 0.02 | 21 | 0.02 | 19 | 0.02 | 2 | 0.03 |  |  | ✓ |
| J089 Eyelash extraction | 99 | 0.03 | 86 | 0.03 | 13 | 0.06 | 25 | 0.02 | 23 | 0.02 | 2 | 0.03 |  |  | ✓ |
| J090 Removal of conjunctival foreign body (per eyelid) | 19 | 0.01 | 17 | 0.01 | 2 | 0.01 | 10 | 0.01 | 9 | 0.01 | 1 | 0.01 |  |  | ✓ |
| J092 Lacrimal sac bougie (including washing) | 35 | 0.01 | 32 | 0.01 | 3 | 0.01 | 12 | 0.01 | 12 | 0.01 | 0 | 0 |  |  | ✓ |
| J095 Ear treatment (including ear bath and ear cleaning) | 81 | 0.02 | 75 | 0.02 | 6 | 0.03 | 22 | 0.02 | 17 | 0.02 | 5 | 0.07 |  |  | ✓ |
| J097 Nasal procedures (including nasal aspiration, simple nose bleed, and nasal vestibular procedures) | 230 | 0.07 | 222 | 0.07 | 8 | 0.03 | 36 | 0.03 | 35 | 0.03 | 1 | 0.01 |  |  | ✓ |
| J098 Oral and pharyngeal procedures | 75 | 0.02 | 72 | 0.02 | 3 | 0.01 | 31 | 0.03 | 31 | 0.03 | 0 | 0 |  |  | ✓ |
| J100 Procedures after paranasal sinus surgery (unilateral) | 183 | 0.05 | 180 | 0.06 | 3 | 0.01 | 67 | 0.06 | 65 | 0.06 | 2 | 0.03 |  |  | ✓ |
| J103 Peritonsillar abscess puncture (including peritonsillaritis) | 145 | 0.04 | 141 | 0.04 | 4 | 0.02 | 29 | 0.02 | 28 | 0.03 | 1 | 0.01 |  |  | ✓ |
| J108 Nasal bleeding hemostasis (with gauze tampon or balloon) | 264 | 0.08 | 235 | 0.07 | 29 | 0.13 | 51 | 0.04 | 45 | 0.04 | 6 | 0.08 |  |  | ✓ |
| J113 Removal of cerumen plug (complicated) | 1179 | 0.35 | 1097 | 0.35 | 82 | 0.35 | 519 | 0.44 | 472 | 0.43 | 47 | 0.63 |  |  | ✓ |
| J115 Ultrasonic nebulizer (per day) | 16021 | 4.72 | 14946 | 4.72 | 1075 | 4.65 | 4731 | 4.01 | 4429 | 4.01 | 302 | 4.02 |  | ✓ | ✓ |
| J116 Arthrocentesis (unilateral) | 825 | 0.24 | 775 | 0.24 | 50 | 0.22 | 314 | 0.27 | 298 | 0.27 | 16 | 0.21 |  |  | ✓ |
| J117 Direct traction with steel wire, etc. (Day 2 or later. Including procedure fee in case of invasive traction) (per day for one site) | 151 | 0.04 | 147 | 0.05 | 4 | 0.02 | 42 | 0.04 | 40 | 0.04 | 2 | 0.03 |  |  | ✓ |
| J118 Intervention traction (per day) | 842 | 0.25 | 799 | 0.25 | 43 | 0.19 | 256 | 0.22 | 247 | 0.22 | 9 | 0.12 |  |  | ✓ |
| J119Anti-inflammatory and analgesic treatment (per day) | 7356 | 2.17 | 7049 | 2.23 | 307 | 1.33 | 2442 | 2.07 | 2351 | 2.13 | 91 | 1.21 |  | ✓ | ✓ |
| J120 Nasal feeding (per day) | 5615 | 1.65 | 4910 | 1.55 | 705 | 3.05 | 1825 | 1.55 | 1634 | 1.48 | 191 | 2.54 |  | ✓ | ✓ |
| J121 Nutritive enema | 65 | 0.02 | 56 | 0.02 | 9 | 0.04 | 10 | 0.01 | 10 | 0.01 | 0 | 0 |  |  | ✓ |
| J122 Limb cast bandage | 4147 | 1.22 | 4061 | 1.28 | 86 | 0.37 | 1319 | 1.12 | 1285 | 1.16 | 34 | 0.45 |  | ✓ | ✓ |
| J123 Trunk cast bandage | 422 | 0.12 | 400 | 0.13 | 22 | 0.1 | 143 | 0.12 | 132 | 0.12 | 11 | 0.15 |  |  | ✓ |
| J129 Casting of prosthetic limb | 3954 | 1.16 | 3776 | 1.19 | 178 | 0.77 | 1553 | 1.32 | 1490 | 1.35 | 63 | 0.84 |  | ✓ | ✓ |
