## Supplementary Table S4 for "Comparison of machine-learning and logistic regression models to predict 30-day unplanned readmission: a development and validation study"

| **Supplementary Table S4. Details and frequency of drug codes (Anatomical Therapeutic Chemical codes)** | | | | | | | | | | | | | | | | |
| --- | --- | --- | --- | --- | --- | --- | --- | --- | --- | --- | --- | --- | --- | --- | --- | --- |
| ATC codes  (EPHMRAversion) | Drug name | Derivation dataset (N=339,513) | | | | | | Validation dataset (N=118,074) | | | | | | Use of the variable | | |
|  |  | Total  N=339,513 | | Outcome (-)  N=316,405 | | Outcome (+)  N=23,108 | | Total  N=118,074 | | Outcome (-)  N=110,567 | | Outcome (+)  N=7,507 | | Data pattern  1&2 | Data pattern  3&4 | Data pattern  5&6 |
|  |  | n | % | n | % | n | % | n | % | n | % | n | % |  |  |  |
| A01A0 | STOMATOLOGICALS | 3299 | 0.97 | 3017 | 0.95 | 282 | 1.22 | 1118 | 0.95 | 1033 | 0.93 | 85 | 1.13 |  |  | ✓ |
| A01A2 | Mouth antiseptics and anti-infectives | 164 | 0.05 | 143 | 0.05 | 21 | 0.09 | 52 | 0.04 | 45 | 0.04 | 7 | 0.09 |  |  | ✓ |
| A01A3 | Mouth anti-inflammatories and mouth analgesics for topical use | 1436 | 0.42 | 1309 | 0.41 | 127 | 0.55 | 255 | 0.22 | 233 | 0.21 | 22 | 0.29 |  |  | ✓ |
| A01A5 | All other stomatologicals | 495 | 0.15 | 440 | 0.14 | 55 | 0.24 | 80 | 0.07 | 73 | 0.07 | 7 | 0.09 |  |  | ✓ |
| A01B0 | MOUTH ANTIFUNGALS | 447 | 0.13 | 373 | 0.12 | 74 | 0.32 | 112 | 0.09 | 98 | 0.09 | 14 | 0.19 |  |  | ✓ |
| A02A1 | Plain antacids | 81345 | 23.96 | 74039 | 23.4 | 7306 | 31.62 | 26857 | 22.75 | 24611 | 22.26 | 2246 | 29.92 | ✓ | ✓ | ✓ |
| A02A2 | Plain antiflatulents and carminatives | 34666 | 10.21 | 31845 | 10.06 | 2821 | 12.21 | 11696 | 9.91 | 10842 | 9.81 | 854 | 11.38 | ✓ | ✓ | ✓ |
| A02A6 | Antacids with other drugs | 939 | 0.28 | 831 | 0.26 | 108 | 0.47 | 171 | 0.14 | 159 | 0.14 | 12 | 0.16 |  |  | ✓ |
| A02B1 | H2 antagonists | 56134 | 16.53 | 52601 | 16.62 | 3533 | 15.29 | 15975 | 13.53 | 15032 | 13.6 | 943 | 12.56 | ✓ | ✓ | ✓ |
| A02B2 | Proton pump inhibitors | 105002 | 30.93 | 95404 | 30.15 | 9598 | 41.54 | 37506 | 31.76 | 34315 | 31.04 | 3191 | 42.51 | ✓ | ✓ | ✓ |
| A02B3 | Prostaglandin antiulcerants | 2332 | 0.69 | 2242 | 0.71 | 90 | 0.39 | 385 | 0.33 | 368 | 0.33 | 17 | 0.23 |  |  | ✓ |
| A02B9 | All other antiulcerants | 70579 | 20.79 | 66460 | 21 | 4119 | 17.82 | 22339 | 18.92 | 21147 | 19.13 | 1192 | 15.88 | ✓ | ✓ | ✓ |
| A02X0 | OTHER STOMACH DISORDER PRODUCTS | 3224 | 0.95 | 2990 | 0.94 | 234 | 1.01 | 909 | 0.77 | 852 | 0.77 | 57 | 0.76 |  |  | ✓ |
| A03A0 | PLAIN ANTISPASMODICS AND ANTICHOLINERGICS | 41854 | 12.33 | 39483 | 12.48 | 2371 | 10.26 | 13410 | 11.36 | 12763 | 11.54 | 647 | 8.62 | ✓ | ✓ | ✓ |
| A03E0 | ANTISPASMODICS COMBINED WITH OTHER PRODUCTS | 865 | 0.25 | 831 | 0.26 | 34 | 0.15 | 541 | 0.46 | 527 | 0.48 | 14 | 0.19 |  |  | ✓ |
| A03F0 | GASTROPROKINETICS | 68909 | 20.3 | 63857 | 20.18 | 5052 | 21.86 | 22499 | 19.05 | 21002 | 18.99 | 1497 | 19.94 | ✓ | ✓ | ✓ |
| A03G0 | GASTRO-INTESTINAL SENSORIMOTOR MODULATORS | 2186 | 0.64 | 1960 | 0.62 | 226 | 0.98 | 649 | 0.55 | 580 | 0.52 | 69 | 0.92 |  |  | ✓ |
| A04A1 | Serotonin antagonist antiemetics/antinauseants | 33854 | 9.97 | 31606 | 9.99 | 2248 | 9.73 | 9965 | 8.44 | 9255 | 8.37 | 710 | 9.46 | ✓ | ✓ | ✓ |
| A04A2 | NK1 antagonist antiemetics/antinauseants | 15927 | 4.69 | 14984 | 4.74 | 943 | 4.08 | 4984 | 4.22 | 4652 | 4.21 | 332 | 4.42 |  | ✓ | ✓ |
| A04A9 | Other antiemetics and antinauseants | 261 | 0.08 | 233 | 0.07 | 28 | 0.12 | 153 | 0.13 | 142 | 0.13 | 11 | 0.15 |  |  | ✓ |
| A05A2 | Bile stone therapy | 13188 | 3.88 | 11555 | 3.65 | 1633 | 7.07 | 4150 | 3.51 | 3667 | 3.32 | 483 | 6.43 |  | ✓ | ✓ |
| A05B0 | HEPATIC PROTECTORS, LIPOTROPICS | 8414 | 2.48 | 7677 | 2.43 | 737 | 3.19 | 2269 | 1.92 | 2078 | 1.88 | 191 | 2.54 |  | ✓ | ✓ |
| A06A2 | Stimulant laxatives | 109121 | 32.14 | 100662 | 31.81 | 8459 | 36.61 | 37374 | 31.65 | 34834 | 31.5 | 2540 | 33.84 | ✓ | ✓ | ✓ |
| A06A3 | Bulk-forming laxatives | 513 | 0.15 | 453 | 0.14 | 60 | 0.26 | 181 | 0.15 | 171 | 0.15 | 10 | 0.13 |  |  | ✓ |
| A06A4 | Enemas | 35225 | 10.38 | 32735 | 10.35 | 2490 | 10.78 | 12457 | 10.55 | 11704 | 10.59 | 753 | 10.03 | ✓ | ✓ | ✓ |
| A06A9 | Other drugs for constipation | 21036 | 6.2 | 18822 | 5.95 | 2214 | 9.58 | 8749 | 7.41 | 7904 | 7.15 | 845 | 11.26 | ✓ | ✓ | ✓ |
| A06B1 | Osmotic bowel cleansers | 13142 | 3.87 | 12621 | 3.99 | 521 | 2.25 | 4476 | 3.79 | 4317 | 3.9 | 159 | 2.12 |  | ✓ | ✓ |
| A06B2 | Osmotic bowel cleansers with electrolytes | 10596 | 3.12 | 9928 | 3.14 | 668 | 2.89 | 3896 | 3.3 | 3693 | 3.34 | 203 | 2.7 |  | ✓ | ✓ |
| A07A0 | INTESTINAL ANTI-INFECTIVES | 1580 | 0.47 | 1410 | 0.45 | 170 | 0.74 | 742 | 0.63 | 649 | 0.59 | 93 | 1.24 |  |  | ✓ |
| A07B0 | INTESTINAL ADSORBENT ANTIDIARRHOEALS | 96 | 0.03 | 91 | 0.03 | 5 | 0.02 | 27 | 0.02 | 27 | 0.02 | 0 | 0 |  |  | ✓ |
| A07E1 | INTESTINAL AMINOSALICYLATE PRODUCTS | 805 | 0.24 | 753 | 0.24 | 52 | 0.23 | 260 | 0.22 | 241 | 0.22 | 19 | 0.25 |  |  | ✓ |
| A07E2 | INTESTINAL CORTICOSTEROID PRODUCTS | 58 | 0.02 | 53 | 0.02 | 5 | 0.02 | 38 | 0.03 | 33 | 0.03 | 5 | 0.07 |  |  | ✓ |
| A07F0 | ANTIDIARRHOEAL MICRO-ORGANISMS | 43858 | 12.92 | 40072 | 12.66 | 3786 | 16.38 | 15067 | 12.76 | 13814 | 12.49 | 1253 | 16.69 | ✓ | ✓ | ✓ |
| A07G0 | ORAL ELECTROLYTE REPLACERS | 169 | 0.05 | 136 | 0.04 | 33 | 0.14 | 25 | 0.02 | 21 | 0.02 | 4 | 0.05 |  |  | ✓ |
| A07H0 | MOTILITY INHIBITORS | 4588 | 1.35 | 4062 | 1.28 | 526 | 2.28 | 1388 | 1.18 | 1221 | 1.1 | 167 | 2.22 |  | ✓ | ✓ |
| A07X0 | INTESTINAL DISORDER PRODUCTS, OTHER | 1273 | 0.37 | 1131 | 0.36 | 142 | 0.61 | 380 | 0.32 | 330 | 0.3 | 50 | 0.67 |  |  | ✓ |
| A09A0 | DIGESTIVES, INCLUDING ENZYMES | 5476 | 1.61 | 4839 | 1.53 | 637 | 2.76 | 1718 | 1.46 | 1548 | 1.4 | 170 | 2.26 |  | ✓ | ✓ |
| A10C1 | Human insulins and analogues, fast-acting | 33421 | 9.84 | 30630 | 9.68 | 2791 | 12.08 | 12147 | 10.29 | 11250 | 10.17 | 897 | 11.95 | ✓ | ✓ | ✓ |
| A10C2 | Human insulins and analogues, intermediate-acting | 409 | 0.12 | 374 | 0.12 | 35 | 0.15 | 131 | 0.11 | 120 | 0.11 | 11 | 0.15 |  |  | ✓ |
| A10C3 | Human insulins and analogues, intermediate- or long-acting, combined with fast-acting | 2326 | 0.69 | 2103 | 0.66 | 223 | 0.97 | 829 | 0.7 | 754 | 0.68 | 75 | 1 |  |  | ✓ |
| A10C5 | Human insulins and analogues, long-acting | 9454 | 2.78 | 8718 | 2.76 | 736 | 3.19 | 3222 | 2.73 | 2981 | 2.7 | 241 | 3.21 |  | ✓ | ✓ |
| A10H0 | SULPHONYLUREA ANTIDIABETICS | 6752 | 1.99 | 6209 | 1.96 | 543 | 2.35 | 1814 | 1.54 | 1690 | 1.53 | 124 | 1.65 |  | ✓ | ✓ |
| A10J1 | Biguanide antidiabetics, plain | 7512 | 2.21 | 7111 | 2.25 | 401 | 1.74 | 2441 | 2.07 | 2314 | 2.09 | 127 | 1.69 |  | ✓ | ✓ |
| A10K1 | Glitazone antidiabetics, plain | 1860 | 0.55 | 1760 | 0.56 | 100 | 0.43 | 499 | 0.42 | 459 | 0.42 | 40 | 0.53 |  |  | ✓ |
| A10L0 | ALPHA-GLUCOSIDASE INHIBITOR ANTIDIABETICS | 6331 | 1.86 | 5770 | 1.82 | 561 | 2.43 | 1823 | 1.54 | 1670 | 1.51 | 153 | 2.04 |  | ✓ | ✓ |
| A10M1 | Glinide antidiabetics, plain | 2604 | 0.77 | 2360 | 0.75 | 244 | 1.06 | 1021 | 0.86 | 914 | 0.83 | 107 | 1.43 |  |  | ✓ |
| A10M9 | Glinide antidiabetic combinations, other | 217 | 0.06 | 205 | 0.06 | 12 | 0.05 | 76 | 0.06 | 70 | 0.06 | 6 | 0.08 |  |  | ✓ |
| A10N1 | DPP-IV inhibitor antidiabetics, plain | 23140 | 6.82 | 21142 | 6.68 | 1998 | 8.65 | 7828 | 6.63 | 7150 | 6.47 | 678 | 9.03 | ✓ | ✓ | ✓ |
| A10N3 | DPP-IV inhibitor and biguanide antidiabetic combinations | 90 | 0.03 | 82 | 0.03 | 8 | 0.03 | 98 | 0.08 | 90 | 0.08 | 8 | 0.11 |  |  | ✓ |
| A10N9 | DPP-IV inhibitor antidiabetic combinations, other | 84 | 0.02 | 79 | 0.02 | 5 | 0.02 | 17 | 0.01 | 17 | 0.02 | 0 | 0 |  |  | ✓ |
| A10P1 | SGLT2 inhibitor antidiabetics, plain | 1263 | 0.37 | 1200 | 0.38 | 63 | 0.27 | 1132 | 0.96 | 1068 | 0.97 | 64 | 0.85 |  |  | ✓ |
| A10S0 | GLP-1 AGONIST ANTIDIABETICS | 887 | 0.26 | 846 | 0.27 | 41 | 0.18 | 436 | 0.37 | 399 | 0.36 | 37 | 0.49 |  |  | ✓ |
| A10X9 | Other drugs used in diabetes | 581 | 0.17 | 532 | 0.17 | 49 | 0.21 | 186 | 0.16 | 171 | 0.15 | 15 | 0.2 |  |  | ✓ |
| A11C2 | Vitamin D | 11910 | 3.51 | 10843 | 3.43 | 1067 | 4.62 | 4506 | 3.82 | 4142 | 3.75 | 364 | 4.85 |  | ✓ | ✓ |
| A11D3 | Vitamin B1 plain | 2254 | 0.66 | 2054 | 0.65 | 200 | 0.87 | 924 | 0.78 | 842 | 0.76 | 82 | 1.09 |  |  | ✓ |
| A11D4 | Vitamin B1 combinations with vitamin B6 and/or vitamin B12 | 38823 | 11.43 | 34855 | 11.02 | 3968 | 17.17 | 12011 | 10.17 | 10915 | 9.87 | 1096 | 14.6 | ✓ | ✓ | ✓ |
| A11D9 | Other vitamin B1 combinations | 3870 | 1.14 | 3418 | 1.08 | 452 | 1.96 | 1168 | 0.99 | 1032 | 0.93 | 136 | 1.81 |  |  | ✓ |
| A11E1 | Plain vitamin B complex | 221 | 0.07 | 198 | 0.06 | 23 | 0.1 | 88 | 0.07 | 79 | 0.07 | 9 | 0.12 |  |  | ✓ |
| A11F0 | PLAIN VITAMIN B12 | 12697 | 3.74 | 11763 | 3.72 | 934 | 4.04 | 3823 | 3.24 | 3527 | 3.19 | 296 | 3.94 |  |  | ✓ |
| A11G1 | Plain vitamin C (including vitamin C salts) | 11839 | 3.49 | 10809 | 3.42 | 1030 | 4.46 | 2837 | 2.4 | 2615 | 2.37 | 222 | 2.96 |  | ✓ | ✓ |
| A11X1 | Nicotinamide (vitamin B3), plain | 49 | 0.01 | 45 | 0.01 | 4 | 0.02 | 22 | 0.02 | 19 | 0.02 | 3 | 0.04 |  |  | ✓ |
| A11X2 | Vitamin B6 (pyridoxine), plain | 750 | 0.22 | 677 | 0.21 | 73 | 0.32 | 293 | 0.25 | 276 | 0.25 | 17 | 0.23 |  |  | ✓ |
| A11X3 | Vitamin E, plain | 116 | 0.03 | 107 | 0.03 | 9 | 0.04 | 39 | 0.03 | 36 | 0.03 | 3 | 0.04 |  |  | ✓ |
| A11X9 | All other vitamins, plain and in combination | 17102 | 5.04 | 15874 | 5.02 | 1228 | 5.31 | 5123 | 4.34 | 4794 | 4.34 | 329 | 4.38 |  | ✓ | ✓ |
| A12A0 | CALCIUM PRODUCTS | 7605 | 2.24 | 6959 | 2.2 | 646 | 2.8 | 2548 | 2.16 | 2317 | 2.1 | 231 | 3.08 |  | ✓ | ✓ |
| A12B0 | POTASSIUM PRODUCTS | 21979 | 6.47 | 19921 | 6.3 | 2058 | 8.91 | 7878 | 6.67 | 7192 | 6.5 | 686 | 9.14 | ✓ | ✓ | ✓ |
| A12C2 | Other mineral supplements | 658 | 0.19 | 564 | 0.18 | 94 | 0.41 | 390 | 0.33 | 351 | 0.32 | 39 | 0.52 |  |  | ✓ |
| A14A1 | Plain anabolic hormones, systemic | 124 | 0.04 | 102 | 0.03 | 22 | 0.1 | 37 | 0.03 | 30 | 0.03 | 7 | 0.09 |  |  | ✓ |
| A16A0 | OTHER ALIMENTARY TRACT AND METABOLISM PRODUCTS | 2608 | 0.77 | 2096 | 0.66 | 512 | 2.22 | 1026 | 0.87 | 853 | 0.77 | 173 | 2.3 |  |  | ✓ |
| B01A0 | VITAMIN K ANTAGONISTS | 12586 | 3.71 | 11314 | 3.58 | 1272 | 5.5 | 3178 | 2.69 | 2844 | 2.57 | 334 | 4.45 |  | ✓ | ✓ |
| B01B1 | Unfractionated heparins | 67850 | 19.98 | 64541 | 20.4 | 3309 | 14.32 | 23361 | 19.79 | 22287 | 20.16 | 1074 | 14.31 | ✓ | ✓ | ✓ |
| B01B2 | Fractionated heparins | 7570 | 2.23 | 7242 | 2.29 | 328 | 1.42 | 2412 | 2.04 | 2315 | 2.09 | 97 | 1.29 |  | ✓ | ✓ |
| B01B3 | Heparins for flushing | 90771 | 26.74 | 83141 | 26.28 | 7630 | 33.02 | 29928 | 25.35 | 27628 | 24.99 | 2300 | 30.64 | ✓ | ✓ | ✓ |
| B01C1 | Cyclo-oxygenase inhibitor platelet aggregation inhibitors | 30387 | 8.95 | 28122 | 8.89 | 2265 | 9.8 | 10708 | 9.07 | 9933 | 8.98 | 775 | 10.32 | ✓ | ✓ | ✓ |
| B01C2 | ADP (adenosine diphosphate) receptor antagonist platelet aggregation inhibitors | 20555 | 6.05 | 19293 | 6.1 | 1262 | 5.46 | 7389 | 6.26 | 6952 | 6.29 | 437 | 5.82 | ✓ | ✓ | ✓ |
| B01C4 | Platelet cAMP enhancing platelet aggregation inhibitors | 10591 | 3.12 | 9735 | 3.08 | 856 | 3.7 | 3271 | 2.77 | 3005 | 2.72 | 266 | 3.54 |  | ✓ | ✓ |
| B01C5 | Platelet aggregation inhibitors, combinations | 1403 | 0.41 | 1340 | 0.42 | 63 | 0.27 | 151 | 0.13 | 146 | 0.13 | 5 | 0.07 |  |  | ✓ |
| B01C9 | Other platelet aggregation inhibitors | 5734 | 1.69 | 5376 | 1.7 | 358 | 1.55 | 1371 | 1.16 | 1286 | 1.16 | 85 | 1.13 |  | ✓ | ✓ |
| B01D0 | FIBRINOLYTICS | 777 | 0.23 | 738 | 0.23 | 39 | 0.17 | 269 | 0.23 | 253 | 0.23 | 16 | 0.21 |  |  | ✓ |
| B01E0 | DIRECT THROMBIN INHIBITORS | 2813 | 0.83 | 2667 | 0.84 | 146 | 0.63 | 1108 | 0.94 | 1048 | 0.95 | 60 | 0.8 |  |  | ✓ |
| B01F0 | DIRECT FACTOR XA INHIBITORS | 14737 | 4.34 | 13617 | 4.3 | 1120 | 4.85 | 6633 | 5.62 | 6116 | 5.53 | 517 | 6.89 |  | ✓ | ✓ |
| B01X0 | OTHER ANTITHROMBOTIC AGENTS | 1063 | 0.31 | 975 | 0.31 | 88 | 0.38 | 357 | 0.3 | 323 | 0.29 | 34 | 0.45 |  |  | ✓ |
| B02A1 | Synthetic antifibrinolytics | 27005 | 7.95 | 25509 | 8.06 | 1496 | 6.47 | 9388 | 7.95 | 8861 | 8.01 | 527 | 7.02 | ✓ | ✓ | ✓ |
| B02B1 | Vitamin K | 4874 | 1.44 | 4256 | 1.35 | 618 | 2.67 | 1333 | 1.13 | 1174 | 1.06 | 159 | 2.12 |  | ✓ | ✓ |
| B02C1 | Coagulation inhibitors | 481 | 0.14 | 432 | 0.14 | 49 | 0.21 | 147 | 0.12 | 133 | 0.12 | 14 | 0.19 |  |  | ✓ |
| B02C2 | Inhibitors of the Kallikrein-kinin-system | 15352 | 4.52 | 13800 | 4.36 | 1552 | 6.72 | 4535 | 3.84 | 4110 | 3.72 | 425 | 5.66 |  | ✓ | ✓ |
| B02D1 | FACTOR VIII, INCLUDING SUBSTITUTES | 49 | 0.01 | 40 | 0.01 | 9 | 0.04 | 12 | 0.01 | 10 | 0.01 | 2 | 0.03 |  |  | ✓ |
| B02D4 | Factor XIII | 139 | 0.04 | 123 | 0.04 | 16 | 0.07 | 55 | 0.05 | 52 | 0.05 | 3 | 0.04 |  |  | ✓ |
| B02D6 | Fresh frozen plasma and antihaemophilic plasma | 3671 | 1.08 | 3376 | 1.07 | 295 | 1.28 | 1011 | 0.86 | 934 | 0.84 | 77 | 1.03 |  |  | ✓ |
| B02D8 | Platelet concentrates | 4441 | 1.31 | 3912 | 1.24 | 529 | 2.29 | 1239 | 1.05 | 1083 | 0.98 | 156 | 2.08 |  | ✓ | ✓ |
| B02E0 | THROMBOPOIETIN AGONISTS | 153 | 0.05 | 122 | 0.04 | 31 | 0.13 | 66 | 0.06 | 57 | 0.05 | 9 | 0.12 |  |  | ✓ |
| B02F0 | TISSUE SEALING PREPARATIONS | 9431 | 2.78 | 9037 | 2.86 | 394 | 1.71 | 3257 | 2.76 | 3123 | 2.82 | 134 | 1.79 |  | ✓ | ✓ |
| B02G0 | SYSTEMIC HAEMOSTATICS | 27579 | 8.12 | 25776 | 8.15 | 1803 | 7.8 | 9181 | 7.78 | 8604 | 7.78 | 577 | 7.69 | ✓ | ✓ | ✓ |
| B03A1 | Plain iron | 21873 | 6.44 | 19707 | 6.23 | 2166 | 9.37 | 7599 | 6.44 | 6895 | 6.24 | 704 | 9.38 | ✓ | ✓ | ✓ |
| B03C0 | ERYTHROPOIETIN PRODUCTS | 7009 | 2.06 | 6209 | 1.96 | 800 | 3.46 | 2470 | 2.09 | 2190 | 1.98 | 280 | 3.73 |  | ✓ | ✓ |
| B03X0 | OTHER ANTI-ANAEMIC PRODUCTS, INCLUDING FOLIC ACID, FOLINIC ACID | 1671 | 0.49 | 1476 | 0.47 | 195 | 0.84 | 656 | 0.56 | 560 | 0.51 | 96 | 1.28 |  |  | ✓ |
| C01A1 | Plain cardiac glycosides | 4101 | 1.21 | 3654 | 1.15 | 447 | 1.93 | 1077 | 0.91 | 975 | 0.88 | 102 | 1.36 |  |  | ✓ |
| C01B0 | ANTI-ARRHYTHMICS | 12802 | 3.77 | 11981 | 3.79 | 821 | 3.55 | 3657 | 3.1 | 3419 | 3.09 | 238 | 3.17 |  | ✓ | ✓ |
| C01C1 | Cardiac stimulants excluding dopaminergic agents | 41002 | 12.08 | 39143 | 12.37 | 1859 | 8.04 | 15592 | 13.21 | 14925 | 13.5 | 667 | 8.89 | ✓ | ✓ | ✓ |
| C01C2 | Cardiac dopaminergic agents | 10831 | 3.19 | 10063 | 3.18 | 768 | 3.32 | 3512 | 2.97 | 3280 | 2.97 | 232 | 3.09 |  | ✓ | ✓ |
| C01D0 | CORONARY THERAPY EXCLUDING CALCIUM ANTAGONISTS AND NITRITES | 14846 | 4.37 | 13938 | 4.41 | 908 | 3.93 | 4295 | 3.64 | 4050 | 3.66 | 245 | 3.26 |  | ✓ | ✓ |
| C01E0 | NITRITES AND NITRATES | 31831 | 9.38 | 29888 | 9.45 | 1943 | 8.41 | 10865 | 9.2 | 10292 | 9.31 | 573 | 7.63 | ✓ | ✓ | ✓ |
| C01F0 | POSITIVE INOTROPIC AGENTS | 1991 | 0.59 | 1779 | 0.56 | 212 | 0.92 | 595 | 0.5 | 529 | 0.48 | 66 | 0.88 |  |  | ✓ |
| C01X0 | ALL OTHER CARDIAC PREPARATIONS | 10092 | 2.97 | 9209 | 2.91 | 883 | 3.82 | 2909 | 2.46 | 2667 | 2.41 | 242 | 3.22 |  | ✓ | ✓ |
| C02A1 | Antihypertensives plain, mainly centrally acting | 529 | 0.16 | 493 | 0.16 | 36 | 0.16 | 137 | 0.12 | 122 | 0.11 | 15 | 0.2 |  |  | ✓ |
| C02A2 | Antihypertensives plain, mainly peripherally acting | 6846 | 2.02 | 6275 | 1.98 | 571 | 2.47 | 2417 | 2.05 | 2230 | 2.02 | 187 | 2.49 |  | ✓ | ✓ |
| C02A3 | Antihypertensives plain, others | 1431 | 0.42 | 1273 | 0.4 | 158 | 0.68 | 541 | 0.46 | 498 | 0.45 | 43 | 0.57 |  |  | ✓ |
| C02C0 | RAUWOLFIA ALKALOIDS AND OTHER ANTIHYPERTENSIVES OF HERBAL ORIGIN | 32 | 0.01 | 26 | 0.01 | 6 | 0.03 | 14 | 0.01 | 13 | 0.01 | 1 | 0.01 |  |  | ✓ |
| C03A1 | Potassium-sparing agents plain | 17138 | 5.05 | 14739 | 4.66 | 2399 | 10.38 | 5659 | 4.79 | 4945 | 4.47 | 714 | 9.51 |  | ✓ | ✓ |
| C03A2 | Loop diuretics plain | 51957 | 15.3 | 45903 | 14.51 | 6054 | 26.2 | 17269 | 14.63 | 15376 | 13.91 | 1893 | 25.22 | ✓ | ✓ | ✓ |
| C03A3 | Thiazides and analogues plain | 4770 | 1.4 | 4279 | 1.35 | 491 | 2.12 | 1551 | 1.31 | 1404 | 1.27 | 147 | 1.96 |  | ✓ | ✓ |
| C03A7 | Vasopressin receptor antagonist diuretics | 5785 | 1.7 | 4757 | 1.5 | 1028 | 4.45 | 2700 | 2.29 | 2261 | 2.04 | 439 | 5.85 |  | ✓ | ✓ |
| C03A9 | Other diuretics | 287 | 0.08 | 262 | 0.08 | 25 | 0.11 | 96 | 0.08 | 83 | 0.08 | 13 | 0.17 |  |  | ✓ |
| C04A1 | Cerebral and peripheral vasotherapeutics excluding calcium antagonists with cerebral activity | 3023 | 0.89 | 2785 | 0.88 | 238 | 1.03 | 768 | 0.65 | 716 | 0.65 | 52 | 0.69 |  |  | ✓ |
| C04A2 | Calcium antagonists with cerebral activity | 248 | 0.07 | 246 | 0.08 | 2 | 0.01 | 80 | 0.07 | 78 | 0.07 | 2 | 0.03 |  |  | ✓ |
| C05A1 | Topical anti-haemorrhoidals with corticosteroids | 3921 | 1.15 | 3532 | 1.12 | 389 | 1.68 | 1240 | 1.05 | 1121 | 1.01 | 119 | 1.59 |  | ✓ | ✓ |
| C05A2 | Topical anti-haemorrhoidals without corticosteroids | 797 | 0.23 | 714 | 0.23 | 83 | 0.36 | 258 | 0.22 | 231 | 0.21 | 27 | 0.36 |  |  | ✓ |
| C05B0 | VARICOSE THERAPY, TOPICAL | 14272 | 4.2 | 12962 | 4.1 | 1310 | 5.67 | 5044 | 4.27 | 4622 | 4.18 | 422 | 5.62 |  | ✓ | ✓ |
| C06B1 | ENDOTHELIN RECEPTOR ANTAGONIST PAH PRODUCTS | 105 | 0.03 | 94 | 0.03 | 11 | 0.05 | 25 | 0.02 | 22 | 0.02 | 3 | 0.04 |  |  | ✓ |
| C06B2 | PDE5 INHIBITOR PAH PRODUCTS | 100 | 0.03 | 93 | 0.03 | 7 | 0.03 | 20 | 0.02 | 19 | 0.02 | 1 | 0.01 |  |  | ✓ |
| C06B3 | PROSTACYCLIN AGONIST PAH PRODUCTS | 22 | 0.01 | 21 | 0.01 | 1 | 0 | 21 | 0.02 | 18 | 0.02 | 3 | 0.04 |  |  | ✓ |
| C06X0 | OTHER CARDIOVASCULAR PRODUCTS | 4990 | 1.47 | 4769 | 1.51 | 221 | 0.96 | 1557 | 1.32 | 1492 | 1.35 | 65 | 0.87 |  | ✓ | ✓ |
| C07A0 | BETA-BLOCKING AGENTS, PLAIN | 33202 | 9.78 | 30436 | 9.62 | 2766 | 11.97 | 12468 | 10.56 | 11465 | 10.37 | 1003 | 13.36 | ✓ | ✓ | ✓ |
| C08A0 | CALCIUM ANTAGONISTS, PLAIN | 69993 | 20.62 | 64760 | 20.47 | 5233 | 22.65 | 24636 | 20.86 | 22852 | 20.67 | 1784 | 23.76 | ✓ | ✓ | ✓ |
| C09A0 | ACE INHIBITORS, PLAIN | 12964 | 3.82 | 11949 | 3.78 | 1015 | 4.39 | 4282 | 3.63 | 3959 | 3.58 | 323 | 4.3 |  | ✓ | ✓ |
| C09C0 | ANGIOTENSIN-II ANTAGONISTS, PLAIN | 38219 | 11.26 | 35365 | 11.18 | 2854 | 12.35 | 12973 | 10.99 | 12093 | 10.94 | 880 | 11.72 | ✓ | ✓ | ✓ |
| C09D1 | Angiotensin-II antagonist combinations with antihypertensives (C2) and/or diuretics | 1436 | 0.42 | 1328 | 0.42 | 108 | 0.47 | 303 | 0.26 | 288 | 0.26 | 15 | 0.2 |  |  | ✓ |
| C09D3 | Angiotensin-II antagonist combinations with calcium antagonists | 2913 | 0.86 | 2718 | 0.86 | 195 | 0.84 | 610 | 0.52 | 573 | 0.52 | 37 | 0.49 |  |  | ✓ |
| C09X0 | OTHER AGENTS ACTING ON THE RENIN-ANGIOTENSIN SYSTEM | 118 | 0.03 | 111 | 0.04 | 7 | 0.03 | 14 | 0.01 | 12 | 0.01 | 2 | 0.03 |  |  | ✓ |
| C10A1 | Statins (HMG-CoA reductase inhibitors) | 36551 | 10.77 | 34236 | 10.82 | 2315 | 10.02 | 13310 | 11.27 | 12495 | 11.3 | 815 | 10.86 | ✓ | ✓ | ✓ |
| C10A2 | Fibrates | 2126 | 0.63 | 1991 | 0.63 | 135 | 0.58 | 606 | 0.51 | 572 | 0.52 | 34 | 0.45 |  |  | ✓ |
| C10A4 | PCSK9 inhibitors | 20 | 0.01 | 20 | 0.01 | 0 | 0 | 22 | 0.02 | 22 | 0.02 | 0 | 0 |  |  | ✓ |
| C10A9 | All other cholesterol/triglyceride regulators | 3985 | 1.17 | 3694 | 1.17 | 291 | 1.26 | 1611 | 1.36 | 1511 | 1.37 | 100 | 1.33 |  | ✓ | ✓ |
| C10B0 | ANTI-ATHEROMA PREPARATIONS OF NATURAL ORIGIN | 2994 | 0.88 | 2801 | 0.89 | 193 | 0.84 | 969 | 0.82 | 916 | 0.83 | 53 | 0.71 |  |  | ✓ |
| C11A1 | Lipid-regulating cardiovascular multitherapy fixed combination products | 227 | 0.07 | 216 | 0.07 | 11 | 0.05 | 91 | 0.08 | 89 | 0.08 | 2 | 0.03 |  |  | ✓ |
| D01A1 | Topical dermatological antifungals | 6533 | 1.92 | 5897 | 1.86 | 636 | 2.75 | 1997 | 1.69 | 1842 | 1.67 | 155 | 2.06 |  | ✓ | ✓ |
| D02A0 | EMOLLIENTS, PROTECTIVES | 6215 | 1.83 | 5645 | 1.78 | 570 | 2.47 | 2002 | 1.7 | 1821 | 1.65 | 181 | 2.41 |  |  | ✓ |
| D03A9 | All other wound healing agents | 23331 | 6.87 | 21618 | 6.83 | 1713 | 7.41 | 8336 | 7.06 | 7745 | 7 | 591 | 7.87 | ✓ | ✓ | ✓ |
| D04A0 | ANTI-PRURITICS, INCLUDING TOPICAL ANTIHISTAMINES, ANAESTHETICS, ETC | 10032 | 2.95 | 9220 | 2.91 | 812 | 3.51 | 3395 | 2.88 | 3150 | 2.85 | 245 | 3.26 |  | ✓ | ✓ |
| D05A0 | TOPICAL ANTIPSORIASIS PRODUCTS | 589 | 0.17 | 540 | 0.17 | 49 | 0.21 | 142 | 0.12 | 127 | 0.11 | 15 | 0.2 |  |  | ✓ |
| D05B0 | SYSTEMIC ANTIPSORIASIS PRODUCTS | 88 | 0.03 | 81 | 0.03 | 7 | 0.03 | 13 | 0.01 | 10 | 0.01 | 3 | 0.04 |  |  | ✓ |
| D05X0 | OTHER NONSTEROIDAL PRODUCTS FOR INFLAMMATORY SKIN DISORDERS | 278 | 0.08 | 249 | 0.08 | 29 | 0.13 | 94 | 0.08 | 85 | 0.08 | 9 | 0.12 |  |  | ✓ |
| D06A0 | TOPICAL ANTIBACTERIALS | 12649 | 3.73 | 11668 | 3.69 | 981 | 4.25 | 3852 | 3.26 | 3605 | 3.26 | 247 | 3.29 |  | ✓ | ✓ |
| D06D1 | Topical antivirals | 1343 | 0.4 | 1227 | 0.39 | 116 | 0.5 | 413 | 0.35 | 383 | 0.35 | 30 | 0.4 |  |  | ✓ |
| D07A0 | PLAIN TOPICAL CORTICOSTEROIDS | 13994 | 4.12 | 12920 | 4.08 | 1074 | 4.65 | 4544 | 3.85 | 4188 | 3.79 | 356 | 4.74 |  | ✓ | ✓ |
| D07B1 | Combinations of corticosteroids with antibacterials | 8469 | 2.49 | 7886 | 2.49 | 583 | 2.52 | 2451 | 2.08 | 2311 | 2.09 | 140 | 1.86 |  | ✓ | ✓ |
| D07B4 | Other corticosteroid combinations | 2173 | 0.64 | 1981 | 0.63 | 192 | 0.83 | 414 | 0.35 | 380 | 0.34 | 34 | 0.45 |  |  | ✓ |
| D08A0 | ANTISEPTICS AND DISINFECTANTS | 50853 | 14.98 | 47279 | 14.94 | 3574 | 15.47 | 17384 | 14.72 | 16169 | 14.62 | 1215 | 16.18 | ✓ | ✓ | ✓ |
| D10A0 | TOPICAL ANTI-ACNE PREPARATIONS | 536 | 0.16 | 496 | 0.16 | 40 | 0.17 | 135 | 0.11 | 130 | 0.12 | 5 | 0.07 |  |  | ✓ |
| D11A0 | OTHER DERMATOLOGICAL PREPARATIONS | 72 | 0.02 | 69 | 0.02 | 3 | 0.01 | 24 | 0.02 | 22 | 0.02 | 2 | 0.03 |  |  | ✓ |
| G01A1 | Systemic trichomonacides | 2265 | 0.67 | 2040 | 0.64 | 225 | 0.97 | 966 | 0.82 | 875 | 0.79 | 91 | 1.21 |  |  | ✓ |
| G01A2 | Topical trichomonacides | 55 | 0.02 | 52 | 0.02 | 3 | 0.01 | 26 | 0.02 | 22 | 0.02 | 4 | 0.05 |  |  | ✓ |
| G01C0 | GYNAECOLOGICAL ANTIBACTERIALS | 589 | 0.17 | 568 | 0.18 | 21 | 0.09 | 273 | 0.23 | 270 | 0.24 | 3 | 0.04 |  |  | ✓ |
| G02A0 | UTEROTONIC PRODUCTS | 479 | 0.14 | 456 | 0.14 | 23 | 0.1 | 117 | 0.1 | 109 | 0.1 | 8 | 0.11 |  |  | ✓ |
| G02E0 | LABOUR INHIBITORS | 83 | 0.02 | 79 | 0.02 | 4 | 0.02 | 15 | 0.01 | 15 | 0.01 | 0 | 0 |  |  | ✓ |
| G03C0 | OESTROGENS, EXCLUDING G3A, G3E, G3F | 455 | 0.13 | 422 | 0.13 | 33 | 0.14 | 126 | 0.11 | 119 | 0.11 | 7 | 0.09 |  |  | ✓ |
| G03D0 | PROGESTOGENS, EXCLUDING G3A, G3F | 83 | 0.02 | 77 | 0.02 | 6 | 0.03 | 27 | 0.02 | 26 | 0.02 | 1 | 0.01 |  |  | ✓ |
| G03F0 | OESTROGEN WITH PROGESTOGEN COMBINATIONS, EXCLUDING G3A | 71 | 0.02 | 63 | 0.02 | 8 | 0.03 | 14 | 0.01 | 13 | 0.01 | 1 | 0.01 |  |  | ✓ |
| G03J0 | SERMS (SELECTIVE OESTROGEN RECEPTOR MODULATORS) | 1063 | 0.31 | 980 | 0.31 | 83 | 0.36 | 312 | 0.26 | 297 | 0.27 | 15 | 0.2 |  |  | ✓ |
| G04C2 | BPH alpha-adrenergic antagonists, plain | 11471 | 3.38 | 10234 | 3.23 | 1237 | 5.35 | 3999 | 3.39 | 3574 | 3.23 | 425 | 5.66 |  | ✓ | ✓ |
| G04C3 | BPH 5-alpha testosterone reductase inhibitors (5-ARI), plain | 1299 | 0.38 | 1178 | 0.37 | 121 | 0.52 | 543 | 0.46 | 491 | 0.44 | 52 | 0.69 |  |  | ✓ |
| G04C9 | BPH products, other | 1055 | 0.31 | 943 | 0.3 | 112 | 0.48 | 301 | 0.25 | 274 | 0.25 | 27 | 0.36 |  |  | ✓ |
| G04D4 | Urinary incontinence products | 5115 | 1.51 | 4627 | 1.46 | 488 | 2.11 | 1970 | 1.67 | 1787 | 1.62 | 183 | 2.44 |  | ✓ | ✓ |
| G04X0 | ALL OTHER UROLOGICAL PRODUCTS | 474 | 0.14 | 428 | 0.14 | 46 | 0.2 | 168 | 0.14 | 156 | 0.14 | 12 | 0.16 |  |  | ✓ |
| H01A0 | ACTH | 137 | 0.04 | 126 | 0.04 | 11 | 0.05 | 62 | 0.05 | 55 | 0.05 | 7 | 0.09 |  |  | ✓ |
| H01C1 | Gonadotrophin-releasing hormones | 36 | 0.01 | 33 | 0.01 | 3 | 0.01 | 12 | 0.01 | 11 | 0.01 | 1 | 0.01 |  |  | ✓ |
| H01C2 | Antigrowth hormones | 505 | 0.15 | 422 | 0.13 | 83 | 0.36 | 223 | 0.19 | 196 | 0.18 | 27 | 0.36 |  |  | ✓ |
| H02A1 | Injectable corticosteroids, plain | 69583 | 20.49 | 64648 | 20.43 | 4935 | 21.36 | 22977 | 19.46 | 21398 | 19.35 | 1579 | 21.03 | ✓ | ✓ | ✓ |
| H02A2 | Oral corticosteroids, plain | 30723 | 9.05 | 27555 | 8.71 | 3168 | 13.71 | 10225 | 8.66 | 9176 | 8.3 | 1049 | 13.97 | ✓ | ✓ | ✓ |
| H02B0 | SYSTEMIC CORTICOSTEROID COMBINATIONS | 862 | 0.25 | 789 | 0.25 | 73 | 0.32 | 156 | 0.13 | 139 | 0.13 | 17 | 0.23 |  |  | ✓ |
| H03A0 | THYROID PREPARATIONS | 5996 | 1.77 | 5337 | 1.69 | 659 | 2.85 | 2233 | 1.89 | 1998 | 1.81 | 235 | 3.13 |  | ✓ | ✓ |
| H03B0 | ANTI-THYROID PREPARATIONS | 691 | 0.2 | 630 | 0.2 | 61 | 0.26 | 203 | 0.17 | 192 | 0.17 | 11 | 0.15 |  |  | ✓ |
| H03C0 | IODINE THERAPY | 179 | 0.05 | 167 | 0.05 | 12 | 0.05 | 41 | 0.03 | 40 | 0.04 | 1 | 0.01 |  |  | ✓ |
| H04A0 | CALCITONINS | 855 | 0.25 | 776 | 0.25 | 79 | 0.34 | 212 | 0.18 | 186 | 0.17 | 26 | 0.35 |  |  | ✓ |
| H04B0 | GLUCAGON | 12117 | 3.57 | 11094 | 3.51 | 1023 | 4.43 | 4140 | 3.51 | 3812 | 3.45 | 328 | 4.37 |  | ✓ | ✓ |
| H04D0 | ANTIDIURETIC HORMONES | 607 | 0.18 | 574 | 0.18 | 33 | 0.14 | 179 | 0.15 | 175 | 0.16 | 4 | 0.05 |  |  | ✓ |
| H04E0 | PARATHYROID HORMONES AND ANALOGUES | 1075 | 0.32 | 1015 | 0.32 | 60 | 0.26 | 395 | 0.33 | 372 | 0.34 | 23 | 0.31 |  |  | ✓ |
| H04F0 | ANTIPARATHYROID PRODUCTS | 2249 | 0.66 | 2040 | 0.64 | 209 | 0.9 | 916 | 0.78 | 825 | 0.75 | 91 | 1.21 |  |  | ✓ |
| J01A0 | TETRACYCLINES AND COMBINATIONS | 4612 | 1.36 | 4253 | 1.34 | 359 | 1.55 | 1326 | 1.12 | 1226 | 1.11 | 100 | 1.33 |  | ✓ | ✓ |
| J01C1 | Oral broad spectrum penicillins | 12817 | 3.78 | 11802 | 3.73 | 1015 | 4.39 | 5240 | 4.44 | 4916 | 4.45 | 324 | 4.32 |  | ✓ | ✓ |
| J01C2 | Injectable broad spectrum penicillins | 41950 | 12.36 | 37802 | 11.95 | 4148 | 17.95 | 15166 | 12.84 | 13754 | 12.44 | 1412 | 18.81 | ✓ | ✓ | ✓ |
| J01D1 | Oral cephalosporins | 29852 | 8.79 | 28392 | 8.97 | 1460 | 6.32 | 7740 | 6.56 | 7369 | 6.66 | 371 | 4.94 | ✓ | ✓ | ✓ |
| J01D2 | Injectable cephalosporins | 148288 | 43.68 | 139299 | 44.03 | 8989 | 38.9 | 51812 | 43.88 | 48910 | 44.24 | 2902 | 38.66 | ✓ | ✓ | ✓ |
| J01E0 | TRIMETHOPRIM AND SIMILAR FORMULATIONS | 10608 | 3.12 | 9649 | 3.05 | 959 | 4.15 | 3532 | 2.99 | 3219 | 2.91 | 313 | 4.17 |  | ✓ | ✓ |
| J01F0 | MACROLIDES AND SIMILAR TYPES | 12986 | 3.82 | 12018 | 3.8 | 968 | 4.19 | 3853 | 3.26 | 3559 | 3.22 | 294 | 3.92 |  | ✓ | ✓ |
| J01G1 | Oral fluoroquinolones | 23603 | 6.95 | 21697 | 6.86 | 1906 | 8.25 | 7585 | 6.42 | 6988 | 6.32 | 597 | 7.95 | ✓ | ✓ | ✓ |
| J01G2 | Injectable fluoroquinolones | 3730 | 1.1 | 3325 | 1.05 | 405 | 1.75 | 1286 | 1.09 | 1147 | 1.04 | 139 | 1.85 |  |  | ✓ |
| J01H1 | Plain medium and narrow spectrum penicillins | 199 | 0.06 | 190 | 0.06 | 9 | 0.04 | 54 | 0.05 | 49 | 0.04 | 5 | 0.07 |  |  | ✓ |
| J01K0 | AMINOGLYCOSIDES | 4151 | 1.22 | 3816 | 1.21 | 335 | 1.45 | 1065 | 0.9 | 989 | 0.89 | 76 | 1.01 |  |  | ✓ |
| J01M0 | RIFAMPICIN/RIFAMYCIN | 580 | 0.17 | 531 | 0.17 | 49 | 0.21 | 184 | 0.16 | 165 | 0.15 | 19 | 0.25 |  |  | ✓ |
| J01P2 | Penems and carbapenems | 10870 | 3.2 | 9516 | 3.01 | 1354 | 5.86 | 3352 | 2.84 | 2917 | 2.64 | 435 | 5.79 |  | ✓ | ✓ |
| J01X1 | Glycopeptide antibacterials | 3776 | 1.11 | 3367 | 1.06 | 409 | 1.77 | 1378 | 1.17 | 1226 | 1.11 | 152 | 2.02 |  | ✓ | ✓ |
| J01X2 | Polymyxins | 406 | 0.12 | 387 | 0.12 | 19 | 0.08 | 126 | 0.11 | 119 | 0.11 | 7 | 0.09 |  |  | ✓ |
| J01X9 | All other antibacterials | 2304 | 0.68 | 2146 | 0.68 | 158 | 0.68 | 703 | 0.6 | 660 | 0.6 | 43 | 0.57 |  |  | ✓ |
| J02A0 | SYSTEMIC AGENTS FOR FUNGAL INFECTIONS | 5258 | 1.55 | 4655 | 1.47 | 603 | 2.61 | 1517 | 1.28 | 1362 | 1.23 | 155 | 2.06 |  | ✓ | ✓ |
| J04A1 | Antituberculars, single ingredient | 277 | 0.08 | 238 | 0.08 | 39 | 0.17 | 97 | 0.08 | 78 | 0.07 | 19 | 0.25 |  |  | ✓ |
| J04A9 | Antituberculars, others | 262 | 0.08 | 231 | 0.07 | 31 | 0.13 | 80 | 0.07 | 69 | 0.06 | 11 | 0.15 |  |  | ✓ |
| J05B3 | Herpes antivirals | 4500 | 1.33 | 4154 | 1.31 | 346 | 1.5 | 1515 | 1.28 | 1401 | 1.27 | 114 | 1.52 |  | ✓ | ✓ |
| J05B4 | Influenza antivirals | 3350 | 0.99 | 3113 | 0.98 | 237 | 1.03 | 1315 | 1.11 | 1238 | 1.12 | 77 | 1.03 |  |  | ✓ |
| J05C1 | Nucleoside and nucleotide reverse transcriptase inhibitors | 37 | 0.01 | 31 | 0.01 | 6 | 0.03 | 15 | 0.01 | 14 | 0.01 | 1 | 0.01 |  |  | ✓ |
| J05D2 | HEPATITIS B ANTIVIRALS | 850 | 0.25 | 755 | 0.24 | 95 | 0.41 | 318 | 0.27 | 290 | 0.26 | 28 | 0.37 |  |  | ✓ |
| J06A1 | Snake-bite sera | 70 | 0.02 | 68 | 0.02 | 2 | 0.01 | 14 | 0.01 | 13 | 0.01 | 1 | 0.01 |  |  | ✓ |
| J06C0 | POLYVALENT IMMUNO-GLOBULINS - INTRAVENOUS | 2147 | 0.63 | 2001 | 0.63 | 146 | 0.63 | 680 | 0.58 | 632 | 0.57 | 48 | 0.64 |  |  | ✓ |
| J06G1 | Tetanus immunoglobulin | 458 | 0.13 | 449 | 0.14 | 9 | 0.04 | 43 | 0.04 | 41 | 0.04 | 2 | 0.03 |  |  | ✓ |
| J07D1 | Pneumococcal vaccines | 44 | 0.01 | 43 | 0.01 | 1 | 0 | 13 | 0.01 | 13 | 0.01 | 0 | 0 |  |  | ✓ |
| J07D5 | Tetanus vaccines | 1118 | 0.33 | 1090 | 0.34 | 28 | 0.12 | 218 | 0.18 | 213 | 0.19 | 5 | 0.07 |  |  | ✓ |
| J08B0 | ANAEROBICIDES | 389 | 0.11 | 352 | 0.11 | 37 | 0.16 | 174 | 0.15 | 155 | 0.14 | 19 | 0.25 |  |  | ✓ |
| K01A1 | 1/1 - Electrolyte solutions | 136 | 0.04 | 130 | 0.04 | 6 | 0.03 | 82 | 0.07 | 78 | 0.07 | 4 | 0.05 |  |  | ✓ |
| K01A3 | 1/2 - Electrolyte solutions | 69179 | 20.38 | 63939 | 20.21 | 5240 | 22.68 | 22643 | 19.18 | 21013 | 19 | 1630 | 21.71 | ✓ | ✓ | ✓ |
| K01A4 | 1/3 - Electrolyte solutions | 180727 | 53.23 | 169085 | 53.44 | 11642 | 50.38 | 58103 | 49.21 | 54578 | 49.36 | 3525 | 46.96 | ✓ | ✓ | ✓ |
| K01A7 | Ringer's and Ringer's lactate solutions | 77251 | 22.75 | 73150 | 23.12 | 4101 | 17.75 | 20897 | 17.7 | 19834 | 17.94 | 1063 | 14.16 | ✓ | ✓ | ✓ |
| K01A9 | Other electrolyte solutions | 172489 | 50.8 | 161232 | 50.96 | 11257 | 48.71 | 64880 | 54.95 | 61098 | 55.26 | 3782 | 50.38 | ✓ | ✓ | ✓ |
| K01B1 | Sodium chloride solutions | 287905 | 84.8 | 268427 | 84.84 | 19478 | 84.29 | 101750 | 86.17 | 95233 | 86.13 | 6517 | 86.81 | ✓ | ✓ | ✓ |
| K01B3 | Carbohydrate solutions (<=10%) | 50256 | 14.8 | 46076 | 14.56 | 4180 | 18.09 | 15558 | 13.18 | 14315 | 12.95 | 1243 | 16.56 | ✓ | ✓ | ✓ |
| K01C1 | Solutions with one carbohydrate (>10%) | 13542 | 3.99 | 12121 | 3.83 | 1421 | 6.15 | 3951 | 3.35 | 3595 | 3.25 | 356 | 4.74 |  | ✓ | ✓ |
| K01C3 | Carbohydrate electrolyte combination solutions (>10%) | 1230 | 0.36 | 1169 | 0.37 | 61 | 0.26 | 323 | 0.27 | 295 | 0.27 | 28 | 0.37 |  |  | ✓ |
| K01D1 | Fat emulsions, plain | 4204 | 1.24 | 3756 | 1.19 | 448 | 1.94 | 1346 | 1.14 | 1217 | 1.1 | 129 | 1.72 |  | ✓ | ✓ |
| K01E1 | Amino acid standard solutions | 53676 | 15.81 | 48538 | 15.34 | 5138 | 22.23 | 17712 | 15 | 16171 | 14.63 | 1541 | 20.53 | ✓ | ✓ | ✓ |
| K01E2 | Multi-litre concept solutions | 199 | 0.06 | 177 | 0.06 | 22 | 0.1 | 52 | 0.04 | 50 | 0.05 | 2 | 0.03 |  |  | ✓ |
| K01E3 | Nephro solutions | 849 | 0.25 | 743 | 0.23 | 106 | 0.46 | 279 | 0.24 | 242 | 0.22 | 37 | 0.49 |  |  | ✓ |
| K01E4 | Hepatic solutions | 1461 | 0.43 | 1111 | 0.35 | 350 | 1.51 | 363 | 0.31 | 273 | 0.25 | 90 | 1.2 |  |  | ✓ |
| K01F1 | Osmotic therapy | 10145 | 2.99 | 9628 | 3.04 | 517 | 2.24 | 3417 | 2.89 | 3222 | 2.91 | 195 | 2.6 |  | ✓ | ✓ |
| K02A1 | Low dextrans | 1806 | 0.53 | 1739 | 0.55 | 67 | 0.29 | 465 | 0.39 | 449 | 0.41 | 16 | 0.21 |  |  | ✓ |
| K02B0 | STARCHES | 25660 | 7.56 | 24508 | 7.75 | 1152 | 4.99 | 8840 | 7.49 | 8507 | 7.69 | 333 | 4.44 | ✓ | ✓ | ✓ |
| K03A0 | WHOLE BLOOD AND PLASMA FRACTIONS | 21792 | 6.42 | 19100 | 6.04 | 2692 | 11.65 | 6917 | 5.86 | 6131 | 5.55 | 786 | 10.47 | ✓ | ✓ | ✓ |
| K03B1 | Protein solutions <5,0% | 475 | 0.14 | 425 | 0.13 | 50 | 0.22 | 131 | 0.11 | 118 | 0.11 | 13 | 0.17 |  |  | ✓ |
| K03B2 | Protein solutions 5,0% | 5005 | 1.47 | 4637 | 1.47 | 368 | 1.59 | 1238 | 1.05 | 1152 | 1.04 | 86 | 1.15 |  | ✓ | ✓ |
| K03B3 | Protein solutions >5,0% | 4595 | 1.35 | 3908 | 1.24 | 687 | 2.97 | 1232 | 1.04 | 1080 | 0.98 | 152 | 2.02 |  | ✓ | ✓ |
| K04A1 | Electrolyte solutions (<=20ml) | 6517 | 1.92 | 6040 | 1.91 | 477 | 2.06 | 2835 | 2.4 | 2608 | 2.36 | 227 | 3.02 |  | ✓ | ✓ |
| K04B1 | Standard solutions (<=20ml) | 7646 | 2.25 | 6825 | 2.16 | 821 | 3.55 | 2295 | 1.94 | 2026 | 1.83 | 269 | 3.58 |  | ✓ | ✓ |
| K04D0 | OTHER INJECTION SOLUTIONS/INFUSION ADDITIVES (<100ml) | 2131 | 0.63 | 1863 | 0.59 | 268 | 1.16 | 517 | 0.44 | 445 | 0.4 | 72 | 0.96 |  |  | ✓ |
| K05A2 | Saline | 962 | 0.28 | 908 | 0.29 | 54 | 0.23 | 309 | 0.26 | 295 | 0.27 | 14 | 0.19 |  |  | ✓ |
| K05A9 | Other irrigating solutions | 3480 | 1.02 | 3366 | 1.06 | 114 | 0.49 | 1276 | 1.08 | 1242 | 1.12 | 34 | 0.45 |  | ✓ | ✓ |
| K06A0 | HAEMODIALYSIS SOLUTIONS | 4208 | 1.24 | 3878 | 1.23 | 330 | 1.43 | 1476 | 1.25 | 1360 | 1.23 | 116 | 1.55 |  | ✓ | ✓ |
| K06B0 | PERITONEAL DIALYSIS SOLUTIONS | 349 | 0.1 | 301 | 0.1 | 48 | 0.21 | 164 | 0.14 | 142 | 0.13 | 22 | 0.29 |  |  | ✓ |
| K06C0 | HAEMOFILTRATION | 1060 | 0.31 | 976 | 0.31 | 84 | 0.36 | 348 | 0.29 | 316 | 0.29 | 32 | 0.43 |  |  | ✓ |
| L01A0 | ALKYLATING AGENTS | 5757 | 1.7 | 5473 | 1.73 | 284 | 1.23 | 1603 | 1.36 | 1513 | 1.37 | 90 | 1.2 |  | ✓ | ✓ |
| L01B0 | ANTIMETABOLITES | 20835 | 6.14 | 19201 | 6.07 | 1634 | 7.07 | 5955 | 5.04 | 5440 | 4.92 | 515 | 6.86 | ✓ | ✓ | ✓ |
| L01C1 | Vinca alkaloid antineoplastics | 4019 | 1.18 | 3844 | 1.21 | 175 | 0.76 | 1210 | 1.02 | 1158 | 1.05 | 52 | 0.69 |  | ✓ | ✓ |
| L01C2 | Taxane antineoplastics | 6674 | 1.97 | 6109 | 1.93 | 565 | 2.45 | 2095 | 1.77 | 1898 | 1.72 | 197 | 2.62 |  | ✓ | ✓ |
| L01C3 | Camptothecin antineoplastics | 4217 | 1.24 | 3916 | 1.24 | 301 | 1.3 | 1146 | 0.97 | 1041 | 0.94 | 105 | 1.4 |  |  | ✓ |
| L01C4 | Podophyllotoxin antineoplastics | 3010 | 0.89 | 2809 | 0.89 | 201 | 0.87 | 1054 | 0.89 | 1002 | 0.91 | 52 | 0.69 |  |  | ✓ |
| L01D0 | ANTINEOPLASTIC ANTIBIOTICS | 8272 | 2.44 | 7843 | 2.48 | 429 | 1.86 | 2377 | 2.01 | 2250 | 2.03 | 127 | 1.69 |  | ✓ | ✓ |
| L01F0 | PLATINUM ANTINEOPLASTICS | 20499 | 6.04 | 19239 | 6.08 | 1260 | 5.45 | 6411 | 5.43 | 6006 | 5.43 | 405 | 5.39 | ✓ | ✓ | ✓ |
| L01G1 | Monoclonal antibody antineoplastics, CD20 | 3544 | 1.04 | 3387 | 1.07 | 157 | 0.68 | 1055 | 0.89 | 998 | 0.9 | 57 | 0.76 |  |  | ✓ |
| L01G2 | Monoclonal antibody antineoplastics, VEGF/VEGFR | 4849 | 1.43 | 4604 | 1.46 | 245 | 1.06 | 1333 | 1.13 | 1227 | 1.11 | 106 | 1.41 |  | ✓ | ✓ |
| L01G3 | Monoclonal antibody antineoplastics, HER-2 | 774 | 0.23 | 733 | 0.23 | 41 | 0.18 | 314 | 0.27 | 296 | 0.27 | 18 | 0.24 |  |  | ✓ |
| L01G4 | Monoclonal antibody antineoplastics, EGFR | 910 | 0.27 | 834 | 0.26 | 76 | 0.33 | 249 | 0.21 | 228 | 0.21 | 21 | 0.28 |  |  | ✓ |
| L01G5 | Monoclonal antibody antineoplastics, PD-1/PD-L1 | 16 | 0 | 10 | 0 | 6 | 0.03 | 30 | 0.03 | 23 | 0.02 | 7 | 0.09 |  |  | ✓ |
| L01G9 | Monoclonal antibody antineoplastics, other | 95 | 0.03 | 87 | 0.03 | 8 | 0.03 | 67 | 0.06 | 55 | 0.05 | 12 | 0.16 |  |  | ✓ |
| L01H1 | Protein kinase inhibitor antineoplastics, BCR-ABL | 339 | 0.1 | 310 | 0.1 | 29 | 0.13 | 83 | 0.07 | 81 | 0.07 | 2 | 0.03 |  |  | ✓ |
| L01H2 | Protein kinase inhibitor antineoplastics, EGFR | 790 | 0.23 | 718 | 0.23 | 72 | 0.31 | 248 | 0.21 | 222 | 0.2 | 26 | 0.35 |  |  | ✓ |
| L01H3 | Protein kinase inhibitor antineoplastics, ALK | 71 | 0.02 | 64 | 0.02 | 7 | 0.03 | 21 | 0.02 | 19 | 0.02 | 2 | 0.03 |  |  | ✓ |
| L01H9 | Protein kinase inhibitor antineoplastics, other | 665 | 0.2 | 558 | 0.18 | 107 | 0.46 | 276 | 0.23 | 247 | 0.22 | 29 | 0.39 |  |  | ✓ |
| L01J0 | PROTEASOME INHIBITOR ANTINEOPLASTICS | 473 | 0.14 | 429 | 0.14 | 44 | 0.19 | 203 | 0.17 | 186 | 0.17 | 17 | 0.23 |  |  | ✓ |
| L01X2 | Lidomide antineoplastics | 266 | 0.08 | 223 | 0.07 | 43 | 0.19 | 130 | 0.11 | 111 | 0.1 | 19 | 0.25 |  |  | ✓ |
| L01X9 | All other antineoplastics | 977 | 0.29 | 854 | 0.27 | 123 | 0.53 | 339 | 0.29 | 302 | 0.27 | 37 | 0.49 |  |  | ✓ |
| L02A1 | Cytostatic oestrogens | 34 | 0.01 | 22 | 0.01 | 12 | 0.05 | 11 | 0.01 | 8 | 0.01 | 3 | 0.04 |  |  | ✓ |
| L02A2 | Cytostatic progestogens | 263 | 0.08 | 228 | 0.07 | 35 | 0.15 | 73 | 0.06 | 64 | 0.06 | 9 | 0.12 |  |  | ✓ |
| L02A3 | Cytostatic gonadotrophin-releasing hormone analogues | 267 | 0.08 | 233 | 0.07 | 34 | 0.15 | 64 | 0.05 | 57 | 0.05 | 7 | 0.09 |  |  | ✓ |
| L02B1 | Cytostatic anti-oestrogens | 227 | 0.07 | 210 | 0.07 | 17 | 0.07 | 49 | 0.04 | 46 | 0.04 | 3 | 0.04 |  |  | ✓ |
| L02B2 | Cytostatic anti-androgens | 885 | 0.26 | 780 | 0.25 | 105 | 0.45 | 319 | 0.27 | 285 | 0.26 | 34 | 0.45 |  |  | ✓ |
| L02B3 | Cytostatic aromatase inhibitors | 527 | 0.16 | 487 | 0.15 | 40 | 0.17 | 191 | 0.16 | 165 | 0.15 | 26 | 0.35 |  |  | ✓ |
| L02B9 | Other cytostatic hormone antagonists | 129 | 0.04 | 112 | 0.04 | 17 | 0.07 | 41 | 0.03 | 39 | 0.04 | 2 | 0.03 |  |  | ✓ |
| L03A1 | Colony-stimulating factors | 8390 | 2.47 | 7744 | 2.45 | 646 | 2.8 | 2421 | 2.05 | 2221 | 2.01 | 200 | 2.66 |  | ✓ | ✓ |
| L03A9 | All other immunostimulating agents excluding interferons | 331 | 0.1 | 295 | 0.09 | 36 | 0.16 | 96 | 0.08 | 90 | 0.08 | 6 | 0.08 |  |  | ✓ |
| L04B0 | ANTI-TNF PRODUCTS | 203 | 0.06 | 188 | 0.06 | 15 | 0.06 | 72 | 0.06 | 69 | 0.06 | 3 | 0.04 |  |  | ✓ |
| L04X0 | OTHER IMMUNOSUPPRESSANTS | 1714 | 0.5 | 1538 | 0.49 | 176 | 0.76 | 590 | 0.5 | 530 | 0.48 | 60 | 0.8 |  |  | ✓ |
| M01A1 | Anti-rheumatics, non-steroidal plain | 97354 | 28.67 | 91640 | 28.96 | 5714 | 24.73 | 30784 | 26.07 | 29202 | 26.41 | 1582 | 21.07 | ✓ | ✓ | ✓ |
| M01A3 | Coxibs, plain | 16155 | 4.76 | 15207 | 4.81 | 948 | 4.1 | 6251 | 5.29 | 5962 | 5.39 | 289 | 3.85 |  | ✓ | ✓ |
| M01C0 | SPECIFIC ANTI-RHEUMATIC AGENTS | 1426 | 0.42 | 1304 | 0.41 | 122 | 0.53 | 519 | 0.44 | 480 | 0.43 | 39 | 0.52 |  |  | ✓ |
| M02A0 | TOPICAL ANTI-RHEUMATICS AND ANALGESICS | 45373 | 13.36 | 41582 | 13.14 | 3791 | 16.41 | 14326 | 12.13 | 13219 | 11.96 | 1107 | 14.75 | ✓ | ✓ | ✓ |
| M03A0 | MUSCLE RELAXANTS, PERIPHERALLY ACTING | 71704 | 21.12 | 69409 | 21.94 | 2295 | 9.93 | 25477 | 21.58 | 24704 | 22.34 | 773 | 10.3 | ✓ | ✓ | ✓ |
| M03B0 | MUSCLE RELAXANTS, CENTRALLY ACTING | 3026 | 0.89 | 2801 | 0.89 | 225 | 0.97 | 867 | 0.73 | 805 | 0.73 | 62 | 0.83 |  |  | ✓ |
| M04A0 | ANTI-GOUT PREPARATIONS | 21290 | 6.27 | 19177 | 6.06 | 2113 | 9.14 | 7152 | 6.06 | 6433 | 5.82 | 719 | 9.58 | ✓ | ✓ | ✓ |
| M05B3 | Bisphosphonates for osteoporosis and related disorders | 6783 | 2 | 6166 | 1.95 | 617 | 2.67 | 2408 | 2.04 | 2230 | 2.02 | 178 | 2.37 |  | ✓ | ✓ |
| M05B4 | Bisphosphonates for tumour-related calcium disorders | 834 | 0.25 | 689 | 0.22 | 145 | 0.63 | 241 | 0.2 | 197 | 0.18 | 44 | 0.59 |  |  | ✓ |
| M05B9 | Other bone calcium regulators | 558 | 0.16 | 500 | 0.16 | 58 | 0.25 | 201 | 0.17 | 183 | 0.17 | 18 | 0.24 |  |  | ✓ |
| M05X0 | ALL OTHER MUSCULOSKELETAL PRODUCTS | 1628 | 0.48 | 1483 | 0.47 | 145 | 0.63 | 503 | 0.43 | 461 | 0.42 | 42 | 0.56 |  |  | ✓ |
| N01A1 | Inhalation general anaesthetics | 60072 | 17.69 | 58116 | 18.37 | 1956 | 8.46 | 21269 | 18.01 | 20628 | 18.66 | 641 | 8.54 | ✓ | ✓ | ✓ |
| N01A2 | Injectable general anaesthetics | 103642 | 30.53 | 99051 | 31.31 | 4591 | 19.87 | 37996 | 32.18 | 36407 | 32.93 | 1589 | 21.17 | ✓ | ✓ | ✓ |
| N01B1 | Anaesthetics local, medical injectables | 123513 | 36.38 | 117372 | 37.1 | 6141 | 26.58 | 43588 | 36.92 | 41570 | 37.6 | 2018 | 26.88 | ✓ | ✓ | ✓ |
| N01B3 | Anaesthetics local, topical | 113964 | 33.57 | 106567 | 33.68 | 7397 | 32.01 | 37151 | 31.46 | 34807 | 31.48 | 2344 | 31.22 | ✓ | ✓ | ✓ |
| N01B9 | Anaesthetics local, others | 338 | 0.1 | 320 | 0.1 | 18 | 0.08 | 54 | 0.05 | 46 | 0.04 | 8 | 0.11 |  |  | ✓ |
| N02A0 | NARCOTICS | 22932 | 6.75 | 20003 | 6.32 | 2929 | 12.68 | 7952 | 6.73 | 7006 | 6.34 | 946 | 12.6 | ✓ | ✓ | ✓ |
| N02B0 | NON-NARCOTICS AND ANTI-PYRETICS | 156012 | 45.95 | 145302 | 45.92 | 10710 | 46.35 | 57257 | 48.49 | 53628 | 48.5 | 3629 | 48.34 | ✓ | ✓ | ✓ |
| N02C1 | Antimigraine triptans | 192 | 0.06 | 180 | 0.06 | 12 | 0.05 | 55 | 0.05 | 51 | 0.05 | 4 | 0.05 |  |  | ✓ |
| N02C9 | All other anti-migraine preparations | 91 | 0.03 | 82 | 0.03 | 9 | 0.04 | 17 | 0.01 | 15 | 0.01 | 2 | 0.03 |  |  | ✓ |
| N03A0 | ANTI-EPILEPTICS | 21214 | 6.25 | 19262 | 6.09 | 1952 | 8.45 | 7280 | 6.17 | 6633 | 6 | 647 | 8.62 | ✓ | ✓ | ✓ |
| N04A0 | ANTI-PARKINSON DRUGS | 3718 | 1.1 | 3327 | 1.05 | 391 | 1.69 | 1206 | 1.02 | 1080 | 0.98 | 126 | 1.68 |  | ✓ | ✓ |
| N05A1 | Atypical antipsychotics | 10804 | 3.18 | 9622 | 3.04 | 1182 | 5.12 | 4530 | 3.84 | 4084 | 3.69 | 446 | 5.94 |  | ✓ | ✓ |
| N05A9 | Conventional antipsychotics | 17623 | 5.19 | 15748 | 4.98 | 1875 | 8.11 | 5603 | 4.75 | 5018 | 4.54 | 585 | 7.79 |  | ✓ | ✓ |
| N05B1 | Non-barbiturates, plain | 84087 | 24.77 | 77494 | 24.49 | 6593 | 28.53 | 27715 | 23.47 | 25639 | 23.19 | 2076 | 27.65 | ✓ | ✓ | ✓ |
| N05B3 | Barbiturates, plain | 1068 | 0.31 | 978 | 0.31 | 90 | 0.39 | 269 | 0.23 | 249 | 0.23 | 20 | 0.27 |  |  | ✓ |
| N05C0 | TRANQUILLISERS | 53011 | 15.61 | 49171 | 15.54 | 3840 | 16.62 | 17834 | 15.1 | 16638 | 15.05 | 1196 | 15.93 | ✓ | ✓ | ✓ |
| N06A3 | Mood stabilisers | 173 | 0.05 | 159 | 0.05 | 14 | 0.06 | 60 | 0.05 | 53 | 0.05 | 7 | 0.09 |  |  | ✓ |
| N06A4 | SSRI antidepressants | 2115 | 0.62 | 1916 | 0.61 | 199 | 0.86 | 648 | 0.55 | 588 | 0.53 | 60 | 0.8 |  |  | ✓ |
| N06A5 | SNRI antidepressants | 1485 | 0.44 | 1344 | 0.42 | 141 | 0.61 | 698 | 0.59 | 630 | 0.57 | 68 | 0.91 |  |  | ✓ |
| N06A9 | Antidepressants, all others | 5600 | 1.65 | 5001 | 1.58 | 599 | 2.59 | 3501 | 2.97 | 3176 | 2.87 | 325 | 4.33 |  | ✓ | ✓ |
| N06D0 | NOOTROPICS | 41 | 0.01 | 39 | 0.01 | 2 | 0.01 | 11 | 0.01 | 10 | 0.01 | 1 | 0.01 |  |  | ✓ |
| N07C0 | ANTIVERTIGO PRODUCTS | 4555 | 1.34 | 4252 | 1.34 | 303 | 1.31 | 1485 | 1.26 | 1400 | 1.27 | 85 | 1.13 |  | ✓ | ✓ |
| N07D1 | Anti-Alzheimer products, cholinesterase inhibitors | 5004 | 1.47 | 4515 | 1.43 | 489 | 2.12 | 1722 | 1.46 | 1530 | 1.38 | 192 | 2.56 |  | ✓ | ✓ |
| N07D9 | All other anti-Alzheimer products | 1686 | 0.5 | 1486 | 0.47 | 200 | 0.87 | 716 | 0.61 | 633 | 0.57 | 83 | 1.11 |  |  | ✓ |
| N07F0 | DRUGS USED IN OPIOID DEPENDENCE | 1960 | 0.58 | 1866 | 0.59 | 94 | 0.41 | 538 | 0.46 | 517 | 0.47 | 21 | 0.28 |  |  | ✓ |
| N07X0 | ALL OTHER CNS DRUGS | 74580 | 21.97 | 71321 | 22.54 | 3259 | 14.1 | 26563 | 22.5 | 25503 | 23.07 | 1060 | 14.12 | ✓ | ✓ | ✓ |
| P01B0 | ANTHELMINTICS, EXCLUDING SCHISTOSOMICIDES | 136 | 0.04 | 123 | 0.04 | 13 | 0.06 | 94 | 0.08 | 79 | 0.07 | 15 | 0.2 |  |  | ✓ |
| P01G0 | OTHER ANTI-PARASITIC AGENTS | 187 | 0.06 | 171 | 0.05 | 16 | 0.07 | 67 | 0.06 | 62 | 0.06 | 5 | 0.07 |  |  | ✓ |
| R01A1 | Nasal corticosteroids without anti-infectives | 773 | 0.23 | 720 | 0.23 | 53 | 0.23 | 288 | 0.24 | 277 | 0.25 | 11 | 0.15 |  |  | ✓ |
| R01A4 | Nasal anti-infectives without corticosteroids | 161 | 0.05 | 145 | 0.05 | 16 | 0.07 | 53 | 0.04 | 49 | 0.04 | 4 | 0.05 |  |  | ✓ |
| R01A6 | Nasal antiallergic agents | 103 | 0.03 | 99 | 0.03 | 4 | 0.02 | 17 | 0.01 | 17 | 0.02 | 0 | 0 |  |  | ✓ |
| R01A7 | Nasal decongestants | 1845 | 0.54 | 1688 | 0.53 | 157 | 0.68 | 484 | 0.41 | 445 | 0.4 | 39 | 0.52 |  |  | ✓ |
| R02A0 | THROAT PREPARATIONS | 16632 | 4.9 | 15439 | 4.88 | 1193 | 5.16 | 4443 | 3.76 | 4151 | 3.75 | 292 | 3.89 |  | ✓ | ✓ |
| R03A2 | B2-agonists, systemic | 14034 | 4.13 | 12900 | 4.08 | 1134 | 4.91 | 4043 | 3.42 | 3745 | 3.39 | 298 | 3.97 |  | ✓ | ✓ |
| R03A3 | Long-acting B2-agonists, inhalant | 684 | 0.2 | 589 | 0.19 | 95 | 0.41 | 160 | 0.14 | 141 | 0.13 | 19 | 0.25 |  |  | ✓ |
| R03A4 | Short-acting B2-agonists, inhalant | 25320 | 7.46 | 23269 | 7.35 | 2051 | 8.88 | 7923 | 6.71 | 7326 | 6.63 | 597 | 7.95 | ✓ | ✓ | ✓ |
| R03B2 | Xanthines, systemic | 4288 | 1.26 | 3788 | 1.2 | 500 | 2.16 | 1128 | 0.96 | 1015 | 0.92 | 113 | 1.51 |  |  | ✓ |
| R03C1 | Non-steroidal respiratory anti-inflammatories, inhalant | 5273 | 1.55 | 4954 | 1.57 | 319 | 1.38 | 1646 | 1.39 | 1556 | 1.41 | 90 | 1.2 |  | ✓ | ✓ |
| R03C2 | Non-steroidal respiratory anti-inflammatories, systemic | 469 | 0.14 | 449 | 0.14 | 20 | 0.09 | 106 | 0.09 | 98 | 0.09 | 8 | 0.11 |  |  | ✓ |
| R03D1 | Corticoids, inhalant | 2165 | 0.64 | 1968 | 0.62 | 197 | 0.85 | 631 | 0.53 | 568 | 0.51 | 63 | 0.84 |  |  | ✓ |
| R03F1 | B2-agonist and corticoid combinations, inhalant | 5029 | 1.48 | 4527 | 1.43 | 502 | 2.17 | 1771 | 1.5 | 1592 | 1.44 | 179 | 2.38 |  | ✓ | ✓ |
| R03J2 | Antileukotriene anti-asthmatics, systemic | 11475 | 3.38 | 10623 | 3.36 | 852 | 3.69 | 3881 | 3.29 | 3594 | 3.25 | 287 | 3.82 |  | ✓ | ✓ |
| R03K1 | Short-acting anticholinergics, plain, inhalant | 124 | 0.04 | 109 | 0.03 | 15 | 0.06 | 39 | 0.03 | 35 | 0.03 | 4 | 0.05 |  |  | ✓ |
| R03K2 | Long-acting anticholinergics, plain, inhalant | 2947 | 0.87 | 2581 | 0.82 | 366 | 1.58 | 802 | 0.68 | 719 | 0.65 | 83 | 1.11 |  |  | ✓ |
| R03L2 | Long-acting anticholinergic combinations with long-acting B2-agonists, inhalant | 1595 | 0.47 | 1391 | 0.44 | 204 | 0.88 | 712 | 0.6 | 629 | 0.57 | 83 | 1.11 |  |  | ✓ |
| R03M0 | INTERLEUKIN INHIBITOR ANTI-ASTHMATICS | 187 | 0.06 | 174 | 0.05 | 13 | 0.06 | 59 | 0.05 | 51 | 0.05 | 8 | 0.11 |  |  | ✓ |
| R03X2 | All other anti-asthma and COPD products, systemic | 21545 | 6.35 | 20694 | 6.54 | 851 | 3.68 | 7171 | 6.07 | 6905 | 6.25 | 266 | 3.54 | ✓ | ✓ | ✓ |
| R05A0 | COLD PREPARATIONS WITHOUT ANTI-INFECTIVES | 2843 | 0.84 | 2667 | 0.84 | 176 | 0.76 | 683 | 0.58 | 642 | 0.58 | 41 | 0.55 |  |  | ✓ |
| R05C0 | EXPECTORANTS | 42916 | 12.64 | 39307 | 12.42 | 3609 | 15.62 | 13669 | 11.58 | 12604 | 11.4 | 1065 | 14.19 | ✓ | ✓ | ✓ |
| R05D1 | Plain antitussives | 63704 | 18.76 | 60697 | 19.18 | 3007 | 13.01 | 22914 | 19.41 | 21925 | 19.83 | 989 | 13.17 | ✓ | ✓ | ✓ |
| R05D2 | Antitussives in combinations | 1335 | 0.39 | 1188 | 0.38 | 147 | 0.64 | 375 | 0.32 | 342 | 0.31 | 33 | 0.44 |  |  | ✓ |
| R06A0 | SYSTEMIC ANTIHISTAMINES | 28103 | 8.28 | 25803 | 8.16 | 2300 | 9.95 | 9286 | 7.86 | 8535 | 7.72 | 751 | 10 | ✓ | ✓ | ✓ |
| R07A0 | RESPIRATORY STIMULANTS | 267 | 0.08 | 253 | 0.08 | 14 | 0.06 | 91 | 0.08 | 90 | 0.08 | 1 | 0.01 |  |  | ✓ |
| R07X0 | ALL OTHER RESPIRATORY SYSTEM PRODUCTS | 634 | 0.19 | 573 | 0.18 | 61 | 0.26 | 165 | 0.14 | 149 | 0.13 | 16 | 0.21 |  |  | ✓ |
| S01A0 | OPHTHALMOLOGICAL ANTI-INFECTIVES | 4903 | 1.44 | 4469 | 1.41 | 434 | 1.88 | 1451 | 1.23 | 1330 | 1.2 | 121 | 1.61 |  | ✓ | ✓ |
| S01B0 | OPHTHALMOLOGICAL CORTICOSTEROIDS | 1726 | 0.51 | 1620 | 0.51 | 106 | 0.46 | 478 | 0.4 | 445 | 0.4 | 33 | 0.44 |  |  | ✓ |
| S01C1 | Ophthalmological corticosteroid and anti-infective combinations | 491 | 0.14 | 465 | 0.15 | 26 | 0.11 | 137 | 0.12 | 131 | 0.12 | 6 | 0.08 |  |  | ✓ |
| S01D0 | OPHTHALMOLOGICAL ANTIVIRAL AGENTS | 166 | 0.05 | 153 | 0.05 | 13 | 0.06 | 49 | 0.04 | 46 | 0.04 | 3 | 0.04 |  |  | ✓ |
| S01E1 | Miotics and antiglaucoma preparations, systemic | 1267 | 0.37 | 1207 | 0.38 | 60 | 0.26 | 386 | 0.33 | 361 | 0.33 | 25 | 0.33 |  |  | ✓ |
| S01E2 | Miotics and antiglaucoma preparations, topical | 3016 | 0.89 | 2719 | 0.86 | 297 | 1.29 | 961 | 0.81 | 872 | 0.79 | 89 | 1.19 |  |  | ✓ |
| S01F0 | MYDRIATICS AND CYCLOPLEGICS | 2695 | 0.79 | 2513 | 0.79 | 182 | 0.79 | 794 | 0.67 | 750 | 0.68 | 44 | 0.59 |  |  | ✓ |
| S01G1 | Ocular anti-allergics, antihistamines | 294 | 0.09 | 277 | 0.09 | 17 | 0.07 | 79 | 0.07 | 72 | 0.07 | 7 | 0.09 |  |  | ✓ |
| S01G2 | Ocular anti-allergics, mast cell stabilisers | 192 | 0.06 | 174 | 0.05 | 18 | 0.08 | 64 | 0.05 | 60 | 0.05 | 4 | 0.05 |  |  | ✓ |
| S01G3 | Ocular anti-allergics, multiple action | 390 | 0.11 | 356 | 0.11 | 34 | 0.15 | 120 | 0.1 | 114 | 0.1 | 6 | 0.08 |  |  | ✓ |
| S01G9 | Other similar ocular products | 145 | 0.04 | 128 | 0.04 | 17 | 0.07 | 45 | 0.04 | 42 | 0.04 | 3 | 0.04 |  |  | ✓ |
| S01H0 | OPHTHALMOLOGICAL LOCAL ANAESTHETICS | 790 | 0.23 | 741 | 0.23 | 49 | 0.21 | 347 | 0.29 | 325 | 0.29 | 22 | 0.29 |  |  | ✓ |
| S01K1 | Artifical tears and ocular lubricants | 4932 | 1.45 | 4493 | 1.42 | 439 | 1.9 | 1570 | 1.33 | 1437 | 1.3 | 133 | 1.77 |  | ✓ | ✓ |
| S01K9 | Dry eye products, other | 554 | 0.16 | 516 | 0.16 | 38 | 0.16 | 172 | 0.15 | 159 | 0.14 | 13 | 0.17 |  |  | ✓ |
| S01M0 | EYE TONICS AND EYE VITAMINS | 670 | 0.2 | 599 | 0.19 | 71 | 0.31 | 228 | 0.19 | 212 | 0.19 | 16 | 0.21 |  |  | ✓ |
| S01N2 | Preparations to prevent cataract and anticataractogenics, topical | 1899 | 0.56 | 1733 | 0.55 | 166 | 0.72 | 559 | 0.47 | 517 | 0.47 | 42 | 0.56 |  |  | ✓ |
| S01R0 | OPHTHALMIC NON-STEROIDAL ANTI-INFLAMMATORIES | 1744 | 0.51 | 1586 | 0.5 | 158 | 0.68 | 608 | 0.51 | 552 | 0.5 | 56 | 0.75 |  |  | ✓ |
| S01S1 | Viscoelastic substances | 343 | 0.1 | 328 | 0.1 | 15 | 0.06 | 144 | 0.12 | 137 | 0.12 | 7 | 0.09 |  |  | ✓ |
| S01S9 | Other surgical aids | 506 | 0.15 | 480 | 0.15 | 26 | 0.11 | 180 | 0.15 | 171 | 0.15 | 9 | 0.12 |  |  | ✓ |
| S01T0 | OPHTHALMOLOGICAL DIAGNOSTIC AGENTS | 1347 | 0.4 | 1256 | 0.4 | 91 | 0.39 | 417 | 0.35 | 390 | 0.35 | 27 | 0.36 |  |  | ✓ |
| S01X2 | Other ophthalmologicals, topical | 101 | 0.03 | 96 | 0.03 | 5 | 0.02 | 29 | 0.02 | 27 | 0.02 | 2 | 0.03 |  |  | ✓ |
| S02A0 | OTIC ANTI-INFECTIVES | 1147 | 0.34 | 1086 | 0.34 | 61 | 0.26 | 293 | 0.25 | 264 | 0.24 | 29 | 0.39 |  |  | ✓ |
| S03B0 | EYE/EAR CORTICOSTEROIDS | 1135 | 0.33 | 1071 | 0.34 | 64 | 0.28 | 350 | 0.3 | 330 | 0.3 | 20 | 0.27 |  |  | ✓ |
| S03C0 | EYE/EAR CORTICOSTEROID/ANTI-INFECTIVE COMBINATIONS | 295 | 0.09 | 273 | 0.09 | 22 | 0.1 | 103 | 0.09 | 95 | 0.09 | 8 | 0.11 |  |  | ✓ |
| T01A0 | LOW OSMOLAR ANGIO-UROGRAPHY | 79683 | 23.47 | 74551 | 23.56 | 5132 | 22.21 | 27340 | 23.15 | 25731 | 23.27 | 1609 | 21.43 | ✓ | ✓ | ✓ |
| T01B0 | IONIC ANGIO-UROGRAPHY | 23249 | 6.85 | 20887 | 6.6 | 2362 | 10.22 | 7540 | 6.39 | 6846 | 6.19 | 694 | 9.24 | ✓ | ✓ | ✓ |
| T01C0 | GASTROENTEROGRAPHY | 1639 | 0.48 | 1459 | 0.46 | 180 | 0.78 | 649 | 0.55 | 599 | 0.54 | 50 | 0.67 |  |  | ✓ |
| T01E0 | MRI AGENTS | 9617 | 2.83 | 8873 | 2.8 | 744 | 3.22 | 3078 | 2.61 | 2842 | 2.57 | 236 | 3.14 |  | ✓ | ✓ |
| T01F0 | ULTRASOUND AGENTS | 438 | 0.13 | 425 | 0.13 | 13 | 0.06 | 140 | 0.12 | 130 | 0.12 | 10 | 0.13 |  |  | ✓ |
| T01G0 | RADIODIAGNOSTIC AGENTS | 7625 | 2.25 | 7326 | 2.32 | 299 | 1.29 | 2501 | 2.12 | 2383 | 2.16 | 118 | 1.57 |  | ✓ | ✓ |
| T01X0 | OTHER IMAGING AGENTS | 72 | 0.02 | 67 | 0.02 | 5 | 0.02 | 24 | 0.02 | 24 | 0.02 | 0 | 0 |  |  | ✓ |
| T02C0 | PREGNANCY AND OVULATION TESTS | 100 | 0.03 | 98 | 0.03 | 2 | 0.01 | 69 | 0.06 | 68 | 0.06 | 1 | 0.01 |  |  | ✓ |
| T02D2 | Diabetes tests, blood | 297 | 0.09 | 286 | 0.09 | 11 | 0.05 | 94 | 0.08 | 91 | 0.08 | 3 | 0.04 |  |  | ✓ |
| T02X0 | ALL OTHER DIAGNOSTIC TESTS | 3081 | 0.91 | 2927 | 0.93 | 154 | 0.67 | 931 | 0.79 | 895 | 0.81 | 36 | 0.48 |  |  | ✓ |
| T02X2 | All other diagnostic tests, blood | 81 | 0.02 | 81 | 0.03 | 0 | 0 | 26 | 0.02 | 26 | 0.02 | 0 | 0 |  |  | ✓ |
| T02X9 | All other diagnostic tests | 2457 | 0.72 | 2308 | 0.73 | 149 | 0.64 | 903 | 0.76 | 854 | 0.77 | 49 | 0.65 |  |  | ✓ |
| V03B1 | Kanpo medicines | 28785 | 8.48 | 26072 | 8.24 | 2713 | 11.74 | 9839 | 8.33 | 9000 | 8.14 | 839 | 11.18 | ✓ | ✓ | ✓ |
| V03D0 | DETOXIFYING AGENTS FOR ANTINEOPLASTIC TREATMENT | 7069 | 2.08 | 6700 | 2.12 | 369 | 1.6 | 2034 | 1.72 | 1883 | 1.7 | 151 | 2.01 |  | ✓ | ✓ |
| V03E0 | ANTIDOTES | 15508 | 4.57 | 14593 | 4.61 | 915 | 3.96 | 5325 | 4.51 | 5045 | 4.56 | 280 | 3.73 |  | ✓ | ✓ |
| V03F0 | IRON-CHELATING AGENTS | 183 | 0.05 | 153 | 0.05 | 30 | 0.13 | 38 | 0.03 | 25 | 0.02 | 13 | 0.17 |  |  | ✓ |
| V03G1 | Hyperkalaemia products | 5209 | 1.53 | 4553 | 1.44 | 656 | 2.84 | 1844 | 1.56 | 1614 | 1.46 | 230 | 3.06 |  | ✓ | ✓ |
| V03G2 | Hyperphosphataemia products | 3151 | 0.93 | 2854 | 0.9 | 297 | 1.29 | 1080 | 0.91 | 973 | 0.88 | 107 | 1.43 |  |  | ✓ |
| V03H0 | ANTI-INFLAMMATORY ENZYMES | 18424 | 5.43 | 16977 | 5.37 | 1447 | 6.26 | 6120 | 5.18 | 5668 | 5.13 | 452 | 6.02 | ✓ | ✓ | ✓ |
| V03X6 | Homeopathic preparations (2) | 290 | 0.09 | 259 | 0.08 | 31 | 0.13 | 57 | 0.05 | 53 | 0.05 | 4 | 0.05 |  |  | ✓ |
| V03X9 | Other therapeutic preparations | 457 | 0.13 | 411 | 0.13 | 46 | 0.2 | 61 | 0.05 | 56 | 0.05 | 5 | 0.07 |  |  | ✓ |
| V06B0 | PROTEIN SUPPLEMENTS | 10540 | 3.1 | 8894 | 2.81 | 1646 | 7.12 | 3488 | 2.95 | 2993 | 2.71 | 495 | 6.59 |  | ✓ | ✓ |
| V06D0 | OTHER NUTRIENTS | 642 | 0.19 | 585 | 0.18 | 57 | 0.25 | 291 | 0.25 | 268 | 0.24 | 23 | 0.31 |  |  | ✓ |
| V07A0 | ALL OTHER NON-THERAPEUTIC PRODUCTS | 49335 | 14.53 | 45696 | 14.44 | 3639 | 15.75 | 16601 | 14.06 | 15419 | 13.95 | 1182 | 15.75 | ✓ | ✓ | ✓ |

EPHMRA = European Pharmaceutical Market Research Association.
