## Supplementary Table S5 for "Comparison of machine-learning and logistic regression models to predict 30-day unplanned readmission: a development and validation study"

| **Supplementary Table S5. Details and distribution of blood-test results** | | | | | | |
| --- | --- | --- | --- | --- | --- | --- |
| Variables | Derivation dataset (N=339,513) | | | Validation dataset (N=118,074) | | |
|  | Total  N=339,513 | Outcome (-)  N=316,405 | Outcome (+)  N=23,108 | Total  N=118,074 | Outcome (-)  N=110,567 | Outcome (+)  N=7,507 |
| White blood cell count (/μL), mean±SD,  median (IQR) | 6758±4082  6100 (4700-7970) | 6742±3735  6100 (4700-7940) | 6982±7333  6100 (4600-8200) | 6752±4047  6100 (4730-7910) | 6741±3911  6100 (4750-7900) | 6920±5680  6080 (4600-8120) |
| Hemoglobin (g/dL), mean±SD,  median (IQR) | 11.8±2.0  11.9 (10.5-13.2) | 11.9±2.0  12.0 (10.6-13.3) | 11.0±2.1  10.9 (9.5-12.4) | 11.9±2.0  12.0 (10.5-13.3) | 12.0±2.0  12.0 (10.6-13.3) | 11.0±2.1  11.0 (9.5-12.5) |
| Platelet count (10^4^/μL), mean±SD,  median (IQR) | 24.6±10.9  22.9 (17.6-29.6) | 24.7±10.8  22.9 (17.7-29.6) | 23.8±12.3  22.2 (15.8-29.9) | 25.1±11.0  23.3 (18.0-30.0) | 25.2±11.0  23.3 (18.1-30.0) | 24.3±12.0  22.6 (16.4-30.3) |
| Sodium (mEq/L), mean±SD,  median (IQR) | 139.5±3.4  140 (138-142) | 139.6±3.3  140 (138-142) | 138.4±4.1  139 (136-141) | 139.7±3.4  140 (138-142) | 139.8±3.3  140 (138-142) | 138.7±4.1  139 (137-141) |
| Potassium (mEq/L), mean±SD,  median (IQR) | 4.2±0.5  4.2 (3.9-4.5) | 4.2±0.5  4.2 (3.9-4.4) | 4.2±0.6  4.2 (3.8-4.5) | 4.2±0.5  4.2 (3.9-4.5) | 4.2±0.5  4.2 (3.9-4.5) | 4.2±0.6  4.2 (3.8-4.5) |
| Chloride (mEq/L), mean±SD,  median (IQR) | 104.3±3.8  105 (102-107) | 104.4±3.7  105 (102-107) | 103.3±4.7  104 (101-106) | 104.3±3.8  105 (102-107) | 104.3±3.7  105 (102-107) | 103.3±4.6  104 (101-106) |
| Aspartate aminotransferase (IU/L), mean±SD,  median (IQR) | 27.9±30.0  22 (17-31) | 27.6±29.8  22 (17-30) | 31.2±33.2  23 (17-33) | 27.7±28.9  21 (17-30) | 27.4±27.1  21 (17-30) | 31.6±48.3  23 (17-33) |
| Alanine aminotransferase (IU/L), mean±SD,  median (IQR) | 25.8±34.8  17 (12-28) | 25.8±34.7  17 (12-28) | 26.3±35.8  17 (11-28) | 25.5±34.9  17 (11-27) | 25.4±34.6  17 (11-27) | 26.4±39.8  16 (11-28) |
| Serum creatinine (mg/dL), mean±SD,  median (IQR) | 0.95±1.28  0.71 (0.55-0.91) | 0.94±1.27  0.70 (0.55-0.90) | 1.09±1.41  0.74 (0.55-1.02) | 0.98±1.31  0.73 (0.56-0.93) | 0.97±1.30  0.72 (0.56-0.93) | 1.15±1.52  0.77 (0.57-1.06) |
| Blood urea nitrogen (mg/dL), mean±SD,  median (IQR) | 15.7±10.5  13.3 (10.0-18.0) | 15.5±10.2  13.2 (10.0-17.8) | 18.6±13.9  14.8 (10.5-21.5) | 15.9±10.8  13.5 (10.0-18.0) | 15.7±10.5  13.4 (10.0-18.0) | 19.0±14.6  15.0 (10.6-22.0) |

SD = standard deviation, IQR = interquartile range.
