## Supplementary Table S6 for "Comparison of machine-learning and logistic regression models to predict 30-day unplanned readmission: a development and validation study"

| **Supplementary Table S6. Hyperparameters in each model for the dataset with the largest number of variables including blood-test results (1543 variables)** | | | | | | | |
| --- | --- | --- | --- | --- | --- | --- | --- |
| Gradient-boosting decision tree | | Random forest | | Deep neural network | | Logistic regression | |
| Hyperparameter | Value | Hyperparameter | Value | Hyperparameter | Value | Hyperparameter | Value |
| col sample rate | 0.8 | number of trees | 50 | epochs | 21.7 | λ for LASSO | 0.000033 |
| col sample rate per tree | 0.8 | score tree interval | 5 | layer | 5 |  | |
| fold assignment | Modulo | fold assignment | Modulo | layer 1 units (type) | 4650 (input) |  |  |
| number of trees | 64 | stopping_metric | logloss | layer 2 units (type) | 100 (Rectifier) |  |  |
| max depth | 15 | stopping_tolerance | 0.0017 | layer 3 units (type) | 100 (Rectifier) |  |  |
| min rows | 100 | max depth | 20 | layer 5 units (type) | 2 (Softmax) |  |  |
| stopping_metric | logloss | sample_rate | 0.63 |  | |  |  |
| stopping tolerance | 0.0017 | col_sample_rate | 1 |  |  |  |  |
| distribution | bernoulli | col_sample_rate_per_tree | 1 |  |  |  |  |
| histogram type | UniformAdaptive | histogram_type | UniformAdaptive |  |  |  |  |
|  | | distribution | bernoulli |  |  |  |  |

LASSO = least absolute shrinkage and selection operator.
