## Supplementary Table S7 for "Comparison of machine-learning and logistic regression models to predict 30-day unplanned readmission: a development and validation study"

| **Supplementary Table S7. Discrimination ability (c-statistic) of each model for validation** | | | | |
| --- | --- | --- | --- | --- |
| C-statistic (95% confidence interval) and P-value* compared with LR-LASSO in the validation dataset | Gradient-boosting decision tree | Random forest | Deep neural network | LR-LASSO |
| Pattern 1: 102 variables, including binary variables that ≥5% of patients had, without blood-test results | 0.740 (0.735 – 0.746)  P<0.001 | 0.734 (0.729 – 0.740)  P<0.001 | 0.664 (0.658 – 0.670)  P<0.001 | 0.720 (0.714 – 0.726)  n/a |
| Pattern 2: 112 variables, including binary variables that ≥5% of patients had, with blood-test results | 0.751 (0.745 – 0.756)  P<0.001 | 0.742 (0.736 – 0.747)  P<0.001 | 0.728 (0.722 – 0.734)  P<0.001 | 0.734 (0.728 – 0.740)  n/a |
| Pattern 3: 296 variables, including binary variables that ≥1% of patients had, without blood-test results | 0.756 (0.751 – 0.762)  P<0.001 | 0.747 (0.742 – 0.753)  P<0.001 | 0.692 (0.685 – 0.698)  P<0.001 | 0.740 (0.734 – 0.746)  n/a |
| Pattern 4: 306 variables, including variables that ≥1% of patients had, with blood-test results | 0.759 (0.754 – 0.765)  P<0.001 | 0.753 (0.748 – 0.759)  P=0.013 | 0.737 (0.731 – 0.743)  P<0.001 | 0.749 (0.744 – 0.755)  n/a |
| Pattern 5: 1533 variables, including binary variables that ≥10 patients had, without blood-test results | 0.759 (0.753 – 0.764)  P<0.001 | 0.747 (0.741 – 0.752)  P=0.612 | 0.701 (0.695 – 0.707)  P<0.001 | 0.747 (0.742 – 0.753)  n/a |
| Pattern 6: 1543 variables, including binary variables that ≥10 patients had, with blood-test results | 0.764 (0.758 – 0.769)  P<0.001 | 0.751 (0.746 – 0.757)  P=0.010 | 0.720 (0.714 – 0.726)  P<0.001 | 0.755 (0.749 – 0.761)  n/a |

LR-LASSO = logistic regression with the least absolute shrinkage and selection operator.

*By Delong’s test.
