## Supplementary Table S8 for "Comparison of machine-learning and logistic regression models to predict 30-day unplanned readmission: a development and validation study"

| **Supplementary Table S8. Regression coefficients in logistic regression with the least absolute shrinkage and selection operator for the dataset with the largest number of variables including blood-test results (1543 variables)** | | |
| --- | --- | --- |
| No. | Variables finally used in the model | Coefficient |
| 1 | Age | 0.07 |
| 2 | Sex (men) | 0.05 |
| 3 | No. of hospitalization in the past year | 0.29 |
| 4 | Admission diagnosis category: Diseases of the blood, blood-forming organs and immune mechanism | 0.11 |
| 5 | Admission diagnosis category: Diseases of the nervous system | <0.01 |
| 6 | Admission diagnosis category: Diseases of the circulatory system | -0.04 |
| 7 | Admission diagnosis category: Diseases of the respiratory system | 0.02 |
| 8 | Admission diagnosis category: Diseases of the digestive system | 0.02 |
| 9 | Admission diagnosis category: Diseases of the musculoskeletal system and connective tissue | -0.09 |
| 10 | Admission diagnosis category: Diseases of the genitourinary system | -0.06 |
| 11 | Admission diagnosis category: Symptoms, signs and abnormal clinical and laboratory findings | 0.01 |
| 12 | Admission diagnosis category: Injury, poisoning and certain other consequences of external causes | -0.05 |
| 13 | Discharge place (discharge to nursing home) | 0.02 |
| 14 | A09 Other gastroenteritis and colitis of infectious and unspecified origin | 0.05 |
| 15 | A41 Other sepsis | 0.24 |
| 16 | A49 Bacterial infection of unspecified site | 0.02 |
| 17 | B18 Chronic viral hepatitis | 0.02 |
| 18 | B37 Candidiasis | 0.11 |
| 19 | B44 Aspergillosis | 0.18 |
| 20 | C15 Malignant neoplasm of oesophagus | 0.08 |
| 21 | C16 Malignant neoplasm of stomach | 0.35 |
| 22 | C17 Malignant neoplasm of small intestine | 0.23 |
| 23 | C18 Malignant neoplasm of colon | 0.09 |
| 24 | C20 Malignant neoplasm of rectum | 0.14 |
| 25 | C22 Malignant neoplasm of liver and intrahepatic bile ducts | 0.21 |
| 26 | C23 Malignant neoplasm of gallbladder | 0.33 |
| 27 | C24 Malignant neoplasm of other and unspecified parts of biliary tract | 0.64 |
| 28 | C25 Malignant neoplasm of pancreas | 0.55 |
| 29 | C34 Malignant neoplasm of bronchus and lung | 0.05 |
| 30 | C50 Malignant neoplasm of breast | 0.04 |
| 31 | C54 Malignant neoplasm of corpus uteri | 0.03 |
| 32 | C61 Malignant neoplasm of prostate | 0.05 |
| 33 | C67 Malignant neoplasm of bladder | 0.26 |
| 34 | C77 Secondary and unspecified malignant neoplasm of lymph nodes | 0.21 |
| 35 | C78 Secondary malignant neoplasm of respiratory and digestive organs | 0.48 |
| 36 | C79 Secondary malignant neoplasm of other and unspecified sites | 0.27 |
| 37 | C80 Malignant neoplasm, without specification of site | 0.11 |
| 38 | D46 Myelodysplastic syndromes | 0.36 |
| 39 | D47 Other neoplasms of uncertain or unknown behaviour of lymphoid, haematopoietic and related tissue | 0.18 |
| 40 | D50 Iron deficiency anaemia | 0.03 |
| 41 | D61 Other aplastic anaemias | 0.20 |
| 42 | D64 Other anaemias | 0.06 |
| 43 | D69 Purpura and other haemorrhagic conditions | 0.24 |
| 44 | D70 Agranulocytosis | 0.13 |
| 45 | E03 Other hypothyroidism | 0.05 |
| 46 | E11 Type 2 diabetes mellitus | 0.03 |
| 47 | E46 Unspecified protein-energy malnutrition | 0.10 |
| 48 | E63 Other nutritional deficiencies | 0.10 |
| 49 | E71 Disorders of branched-chain amino-acid metabolism and fatty-acid metabolism | <0.01 |
| 50 | E78 Disorders of lipoprotein metabolism and other lipidaemias | >-0.01 |
| 51 | E83 Disorders of mineral metabolism | 0.05 |
| 52 | E86 Volume depletion | 0.06 |
| 53 | E87 Other disorders of fluid, electrolyte and acid-base balance | 0.09 |
| 54 | E88 Other metabolic disorders | 0.07 |
| 55 | F03 Unspecified dementia | 0.10 |
| 56 | F20 Schizophrenia | 0.15 |
| 57 | F32 Depressive episode | 0.09 |
| 58 | F41 Other anxiety disorders | 0.02 |
| 59 | G20 Parkinson disease | 0.19 |
| 60 | G30 Alzheimer disease | 0.10 |
| 61 | G31 Other degenerative diseases of nervous system, not elsewhere classified | 0.03 |
| 62 | G40 Epilepsy | 0.30 |
| 63 | G47 Sleep disorders | 0.03 |
| 64 | G64 Other disorders of peripheral nervous system | 0.02 |
| 65 | G80 Cerebral palsy | 0.19 |
| 66 | I10 Essential (primary) hypertension | 0.03 |
| 67 | I20 Angina pectoris | 0.02 |
| 68 | I21 Acute myocardial infarction | 0.02 |
| 69 | I35 Nonrheumatic aortic valve disorders | 0.18 |
| 70 | I42 Cardiomyopathy | 0.15 |
| 71 | I47 Paroxysmal tachycardia | 0.08 |
| 72 | I48 Atrial fibrillation and flutter | 0.08 |
| 73 | I50 Heart failure | 0.08 |
| 74 | I62 Other nontraumatic intracranial haemorrhage | 0.07 |
| 75 | I69 Sequelae of cerebrovascular disease | 0.09 |
| 76 | J06 Acute upper respiratory infections of multiple and unspecified sites | 0.09 |
| 77 | J14 Pneumonia due to Haemophilus influenzae | 0.14 |
| 78 | J15 Bacterial pneumonia, not elsewhere classified | 0.06 |
| 79 | J18 Pneumonia, organism unspecified | 0.18 |
| 80 | J40 Bronchitis, not specified as acute or chronic | 0.07 |
| 81 | J42 Unspecified chronic bronchitis | 0.15 |
| 82 | J44 Other chronic obstructive pulmonary disease | 0.40 |
| 83 | J45 Asthma | 0.09 |
| 84 | J47 Bronchiectasis | 0.22 |
| 85 | J69 Pneumonitis due to solids and liquids | 0.31 |
| 86 | J84 Other interstitial pulmonary diseases | 0.43 |
| 87 | J90 Pleural effusion, not elsewhere classified | 0.20 |
| 88 | J93 Pneumothorax | 0.61 |
| 89 | J96 Respiratory failure, not elsewhere classified | 0.04 |
| 90 | J98 Other respiratory disorders | 0.18 |
| 91 | K12 Stomatitis and related lesions | 0.13 |
| 92 | K21 Gastro-oesophageal reflux disease | 0.02 |
| 93 | K22 Other diseases of oesophagus | 0.12 |
| 94 | K25 Gastric ulcer | 0.03 |
| 95 | K29 Gastritis and duodenitis | 0.02 |
| 96 | K31 Other diseases of stomach and duodenum | 0.18 |
| 97 | K56 Paralytic ileus and intestinal obstruction without hernia | 0.45 |
| 98 | K59 Other functional intestinal disorders | 0.02 |
| 99 | K62 Other diseases of anus and rectum | 0.09 |
| 100 | K64 Haemorrhoids and perianal venous thrombosis | 0.18 |
| 101 | K65 Peritonitis | 0.28 |
| 102 | K72 Hepatic failure, not elsewhere classified | 0.39 |
| 103 | K74 Fibrosis and cirrhosis of liver | 0.11 |
| 104 | K76 Other diseases of liver | 0.04 |
| 105 | K81 Cholecystitis | 0.30 |
| 106 | K83 Other diseases of biliary tract | 0.55 |
| 107 | K85 Acute pancreatitis | 0.03 |
| 108 | K86 Other diseases of pancreas | 0.18 |
| 109 | K91 Postprocedural disorders of digestive system, not elsewhere classified | 0.56 |
| 110 | K92 Other diseases of digestive system | 0.04 |
| 111 | L03 Cellulitis | 0.25 |
| 112 | L89 Decubitus ulcer and pressure area | 0.13 |
| 113 | M31 Other necrotizing vasculopathies | 0.06 |
| 114 | M54 Dorsalgia | 0.15 |
| 115 | M62 Other disorders of muscle | 0.09 |
| 116 | N10 Acute tubulo-interstitial nephritis | 0.26 |
| 117 | N12 Tubulo-interstitial nephritis, not specified as acute or chronic | 0.04 |
| 118 | N13 Obstructive and reflux uropathy | 0.41 |
| 119 | N17 Acute renal failure | 0.06 |
| 120 | N18 Chronic kidney disease | 0.09 |
| 121 | N19 Unspecified kidney failure | <0.01 |
| 122 | N28 Other disorders of kidney and ureter, not elsewhere classified | 0.06 |
| 123 | N30 Cystitis | 0.08 |
| 124 | N31 Neuromuscular dysfunction of bladder, not elsewhere classified | 0.09 |
| 125 | N39 Other disorders of urinary system | 0.20 |
| 126 | N40 Hyperplasia of prostate | 0.03 |
| 127 | Q90 Down syndrome | 0.33 |
| 128 | R03 Abnormal blood-pressure reading, without diagnosis | 0.02 |
| 129 | R04 Haemorrhage from respiratory passages | 0.32 |
| 130 | R06 Abnormalities of breathing | 0.01 |
| 131 | R11 Nausea and vomiting | -0.03 |
| 132 | R13 Dysphagia | 0.10 |
| 133 | R18 Ascites | 0.39 |
| 134 | R26 Abnormalities of gait and mobility | 0.19 |
| 135 | R31 Unspecified haematuria | 0.09 |
| 136 | R50 Fever of other and unknown origin | 0.04 |
| 137 | R52 Pain, not elsewhere classified | 0.20 |
| 138 | R56 Convulsions, not elsewhere classified | 0.02 |
| 139 | R57 Shock, not elsewhere classified | 0.02 |
| 140 | R58 Haemorrhage, not elsewhere classified | 0.02 |
| 141 | R60 Oedema, not elsewhere classified | 0.26 |
| 142 | R63 Symptoms and signs concerning food and fluid intake | 0.27 |
| 143 | S42 Fracture of shoulder and upper arm | 0.02 |
| 144 | T81 Complications of procedures, not elsewhere classified | 0.09 |
| 145 | T88 Other complications of surgical and medical care, not elsewhere classified | -0.05 |
| 146 | Z90 Acquired absence of organs, not elsewhere classified | 0.17 |
| 147 | Z93 Artificial opening status | 0.17 |
| 148 | K000 Wound treatment | 0.02 |
| 149 | K164 Intracranial hematoma removal (craniotomy) | 0.69 |
| 150 | K522 Esophageal stenosis extension | 0.21 |
| 151 | K611 Placement of implantable catheter for continuous infusion of antineoplastic agent into artery, vein or abdominal cavity | 0.49 |
| 152 | K635 thoracic fluid and ascites filtration and concentration re-infusion method | 0.25 |
| 153 | K655 Gastrectomy | 0.04 |
| 154 | K657 Total gastric resection | 0.39 |
| 155 | K662 Gastrointestinal anastomosis (Blanco-anastomosis included.) | 0.08 |
| 156 | K681 External gallbladder fistula construction | 0.17 |
| 157 | K688 Endoscopic biliary stenting | 0.19 |
| 158 | K775 Transcutaneous renal (pelvic) Fistulotomy | 0.15 |
| 159 | K783 Urethral stricture extension via urethra | <0.01 |
| 160 | K879 Uterine cancer ulcer surgery | 0.37 |
| 161 | K920 Blood transfusion | 0.06 |
| 162 | K931 Additional charge for ultrasonic coagulation and incision device | -0.04 |
| 163 | J000 Wound care | -0.03 |
| 164 | J008 Thoracentesis (including washing, injection and drainage) | 0.18 |
| 165 | J010 Abdominocentesis (including artificial insufflation, washing, infusion, and drainage) | 0.43 |
| 166 | J018 Sputum suction (per day) | 0.10 |
| 167 | J019 Continuous thoracic drainage (first day) | 0.07 |
| 168 | J024 Oxygen inhalation (per day) | 0.04 |
| 169 | J026 Intermittent positive pressure inhalation (per day) | 0.06 |
| 170 | J043 Phototherapy for neonatal hyperbilirubinemia (per day) | 0.12 |
| 171 | J045 Artificial respiration | 0.18 |
| 172 | J060 Cystourethral lavage (per day) | 0.14 |
| 173 | J061 Perineal pelvic lavage (unilateral) | 0.08 |
| 174 | J063 Placement of indwelling catheter | >-0.01 |
| 175 | A01A0 STOMATOLOGICALS | 0.05 |
| 176 | A01B0 MOUTH ANTIFUNGALS | 0.06 |
| 177 | A02A1 Plain antacids | 0.01 |
| 178 | A02A2 Plain antiflatulents and carminatives | -0.02 |
| 179 | A02A6 Antacids with other drugs | 0.17 |
| 180 | A02B1 H2 antagonists | -0.02 |
| 181 | A02B2 Proton pump inhibitors | >-0.01 |
| 182 | A02B9 All other antiulcerants | -0.01 |
| 183 | A03A0 PLAIN ANTISPASMODICS AND ANTICHOLINERGICS | -0.01 |
| 184 | A03F0 GASTROPROKINETICS | 0.03 |
| 185 | A03G0 GASTRO-INTESTINAL SENSORIMOTOR MODULATORS | 0.32 |
| 186 | A04A1 Serotonin antagonist antiemetics/antinauseants | -0.12 |
| 187 | A04A2 NK1 antagonist antiemetics/antinauseants | -0.08 |
| 188 | A05A2 Bile stone therapy | 0.05 |
| 189 | A06A2 Stimulant laxatives | 0.02 |
| 190 | A06A4 Enemas | 0.02 |
| 191 | A06A9 Other drugs for constipation | 0.03 |
| 192 | A06B1 Osmotic bowel cleansers | -0.04 |
| 193 | A06B2 Osmotic bowel cleansers with electrolytes | 0.05 |
| 194 | A07F0 ANTIDIARRHOEAL MICRO-ORGANISMS | 0.05 |
| 195 | A07H0 MOTILITY INHIBITORS | 0.05 |
| 196 | A09A0 DIGESTIVES, INCLUDING ENZYMES | 0.08 |
| 197 | A10C3 Human insulins and analogues, intermediate- or long-acting, combined with fast-acting | 0.07 |
| 198 | A10H0 SULPHONYLUREA ANTIDIABETICS | 0.01 |
| 199 | A10L0 ALPHA-GLUCOSIDASE INHIBITOR ANTIDIABETICS | 0.07 |
| 200 | A10M1 Glinide antidiabetics, plain | <0.01 |
| 201 | A10N1 DPP-IV inhibitor antidiabetics, plain | <0.01 |
| 202 | A11C2 Vitamin D | 0.01 |
| 203 | A11D4 Vitamin B1 combinations with vitamin B6 and/or vitamin B12 | 0.05 |
| 204 | A11D9 Other vitamin B1 combinations | 0.03 |
| 205 | A11G1 Plain vitamin C (including vitamin C salts) | -0.05 |
| 206 | A12B0 POTASSIUM PRODUCTS | >-0.01 |
| 207 | A12C2 Other mineral supplements | 0.17 |
| 208 | A16A0 OTHER ALIMENTARY TRACT AND METABOLISM PRODUCTS | 0.14 |
| 209 | B01A0 VITAMIN K ANTAGONISTS | 0.08 |
| 210 | B01B1 Unfractionated heparins | -0.07 |
| 211 | B01B2 Fractionated heparins | -0.04 |
| 212 | B01B3 Heparins for flushing | 0.06 |
| 213 | B01C1 Cyclo-oxygenase inhibitor platelet aggregation inhibitors | 0.01 |
| 214 | B02A1 Synthetic antifibrinolytics | -0.03 |
| 215 | B02B1 Vitamin K | 0.03 |
| 216 | B02D8 Platelet concentrates | 0.11 |
| 217 | B03A1 Plain iron | 0.01 |
| 218 | C01A1 Plain cardiac glycosides | 0.01 |
| 219 | C01B0 ANTI-ARRHYTHMICS | 0.03 |
| 220 | C01E0 NITRITES AND NITRATES | >-0.01 |
| 221 | C01X0 ALL OTHER CARDIAC PREPARATIONS | 0.09 |
| 222 | C03A1 Potassium-sparing agents plain | 0.06 |
| 223 | C03A2 Loop diuretics plain | 0.05 |
| 224 | C03A7 Vasopressin receptor antagonist diuretics | 0.24 |
| 225 | C07A0 BETA-BLOCKING AGENTS, PLAIN | 0.05 |
| 226 | C08A0 CALCIUM ANTAGONISTS, PLAIN | -0.01 |
| 227 | C09C0 ANGIOTENSIN-II ANTAGONISTS, PLAIN | -0.03 |
| 228 | C10A1 Statins (HMG-CoA reductase inhibitors) | -0.01 |
| 229 | D01A1 Topical dermatological antifungals | 0.00 |
| 230 | D06A0 TOPICAL ANTIBACTERIALS | 0.03 |
| 231 | D08A0 ANTISEPTICS AND DISINFECTANTS | 0.02 |
| 232 | G04C2 BPH alpha-adrenergic antagonists, plain | 0.01 |
| 233 | G04D4 Urinary incontinence products | 0.07 |
| 234 | H02A1 Injectable corticosteroids, plain | 0.01 |
| 235 | H02A2 Oral corticosteroids, plain | 0.12 |
| 236 | H04F0 ANTIPARATHYROID PRODUCTS | 0.01 |
| 237 | J01C1 Oral broad spectrum penicillins | 0.03 |
| 238 | J01C2 Injectable broad spectrum penicillins | 0.01 |
| 239 | J01D1 Oral cephalosporins | -0.02 |
| 240 | J01D2 Injectable cephalosporins | -0.02 |
| 241 | J01G1 Oral fluoroquinolones | -0.05 |
| 242 | J01P2 Penems and carbapenems | 0.03 |
| 243 | J02A0 SYSTEMIC AGENTS FOR FUNGAL INFECTIONS | 0.13 |
| 244 | J05D2 HEPATITIS B ANTIVIRALS | 0.02 |
| 245 | K01A3 1/2 - Electrolyte solutions | 0.02 |
| 246 | K01A4 1/3 - Electrolyte solutions | -0.01 |
| 247 | K01A7 Ringer's and Ringer's lactate solutions | -0.02 |
| 248 | K01A9 Other electrolyte solutions | 0.05 |
| 249 | K01B1 Sodium chloride solutions | -0.01 |
| 250 | K01B3 Carbohydrate solutions (<=10%) | 0.01 |
| 251 | K01C1 Solutions with one carbohydrate (>10%) | 0.05 |
| 252 | K01E1 Amino acid standard solutions | 0.03 |
| 253 | K01E4 Hepatic solutions | 0.20 |
| 254 | K01F1 Osmotic therapy | -0.14 |
| 255 | K02B0 STARCHES | 0.04 |
| 256 | K03A0 WHOLE BLOOD AND PLASMA FRACTIONS | 0.17 |
| 257 | L01B0 ANTIMETABOLITES | -0.06 |
| 258 | L01F0 PLATINUM ANTINEOPLASTICS | -0.13 |
| 259 | L01H9 Protein kinase inhibitor antineoplastics, other | 0.15 |
| 260 | M01A1 Anti-rheumatics, non-steroidal plain | 0.04 |
| 261 | M01A3 Coxibs, plain | 0.03 |
| 262 | M02A0 TOPICAL ANTI-RHEUMATICS AND ANALGESICS | 0.03 |
| 263 | M03A0 MUSCLE RELAXANTS, PERIPHERALLY ACTING | -0.23 |
| 264 | M05B3 Bisphosphonates for osteoporosis and related disorders | 0.01 |
| 265 | M05B4 Bisphosphonates for tumour-related calcium disorders | 0.04 |
| 266 | N01A1 Inhalation general anaesthetics | -0.01 |
| 267 | N01A2 Injectable general anaesthetics | -0.06 |
| 268 | N01B1 Anaesthetics local, medical injectables | -0.10 |
| 269 | N01B3 Anaesthetics local, topical | -0.04 |
| 270 | N02A0 NARCOTICS | 0.07 |
| 271 | N02B0 NON-NARCOTICS AND ANTI-PYRETICS | 0.03 |
| 272 | N03A0 ANTI-EPILEPTICS | 0.05 |
| 273 | N05A9 Conventional antipsychotics | 0.03 |
| 274 | N05B1 Non-barbiturates, plain | 0.01 |
| 275 | N05C0 TRANQUILLISERS | >-0.01 |
| 276 | N06A4 SSRI antidepressants | 0.01 |
| 277 | N06A9 Antidepressants, all others | 0.09 |
| 278 | N07D9 All other anti-Alzheimer products | 0.12 |
| 279 | N07X0 ALL OTHER CNS DRUGS | 0.02 |
| 280 | R02A0 THROAT PREPARATIONS | -0.04 |
| 281 | R03A2 B2-agonists, systemic | 0.04 |
| 282 | R03A3 Long-acting B2-agonists, inhalant | 0.05 |
| 283 | R03A4 Short-acting B2-agonists, inhalant | 0.05 |
| 284 | R03B2 Xanthines, systemic | 0.10 |
| 285 | R03D1 Corticoids, inhalant | 0.03 |
| 286 | R05C0 EXPECTORANTS | 0.04 |
| 287 | R05D1 Plain antitussives | 0.01 |
| 288 | R05D2 Antitussives in combinations | 0.01 |
| 289 | R06A0 SYSTEMIC ANTIHISTAMINES | 0.03 |
| 290 | S01A0 OPHTHALMOLOGICAL ANTI-INFECTIVES | 0.10 |
| 291 | S01E2 Miotics and antiglaucoma preparations, topical | 0.05 |
| 292 | S01R0 OPHTHALMIC NON-STEROIDAL ANTI-INFLAMMATORIES | 0.06 |
| 293 | T01A0 LOW OSMOLAR ANGIO-UROGRAPHY | -0.02 |
| 294 | T01B0 IONIC ANGIO-UROGRAPHY | 0.05 |
| 295 | T01C0 GASTROENTEROGRAPHY | 0.14 |
| 296 | T01E0 MRI AGENTS | 0.05 |
| 297 | V03B1 Kanpo medicines | 0.03 |
| 298 | V03D0 DETOXIFYING AGENTS FOR ANTINEOPLASTIC TREATMENT | -0.13 |
| 299 | V03G1 Hyperkalaemia products | 0.04 |
| 300 | V03H0 ANTI-INFLAMMATORY ENZYMES | -0.01 |
| 301 | V06B0 PROTEIN SUPPLEMENTS | 0.15 |
| 302 | V07A0 ALL OTHER NON-THERAPEUTIC PRODUCTS | >-0.01 |
| 303 | White blood cell count | 0.08 |
| 304 | Aspartate aminotransferase | 0.07 |
| 305 | Blood urea nitrogen | 0.09 |
